# Targeting the Microbiome With KB109 in Outpatients with Mild to Moderate COVID-19 Reduced Medically Attended Acute Care Visits and Improved Symptom Duration in Patients With Comorbidities

**DOI:** 10.1101/2021.03.26.21254422

**Authors:** John P. Haran, Yan Zheng, Katharine Knobil, Norma Alonzo Palma, Jonathan F. Lawrence, Mark A. Wingertzahn

**Affiliations:** Department of Emergency Medicine, Department of Microbiology and Physiological Systems, Program in Microbiome Dynamics, University of Massachusetts Medical School, UMass Memorial Medical Group, Worcester, MA; Kaleido Biosciences, Inc, Lexington, MA

**Author notes:** **Corresponding Author:** John P. Haran, MD, PhD, Associate Professor, Department of Emergency Medicine, Department of Microbiology and Physiological Systems, Clinical Director of the Center for Microbiome Research, University of Massachusetts Medical School, UMass Memorial Medical Group, 55 Lake Avenue North, Worcester, MA 01655.

## Abstract

**Introduction:** In 2020, the world experienced the beginning of the severe acute respiratory syndrome coronavirus 2 (SARS-CoV-2), also known as the coronavirus disease 2019 (COVID-19) pandemic. Mounting evidence indicates that the gut microbiome plays a role in host immune response to infections and, in turn, may have an impact on the disease trajectory of SARS-CoV2 infection. However, it remains to be established whether modulation of the microbiome can impact COVID-19–related symptomatology and patient outcomes. Therefore, we conducted a study designed to modulate the microbiome evaluating the safety and physiologic effects of KB109 combined with self-supportive care (SSC) vs SSC alone in non-hospitalized patients with mild to moderate COVID-19. KB109 is a novel synthetic glycan developed to increase the production of gut microbial metabolites that support immune system homeostasis through gut microbiome modulation. Our goal was to gain a better understanding of the safety of KB109, the natural course of COVID-19 symptomatology, and the possible role of the gut microbiome in patients with mild to moderate COVID-19.

**Methods:** Adult patients who tested positive for COVID-19 were randomized 1:1 to receive KB109 combined with SSC or SSC alone for 14 days and were then followed for an additional 21 days (35 days in total). Patients self-assessed their COVID-19–related symptoms (8 cardinal symptoms plus 5 additional symptoms) and self-reported comorbidities. The primary and secondary objectives were to evaluate the safety of KB109 plus SSC compared with that of SSC alone and to evaluate selected measures of health, respectively.

**Results:** Between July 2, 2020 and December 23, 2020, 350 patients were randomized to receive KB109 and SSC (n=174) or SSC alone (n=176). Overall, the most common comorbidities reported were hypertension (18.0% [63/350 patients]) followed by chronic lung disease (8.6% 30/350 patients). KB109 was well tolerated with most treatment-emergent adverse events being mild to moderate in severity. The administration of KB109 plus SSC reduced medically-attended visits (ie, hospitalization, emergency room visits, or urgent care visits) by 50.0% in the overall population and by 61.7% in patients with ≥1 comorbidity; in patients aged ≥45 years or with ≥1 comorbidity, medically-attended visits were reduced by 52.8%, In the SSC group, patients reporting ≥1 comorbidity had a longer median time to resolution of symptoms than those who reported no comorbidities at baseline (13 overall symptoms: 30 vs 21 days, respectively; hazard ratio [HR]=1.163 [95% CI, 0.723-1.872]; 8 cardinal symptoms: 21 vs 15 days, respectively; HR=1.283 [95% CI, 0.809-2.035]). In patients reporting ≥1 comorbidity, median time to resolution of symptoms was shorter in the KB109 plus SSC group compared with the SSC alone group (13 overall symptoms: 30 vs 21 days, respectively; HR=1.422 [95% CI, 0.898-2.250]; 8 cardinal symptoms: 17 vs 21 days, respectively; HR=1.574 [95% CI, 0.997-2.485]). In the KB109 plus SSC group, patients aged ≥45 years or with ≥1 comorbidity had a shorter median time to resolution of symptoms compared with SSC alone (overall 13 symptoms: 21 vs 31 days; HR=1.597 [95% CI, 1.064-2.398]).

**Conclusions:** Results from our study show that KB109 is well tolerated among patients with mild to moderate COVID-19. Patients with ≥1 comorbidity had a longer duration of COVID-19 symptoms than those without comorbidities. Moreover, in patients reporting ≥1 comorbidity or aged ≥45 years (at-risk population), administration of KB109 plus SSC improved median time to resolution of COVID-19–related symptoms and reduced the rate of medically-attended visits compared with SSC alone.

## Introduction

The severe acute respiratory syndrome coronavirus 2 (SARS-CoV-2), is a deadly virus and the cause of the coronavirus disease 2019 (COVID-19) pandemic.^1^ As of March 25, 2021, more than 124 million confirmed cases and more than 2.7 million deaths have been reported globally. Of these, the US accounts for >30 million confirmed cases and >545,000 deaths, higher than any other affected country.^2^ More than 1 year after the identification of COVID-19 and the start of the pandemic, understanding of symptomatology and disease course in both hospitalized and non-hospitalized patients continues to be refined.^1,3^

Despite the development of vaccines and various treatment methods, little information is available regarding the clinical course of a mild-to-moderate COVID-19 infection in the outpatient setting.^4^ Individuals infected with the SARS-CoV-2 virus may have a wide range of symptoms, including ones beyond the respiratory tract. Reported symptoms include fever, muscle pain, headache, shortness of breath, cough, sore throat, diarrhea, congestion/runny nose, fatigue, and loss of taste or smell.^5,6^ Many patients will recover from their initial symptoms over the course of 2 to 3 weeks. However, a sizable proportion of patients have a protracted clinical course, with upwards of 75% of hospitalized patients with COVID-19 still experiencing symptoms 6 months after their initial infection.^7,8^

According to the Centers for Disease Control and Prevention, individuals with specific underlying medical comorbidities are at an increased risk of severe disease due to SARS-CoV-2 infection.^9^ Comorbidities associated with a worse prognosis include hypertension, obesity, chronic lung disease, cardiovascular disease, and diabetes.^10^ Many different treatment options have been explored to improve COVID-19 outcomes (eg, monoclonal antibodies, steroids, antibiotics, immune regulators, and antiviral medications). There is extensive evidence that the gut microbiome plays a major role in regulation of innate and adaptive immunity, thereby impacting antiviral responses and potentially also the risk of excessive hyperinflammation in patients with COVID-19.^11–13^ The intestinal microbiome is a complex microbial ecosystem integrating environmental inputs with genetic and immune signals to affect a host’s metabolism, immunity, and response to infection.^14^ In addition to local effects on gut immune regulation by the resident microbiota, the immuno-modulatory impact in other organs, including on the pulmonary immune system, is now being recognized.^15^ The gut microbiota is reported to affect pulmonary health through a vital crosstalk between the gut microbiota and the lungs—the “gut-lung axis”.^16^ The gut-lung axis is thought to be bidirectional such that endotoxins and microbial metabolites can affect the lung via the bloodstream and inflammation occurrence in the lung can affect the gut microbiota. Recent studies have shown differences in gut microbiome diversity in patients with COVID-19 compared with those without COVID-19.^17,18^ Results from a recent study indicate that this altered microbiome may be involved in the severity of COVID-19 disease.^17^

Gut microbiome–derived metabolites influence peripheral inflammatory responses in addition to direct activation of host immune cells and pathways that impact the progression of pulmonary infections.^19^ Various plasma inflammatory cytokines and blood markers of inflammation have been associated with gut microbiota composition in patients with COVID-19.^17^ Because gut microbiota composition has been associated with COVID-19 severity and resulting tissue damage, alterations in the gut microbiota may influence COVID-19 severity through modulation of immune responses.^17^ Furthermore, an altered microbiome in healthy subjects may inflict an inflammatory state that increases the risk of COVID-19 susceptibility and disease severity.^20^ This study and others suggest that an altered microbiome (exhibited as a loss in overall commensal diversity and/or pathobiont enrichment) might contribute to unfavorable outcomes in respiratory infections.^13,17,19^ Data also support that gut-derived metabolites, like short-chain fatty acids (SCFAs), along with the migration of immune cells from the gut to the lung may play a role in pulmonary infections.^19^ The downstream consequences of this altered microbiome may be modulated by the SCFAs that are produced by the microbiota in the large intestine through anaerobic fermentation of dietary glycans.^21^ SCFAs modulate host inflammation, control adaptive immunity, and promote immune tolerance locally in the gut as well as systemically.^21^ A clinical report indicated that SCFAs and SCFA producing taxa are associated with a reduced risk of acquiring viral infections, including corona-viral infections, in a high-risk hematopoietic stem cell transplantation population.^12^ KB109, a novel synthetic glycan developed by Kaleido Biosciences, Inc, has been shown to increase SCFA production compared with a negative control in *ex vivo* studies, and importantly shows a consistent response across multiple healthy donor fecal communities.^22^ KB109 is related to a class of compounds that is generally recognized as safe (GRAS) or determined to be GRAS based on history of safe human exposure. This class is commonly accepted by regulators as safey for use in food and enables rapid advancement into human clinical studies.

Given the substantial data supporting a key role for both gut-derived metabolites (eg, SCFAs) and the direct interaction and migration of immune cells from gut to lung by the common mucosal immune system in pulmonary infections, we developed this non-investigational new drug study to examine the natural history of disease progression, the safety of KB109 and the influence of KB109 in patients with mild to moderate symptoms of COVID-19 in the outpatient setting.

## Methods

### Study Design

KB109 was evaluated in a virtual, randomized, controlled, multi-site, open-label study (NCT04414124, ClinicalTrials.org). Non-hospitalized patients with mild to moderate COVID-19 were randomized to receive either KB109 plus supportive self-care (SSC) or SSC alone (**Figure 1**). The primary objective was to evaluate the safety of KB109 when combined with SSC compared with that of SSC alone. The secondary objective was to evaluate selected measures of health in these patients.

**Figure 1.**
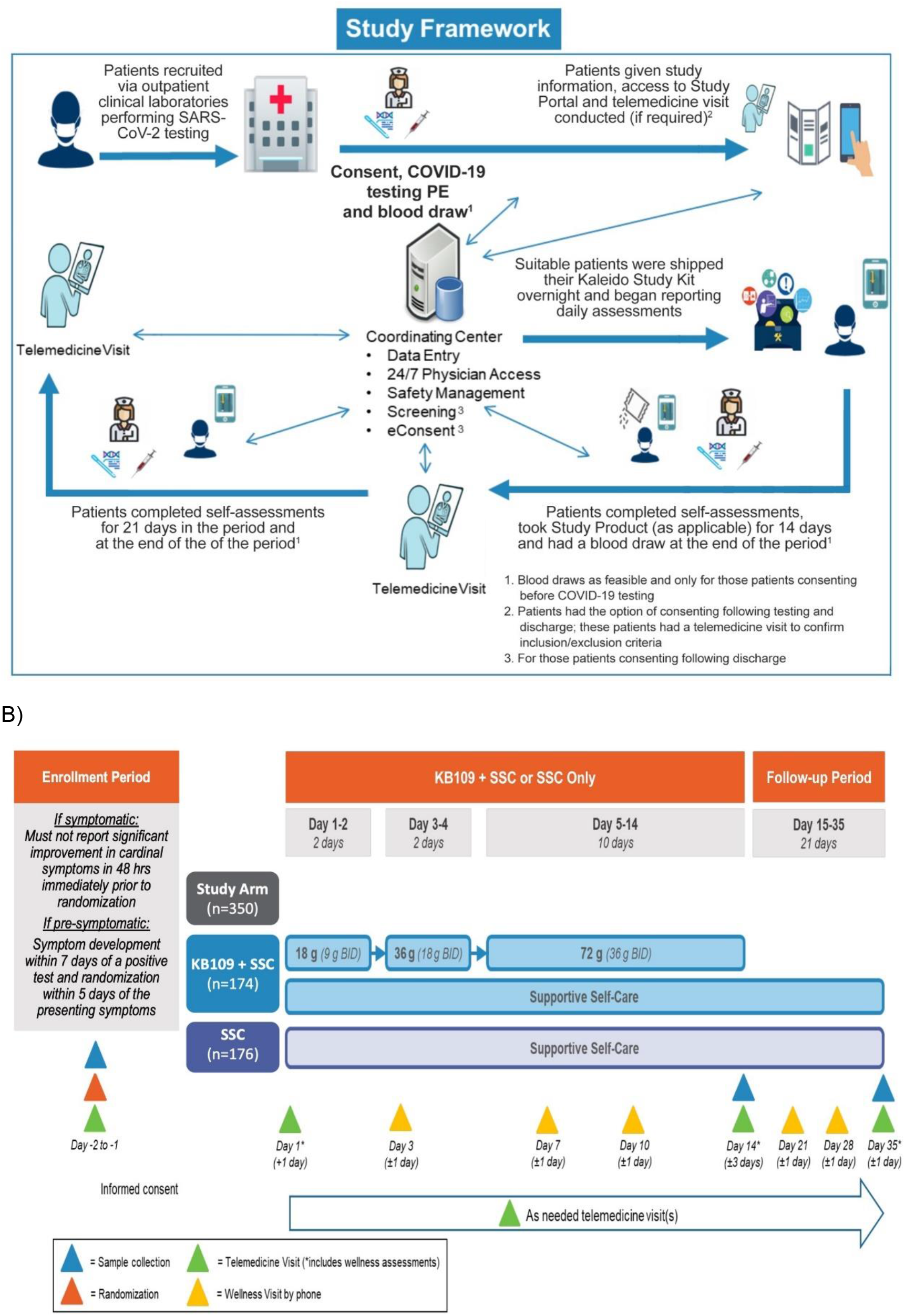
Evaluation of KB109 in non-hospitalized patients with mild to moderate COVID-19. A) The framework of this study allowed for virtual assessments and follow-up of patients. B) Study* overview and end points. *Non-IND clinical study conducted under regulations supporting research with food, evaluating safety, tolerability, and potential markers of human effect.

Patients were recruited through outpatient clinics or testing centers performing SARS-CoV-2 testing or through online portals (**Figure 1A**). Eligible patients were randomized in a 1:1 ratio to KB109 plus SSC or SSC alone groups via an interactive response technology system. Randomization was stratified by study site/center, age group (≥18 to <45 years, ≥45 to <65 years, ≥65 years), and comorbidity status (yes, no). After randomization, patients were given KB109 (if assigned) and the Kaleido At-home Study Kit (KaSK) containing dosing instructions (as applicable), a thermometer, a pulse oximeter, and telemedicine contact information. The study included an intake period (days 1-14) and a follow-up period (days 15-35) (**Figure 1B**). During the intake period, KB109 was reconstituted in water by the patients and consumed by the patient twice daily (BID), following an up-titration dosing schedule: 9 g BID on days 1 and 2; 18 g BID on days 3 and 4; and 36 g BID on days 5 through 14. SSC included over-the-counter cough, cold, and anti-pyretic medication that was used as necessary by patients in accordance with their respective drug facts label or as instructed by their healthcare provider.

### Eligibility Criteria

Enrolled patients were ≥18 years of age, had tested positive for COVID-19, and had mild to moderate COVID-19, and had self-reported outpatient management. Patients symptomatic at the time of COVID-19 testing must have reported new or worsening symptoms (**Table 1**) at baseline that were not present for more than 5 days; symptomatic patients were screened and randomized within 48 hours of a positive COVID-19 test result. Patients pre-symptomatic at the time of testing had to report new cardinal symptoms within 7 days of a positive test result and were screened and randomized within 5 days of developing symptoms. Patients were also required to have consistent internet or cellphone access with a data plan and access to a smartphone, tablet, or computer. Select exclusion criteria included patients likely to require hospitalization for COVID-19 or hospitalized for inpatient treatment or currently being evaluated for potential hospitalization at the time of informed consent for conditions other than COVID-19. Prebiotic and probiotic intake was not changed during the study. Full inclusion/exclusion criteria are available in the protocol (supplement 1).

**Table 1.** Patient-assessed COVID-19 Symptom Score

| **8 cardinal symptoms known to be associated with COVID-19:** | **5 additional symptoms associated with COVID-19** |
| --- | --- |
| — Fever<br>— Chills/Repeated shaking with chills<br>— Cough<br>— Shortness of breath<br>— Headache<br>— Muscle pain<br>— Anosmia/ageusia<br>— Sore throat | — GI disturbance/symptoms (other than diarrhea as this will be assessed separately)<br>— Diarrhea<br>— Fatigue<br>— Nasal congestion<br>— Chest tightness |

Patients will be instructed to rate their symptoms on a scale of 0-3 where:
0 = Absent (no symptoms evident)
1 = Mild (symptoms present but easily tolerated)
2 = Moderately severe (definite awareness of symptoms; bothersome but tolerable)
3 = Very severe (hard to tolerate; considerable interference with daily activity)
Abbreviation: COVID-19, coronavirus disease 2019.

### Study assessments

Patients assessments were primarily conducted virtually via telemedicine or telephone visits. Physical examinations were completed at screening (in person or via telemedicine) and at days 1, 14, and 35 (via telemedicine). Patient assessments were recorded in TrialPACE, a secure website housing the patient “diary.” Patients assessed and recorded their COVID-19 condition days 1 to 35; signs (temperature and oxygen saturation) were assessed and recorded daily from days 1 to 14 and on days 21, 28, and 35. Patient Global Impression on COVID-19 Condition (PGIC) responses were recorded on days 2 to 35 in which patients rated overall COVID-19 condition change over the past 24 hours using 7 categories (very much worse, much worse, minimally worse, no change, minimally improved, much improved, very much improved). Wellness visits by telephone on days 1, 3, 7, 10, and 14 were conducted by a healthcare provider (principal investigator or designee or a telemedicine vendor) to follow-up on the patient’s health status, to ascertain compliance with KB109 usage or completion of TrialPace™ questions, and to reeducate the patient on the importance of adherence to study instructions including KB109 usage (where applicable); on days 21, 28, and 35 these telephone visits followed-up on patient health status.

Healthcare utilization questions (supplement 2) were assessed on days 2 to 35 and patient-assessed bedrest time was recorded on days 1 to 35. For all patients in both groups, temperature and oxygen saturation were was measured and recorded as needed throughout the day before taking anti-pyretic medication.

Safety was monitored throughout the study by adverse events (AEs). COVID-19–related symptoms were not classified as treatment-emergent AEs (TEAEs) as long as they were within the normal day-to-day fluctuation or expected progression of the disease—not including hospitalizations—and were part of the clinical data of the disease that were being collected. Any patient experiencing a TEAE, significant worsening of COVID-19 symptoms, or intolerable gastrointestinal (GI) symptoms was evaluated by the principal investigator or designee via a telemedicine visit and referred as needed for emergent follow-up, or in the case of intolerable GI symptoms for patients in the SSC and KB109 group, for interruption and/or down-titration of KB109 dose to the previously tolerated level.

### Study Endpoints

The primary endpoint was the number of patients experiencing KB109-related TEAEs. Secondary endpoints included time to resolution of the overall 13 COVID-19–related symptoms and the 8 cardinal COVID-19–related symptoms (from day 1 until symptom score ≤ 1 and remained ≤ 1 for the rest of the intake and follow-up periods) (**Table 1**), proportion of patients with reduction from baseline in each of the 13 individual COVID-19–related symptoms at end of intake period (EOI) and follow-up, proportion of patients with baseline symptoms becoming absent at the end of intake EOI and follow-up for each of the 13 individual COVID-19–related symptoms, change from baseline to EOI in overall composite score of the 13 COVID-19–related symptoms and the 8 cardinal COVID-19–related symptoms, and the proportion of patients experiencing medically-attended visits (ie, hospitalization, emergency room visits, or urgent care visits) and healthcare utilizations during the intake and follow-up periods.

### Ethics Approval

The authors ensure this study was conducted in full conformity with Regulations for the Protection of Human Patients of Research codified in 45 CFR Part 46, 21 CFR Parts 50 and 56, and /or the principles in the International Council for Harmonization E6 (R2) Good Clinical Practice guideline. Each study site obtained institutional review board approval before study initiation and each patient provided written consent for study participation.

### Statistical Methods

Analyses were conducted using SAS Version 9.4 or later (SAS Institute, Inc, Cary, NC). A sample size of 350 to 400 patients was planned assuming a 15% attrition rate to provide 296 to 340 evaluable patients (148-170 per group). The full analysis set (FAS) included all randomized patients with both baseline and ≥1 post-baseline endpoint observation during the intake period. Patients were analyzed in the group to which they were randomized. The FAS was used with the Kaplan-Meier method to estimate the median time to resolution of the 13 overall COVID-19–related symptoms, 8 cardinal COVID-19–related symptoms, fever, and resolution rate specific to each group. The hazard ratio of the KB109 plus SSC group to SSC alone group along with 95% CI was estimated using Cox proportional hazards model, which includes factors for group, study site/center, age group (≥18 to <45 years, ≥45 to <65 years, ≥65 years), comorbidity status (yes, no), and baseline overall composite symptom score. The change from baseline to EOI in overall composite score of 13 COVID-19–related symptoms and overall composite score of 8 cardinal COVID-19–related symptoms were analyzed using analysis of covariance model.

Proportion of patients experiencing hospital admissions during the intake and follow-up periods were summarized by group using frequencies and percentages.

The safety analysis set (SAS) included all randomized patients. Patients who actually consumed any amount of KB109 were included in the KB109 plus SSC group; otherwise, patients were included in the SSC alone group. SAS was used to analyze AEs, proportion of patients experiencing hospital admissions (all cause, and COVID-19-related), measures collected from healthcare provider wellness visits and healthcare utilizations. TEAEs were coded using the most recent version of Medical Dictionary for Regulatory Activities (MedDRA) version 23.0. These endpoints were summarized using descriptive statistics.

### Subgroup analysis

Analysis for the secondary endpoint of time to resolution of overall 13 COVID-19–related symptoms and time to resolution of overall 8 cardinal COVID-19–related symptoms, change from baseline to EOI in overall composite score of 13 COVID-19–related symptoms and overall composite score of the 8 cardinal COVID-19–related symptoms was conducted for the following subgroups based on the FAS: study site/center, age group (≥18 to <45 years, ≥45 to <65 years, ≥65 years), comorbidity status (yes, no), baseline body mass index subgroup (<30 kg/m^2^, ≥30 kg/m^2^), and ethnicity (Hispanic or Latino, not Hispanic or Latino).

The Kaplan-Meier method was used to estimate median time to resolution and resolution rate specific to each group for each subgroup. A similar analysis of covariance model as the analysis of change from baseline to EOI in overall composite score of 13 COVID-19–related symptoms and overall composite score of the 8 cardinal COVID-19–related symptoms was performed. For each subgroup, summary statistics were provided.

## Results

Patients were enrolled between July 2, 2020 and December 23, 2020 at 16 study sites in the United States. A total of 350 patients were randomized to receive KB109 plus SSC (n=174) or SSC alone (n=176) (**Figure 2**). Five patients randomized to receive KB109 plus SSC did not take any KB109 and were analyzed for safety in the SSC alone group. Baseline characteristics were well matched between the groups, except for a higher proportion of patients reporting comorbidities at baseline with KB109 plus SSC compared with SSC alone (40.8% [69/169 patients] vs 35.5% [61/172 patients], respectively) (**Table 2**). Of note, most patients were white, of Hispanic or Latino ethnicity, and were <65 years of age.

**Table 2.**
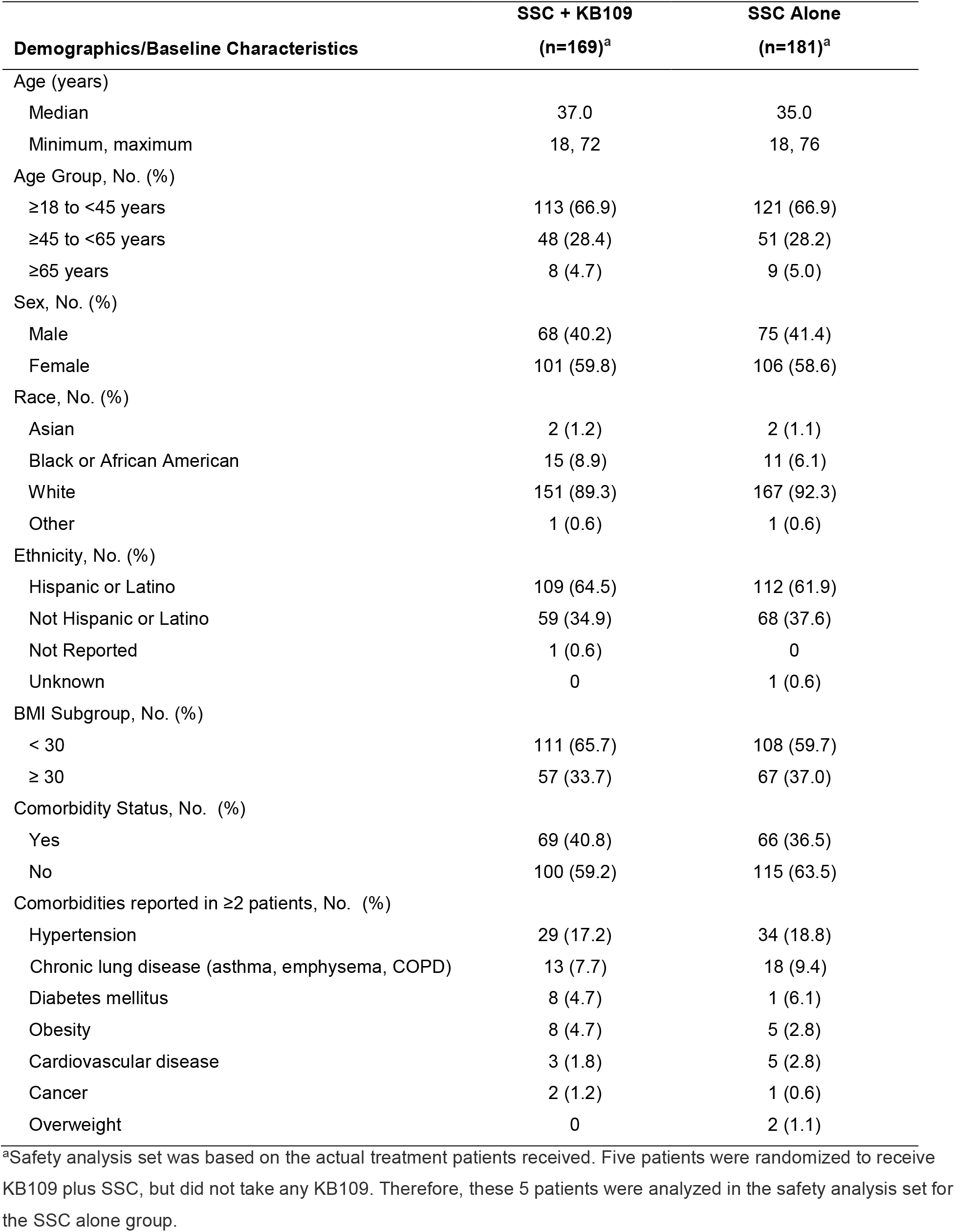
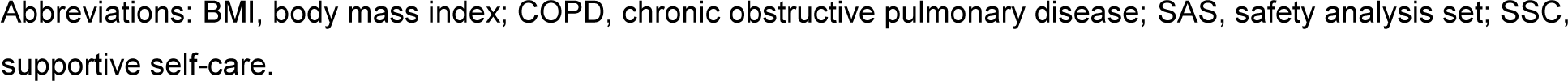
Demographics and Baseline Characteristics (SAS)

| **Demographics/Baseline Characteristics** | **SSC + KB109**<br>**(n=169)<sup>a</sup>** | **SSC Alone**<br>**(n=181)<sup>a</sup>** |
| --- | --- | --- |
| Age (years) |  |  |
| Median | 37.0 | 35.0 |
| Minimum, maximum | 18, 72 | 18, 76 |
| Age Group, No. (%) |  |  |
| ≥18 to <45 years | 113 (66.9) | 121 (66.9) |
| ≥45 to <65 years | 48 (28.4) | 51 (28.2) |
| ≥65 years | 8 (4.7) | 9 (5.0) |
| Sex, No. (%) |  |  |
| Male | 68 (40.2) | 75 (41.4) |
| Female | 101 (59.8) | 106 (58.6) |
| Race, No. (%) |  |  |
| Asian | 2 (1.2) | 2 (1.1) |
| Black or African American | 15 (8.9) | 11 (6.1) |
| White | 151 (89.3) | 167 (92.3) |
| Other | 1 (0.6) | 1 (0.6) |
| Ethnicity, No. (%) |  |  |
| Hispanic or Latino | 109 (64.5) | 112 (61.9) |
| Not Hispanic or Latino | 59 (34.9) | 68 (37.6) |
| Not Reported | 1 (0.6) | 0 |
| Unknown | 0 | 1 (0.6) |
| BMI Subgroup, No. (%) |  |  |
| < 30 | 111 (65.7) | 108 (59.7) |
| ≥ 30 | 57 (33.7) | 67 (37.0) |
| Comorbidity Status, No. (%) |  |  |
| Yes | 69 (40.8) | 66 (36.5) |
| No | 100 (59.2) | 115 (63.5) |
| Comorbidities reported in ≥2 patients, No. (%) |  |  |
| Hypertension | 29 (17.2) | 34 (18.8) |
| Chronic lung disease (asthma, emphysema, COPD) | 13 (7.7) | 18 (9.4) |
| Diabetes mellitus | 8 (4.7) | 1 (6.1) |
| Obesity | 8 (4.7) | 5 (2.8) |
| Cardiovascular disease | 3 (1.8) | 5 (2.8) |
| Cancer | 2 (1.2) | 1 (0.6) |
| Overweight | 0 | 2 (1.1) |

<sup>a</sup>Safety analysis set was based on the actual treatment patients received. Five patients were randomized to receive KB109 plus SSC, but did not take any KB109. Therefore, these 5 patients were analyzed in the safety analysis set for the SSC alone group.
Abbreviations: BMI, body mass index; COPD, chronic obstructive pulmonary disease; SAS, safety analysis set; SSC, supportive self-care.

**Figure 2.**
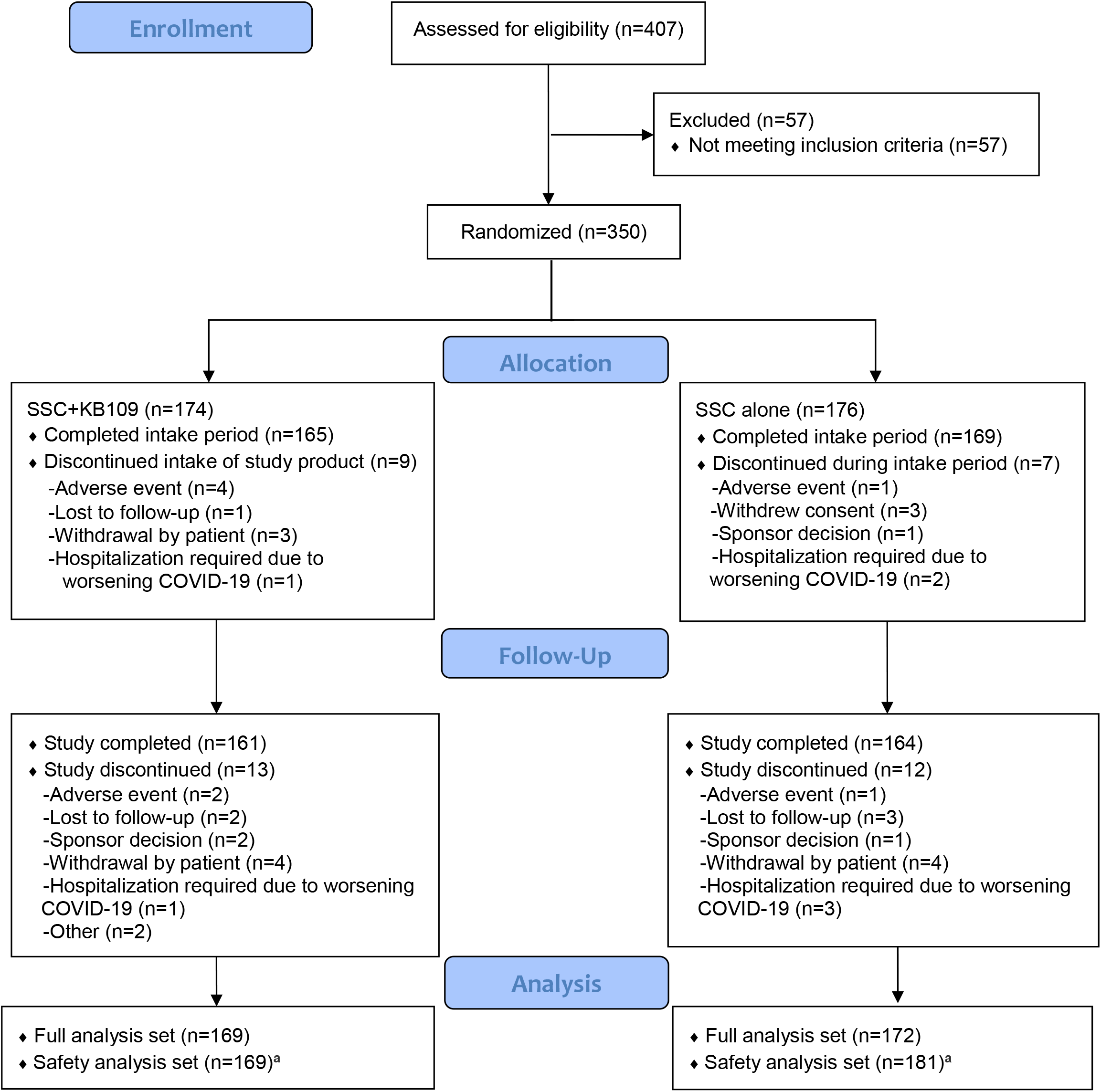
CONSORT flow diagram. ^a^Safety analysis set was based on the actual treatment patients received. Five patients were randomized to receive KB109 plus SSC, but did not take any KB109. Therefore, these 5 patients were analyzed in the safety analysis set for the SSC alone group.

### Primary Endpoint: Safety

As shown in **Table 3**, 36.1% (61/169) of patients receiving KB109 and SSC experienced ≥1 TEAE compared with 26.5% (48/181) of patients in the SSC-alone group. KB109 was well tolerated, with most TEAEs being mild to moderate in severity. GI-related TEAEs were most commonly reported and occurred more frequently in patients receiving KB109. All GI-related AEs were mild to moderate—with one leading to discontinuation (mild GI disturbance). Five patients receiving KB109 and SSC experienced TEAEs that led to discontinuation of KB109; in 2 patients, these TEAEs were related to KB109 (GI disorder and nocturia) whereas in 3 patients, these TEAEs were not related to KB109 (urinary tract infection, hypoxia, pneumonia, and ear infection). TEAEs leading to study discontinuation were reported in 2 patients receiving KB109 and SSC (hypoxia and COVID-19, neither related to KB109) and in 2 patients receiving SSC alone (COVID-19). Treatment emergent serious AEs were reported in 2 patients receiving KB109 and SSC (hypoxia, pneumonia, and COVID-19; none related to KB109) and 3 patients receiving SSC alone (COVID-19 and pneumonia). Only 1 death due to COVID-19 (2 weeks after withdrawal from the study in SSC-alone group) was reported.

**Table 3.** TEAEs Reported in >2 Patients (SAS)

|  | **SSC + KB109** | **SSC Alone** |
| --- | --- | --- |
|  | **No. (%)** | **No. (%)** |
|  | **(n=169)<sup>a</sup>** | **(n=181)<sup>a</sup>** |
| Patients with any TEAE | 61 (36.1) | 48 (26.5) |
| Patients with TEAEs by severity |  |  |
| Mild | 29 (17.2) | 17 (9.4) |
| Moderate | 28 (16.6) | 26 (14.4) |
| Severe | 4 (2.4) | 5 (2.8) |
| Patients with KB109-related TEAEs | 27 (16.0) | -- |
| Patients with Treatment-emergent Serious AEs | 2 (1.2) | 3 (1.7) |
| Patients with TEAEs leading to KB109 interruption | 2 (1.2) | -- |
| Patients with TEAEs leading to KB109 discontinuation | 5 (3.0) | -- |
| Patients with TEAEs Leading to Study Discontinuation | 2 (1.2) | 2 (1.1) |
| Patients with TEAEs leading to death | 0 | 0<sup>b</sup> |
| GI disorders |  |  |
| Diarrhea | 18 (10.7) | 6 (3.3) |
| Nausea | 6 (3.6) | 1 (0.6) |
| Abdominal pain | 5 (3.0) | 1 (0.6) |
| Abdominal distension | 4 (2.4) | 0 |
| Flatulence | 4 (2.4) | 0 |
| GI disorders | 3 (1.8) | 0 |
| Respiratory, thoracic and mediastinal disorders |  |  |
| Dyspnea | 8 (4.7) | 10 (5.5) |
| Oropharyngeal pain | 4 (2.4) | 7 (3.9) |
| Cough | 2 (1.2) | 8 (4.4) |
| Nasal congestion | 2 (1.2) | 3 (1.7) |
| Dyspnea exertional | 2 (1.2) | 1 (0.6) |
| Lower respiratory tract congestion | 0 | 2 (1.1) |
| Nervous system disorders |  |  |
| Ageusia | 4 (2.4) | 1 (0.6) |
| Anosmia | 4 (2.4) | 1 (0.6) |
| Headache | 3 (1.8) | 5 (2.8) |
| Infections and infestations |  |  |
| Pneumonia | 1 (0.6) | 4 (2.2) |
| General disorders and administration site conditions |  |  |
| Chills | 2 (1.2) | 2 (1.1) |
| Chest discomfort | 1 (0.6) | 2 (1.1) |
| Fatigue | 1 (0.6) | 2 (1.1) |
| Musculoskeletal and connective tissue disorders |  |  |
| Myalgia | 0 | 4 (2.2) |

<sup>a</sup>Safety analysis set was based on the actual treatment patients received. Five patients were randomized to receive KB109 plus SSC, but did not take any KB109. Therefore, these 5 patients were analyzed in the safety analysis set for the SSC alone group.
<sup>b</sup>One death due to COVID-19 was reported 2 weeks after withdrawal from the study in the SSC-alone group.
Abbreviations: AEs, adverse events; GI, gastrointestinal; SAS, safety analysis set; SSC, supportive self-care; TEAEs, treatment-emergent adverse events.

### Select Secondary Endpoints

#### Healthcare Utilization

In the overall study population, 4.1% (7/169) of patients receiving KB109 plus SSC group reported hospitalization, emergency room visits, or urgent care visits compared with 8.3% (15/181) of patients treated with SSC alone, resulting in a 50.0% reduction in these medically-attended visits. In the comorbidity subgroup analysis, 5.8% (4/69) of patient in the KB109 and SSC group reported hospitalization, emergency room visits, or urgent care visits compared with 15.2% (10/66) of patients treated with SSC alone, resulting in a 61.7% reduction in medically-attended visits. Similar results were seen in patients aged ≥45 years or with ≥1 comorbidity with medically-attended visits reduced by 52.8% in patients receiving KB109 plus SSC (5.6% [5/90 patients]) compared with SSC along (11.8% [10/85 patients]).

#### The natural history of COVID-19 infection in SSC-alone group in patients with and without comorbidities

When evaluating selected measures of health in outpatients with mild to moderate COVID-19, patients with ≥1 comorbidity had an extended duration of symptoms. The presence of ≥1 comorbidity extended the median time to symptom resolution in patients with mild to moderate COVID-19 who received SSC alone. Similar findings were reported for patients with no comorbidities at baseline and those with ≥1 comorbidity in both the 13 overall symptoms (21 vs 30 days, respectively; HR=1.163 [95% CI, 0.723-1.872]) (**Figure 3A)** and 8 cardinal symptoms (15 vs 21 days, respectively; HR=1.283 [95% CI, 0.809-2.035]) (**Figure 3B)**.

**Figure 3.**
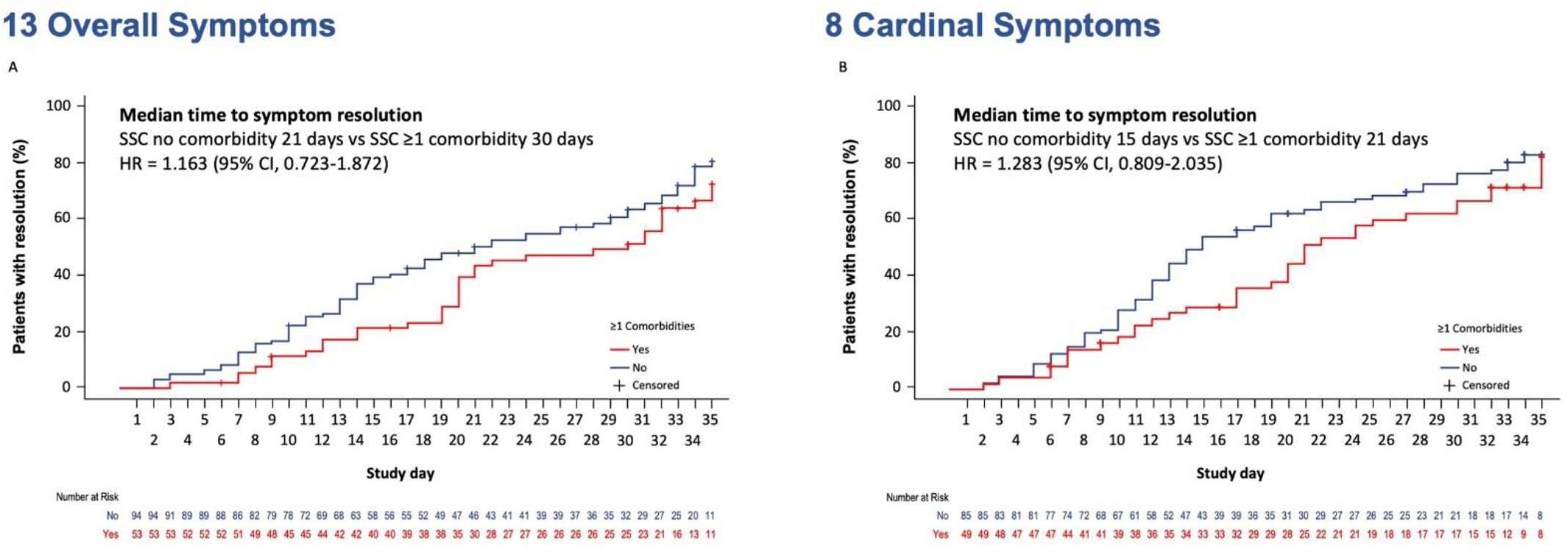
Natural history of COVID-19 infection: SSC-alone group median time to resolution of A) 13 overall symptoms and B) 8 cardinal symptoms.

#### Median time to resolution of the 13 overall and 8 cardinal symptoms by treatment group and ≥1 comorbidity

Overall, there was a modest difference in the time to resolution of the 13 overall symptoms with the administration of KB109 plus SSC vs SSC alone (HR=1.254 [95% CI 0.957-1.644]) as shown in **Figure 4A**. However, as shown in **Figure 4B**, the administration of KB109 plus SSC reduced median time to resolution of symptoms to 21 days compared with 30 days with SSC alone in patients reporting ≥1 comorbidity (HR=1.422 [95% CI, 0.898-2.250]). Similar results were seen with the analysis of the 8 cardinal symptoms (**Figure 5A**). Overall, there was a modest difference in the time to resolution of the 8 cardinal symptoms with the administration of KB109 plus SSC compared with SSC alone (HR=1.126 [95% CI 0.856-1.482]). However, in patients reporting ≥1 comorbidity, the addition of KB109 reduced the median time to resolution of symptoms to 17 days compared with 21 days in the SSC alone group (HR=1.574 [95% CI, 0.997-2.485]) (**Figure 5B**). Interestingly, the administration of KB109 plus SSC to patients with ≥1 comorbidity resulted in similar median times to resolution of symptoms as patients with no comorbidities receiving SSC alone (**Figure 6**).

**Figure 4.**
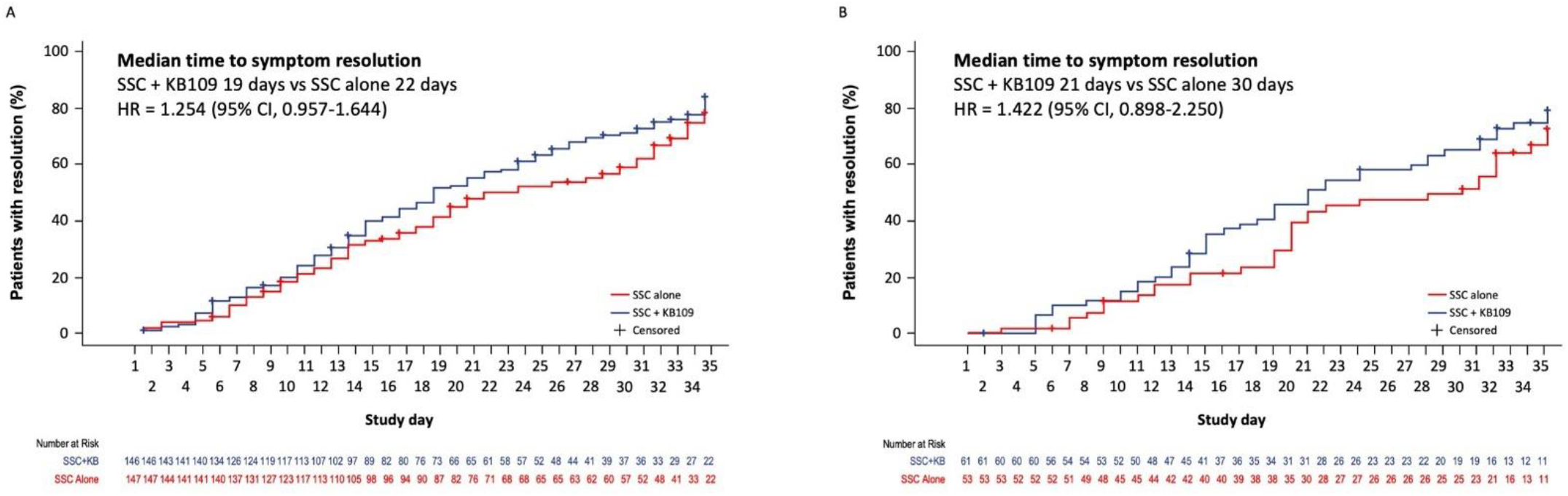
Median time to resolution of 13 overall symptoms in A) all patients and in B) patients with ≥1 comorbidity.

**Figure 5.**
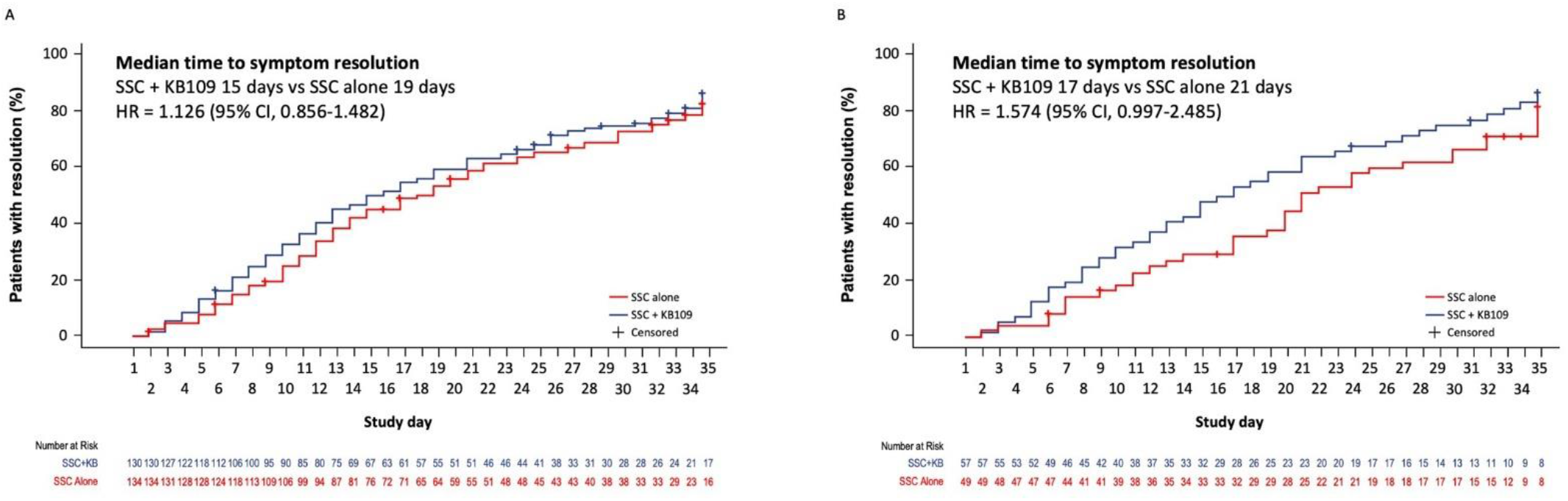
Time to resolution of 8 cardinal symptoms in A) all patients and in B) patients with ≥1 comorbidity.

**Figure 6.**
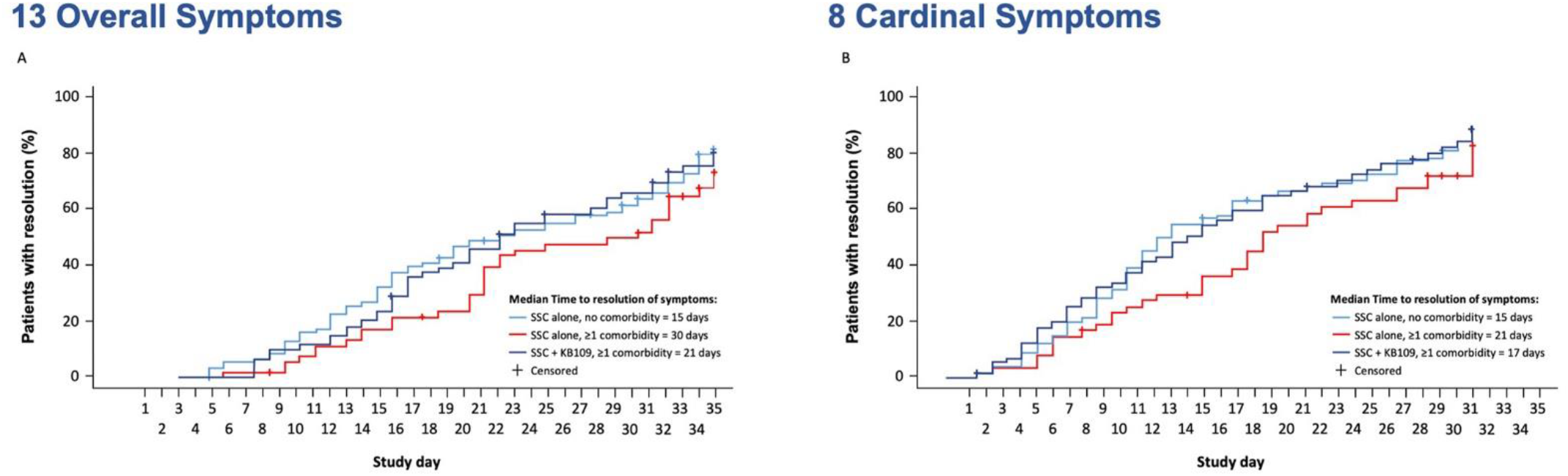
Median time to resolution of 13 overall and 8 cardinal symptoms by treatment group and comorbidity status.

#### Median time to resolution of symptoms in subgroup analyses

In subgroup analyses, administration of KB109 with SSC was associated with a reduction in the median time to resolution of the 13 overall symptoms in patients aged ≥45 to <65 years and in patients of Hispanic or Latino ethnicity (**Table 4**). Similar to the subgroup analyses for the 13 overall symptoms, administration of KB109 plus SSC was associated with a reduction in the median time to resolution of the 8 cardinal symptoms in patients aged ≥45 to <65 years (**Table 5**).

**Table 4.**
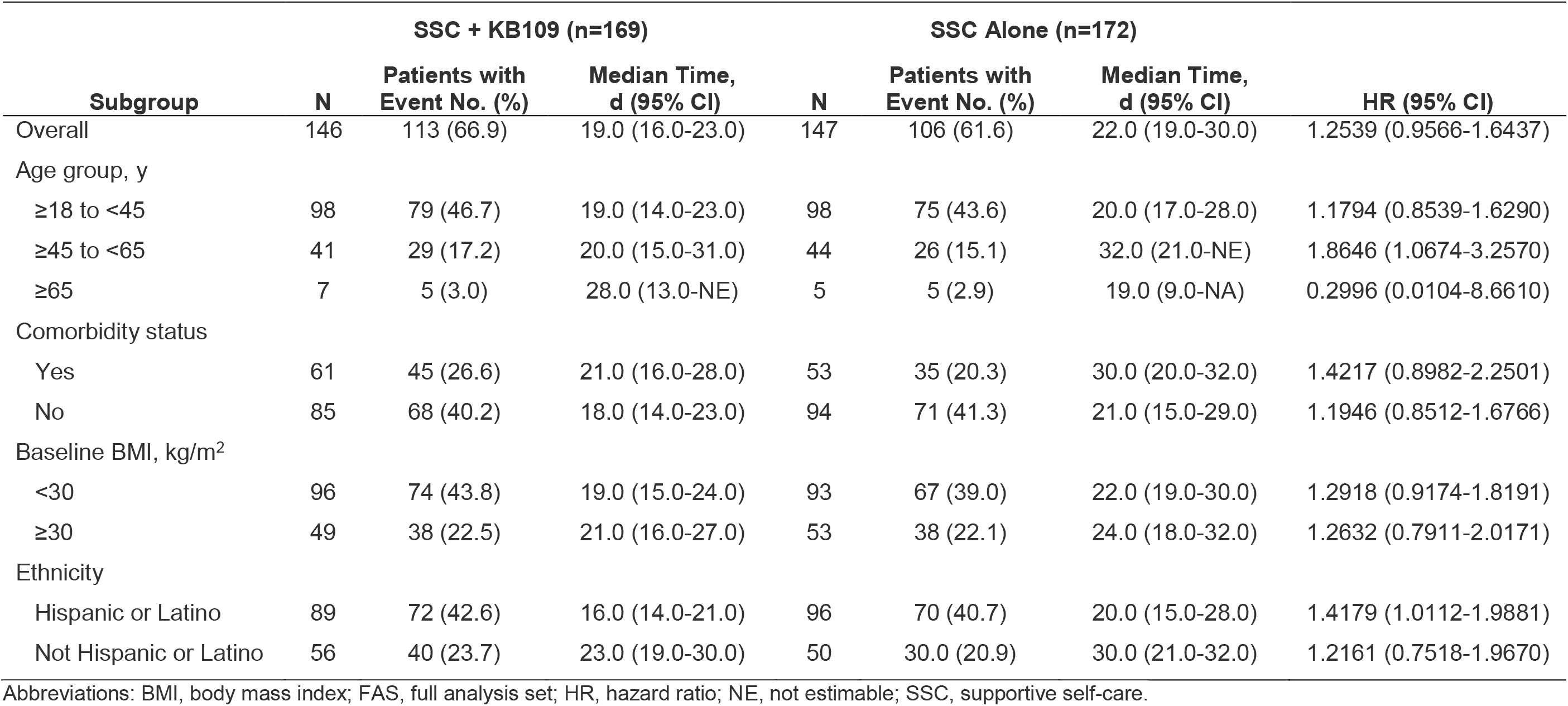
Subgroup analysis of time to resolution of 13 overall symptoms (FAS)

**Table 5.** Subgroup analyses of time to resolution of 8 overall symptoms (FAS)

| Subgroup | SSC + KB109 (n=169) |  |  | SSC Alone (n=172) |  |  | HR (95% CI) |
| --- | --- | --- | --- | --- | --- | --- | --- |
|  | N | Patients with Event No. (%) | Median Time, d (95% CI) | N | Patients with Event No. (%) | Median Time, d (95% CI) |  |
| Overall | 130 | 107 (63.3) | 15.0 (13.0-19.0) | 134 | 105 (61.0) | 19.0 (14.0-21.0) | 1.1262 (0.8558-1.4819) |
| Age group, y |  |  |  |  |  |  |  |
| ≥18 to <45 | 85 | 68 (40.2) | 16.0 (11.0-21.0) | 89 | 72 (41.9) | 15.0 (13.0-19.0) | 0.9764 (0.6978-1.3662) |
| ≥45 to <65 | 38 | 32 (18.9) | 15.0 (12.0-23.0) | 41 | 29 (16.9) | 24.0 (20.0-30.0) | 1.9645 (1.1301-3.4152) |
| ≥65 | 7 | 7 (4.1) | 21.0 (7.0-NE) | 4 | 4 (2.3) | 11.0 (9.0-NE) | 0.0526 (0.0031-0.8865) |
| Comorbidity status |  |  |  |  |  |  |  |
| Yes | 57 | 48 (28.4) | 17.0 (12.0-21.0) | 49 | 36 (20.9) | 21.0 (17.0-30.0) | 1.5743 (0.9974-2.4848) |
| No | 73 | 59 (34.9) | 14.0 (11.0-21.0) | 85 | 69 (40.1) | 15.0 (12.0-19.0) | 0.9623 (0.6742-1.3735) |
| Baseline BMI, kg/m<sup>2</sup> |  |  |  |  |  |  |  |
| <30 | 87 | 72 (42.6) | 15.0 (12.0-21.0) | 87 | 69 (40.1) | 17.0 (13.0-21.0) | 1.0754 (0.7634-1.5149) |
| ≥30 | 42 | 34 (20.1) | 16.5 (11.0-21.0) | 46 | 35 (20.3) | 20.0 (14.0-30.0) | 1.5441 (0.9293-2.5655) |
| Ethnicity |  |  |  |  |  |  |  |
| Hispanic or Latino | 81 | 69 (40.8) | 15.0 (10.0-18.0) | 85 | 68 (39.5) | 15.0 (12.0-20.0) | 1.1566 (0.8181-1.6353) |
| Not Hispanic or Latino | 48 | 37 (21.9) | 19.0 (12.0-25.0) | 48 | 37 (21.5) | 22.0 (17.0-30.0) | 1.1762 (0.7325-1.8888) |

Abbreviations: BMI, body mass index; FAS, full analysis set; HR, hazard ratio; NE, not estimable; SSC, supportive self-care.

When patients aged ≥45 years or with ≥1 comorbidity were evaluated together, the median time to resolution of the 13 overall symptoms was reduced with the administration of KB109 plus SSC vs SSC alone (21 vs 31 days, respectively; HR=1.597 [95% CI, 1.064-2.398]) (**Figure 7A**). Similarly, administration of KB109 plus SSC also reduced the median time to resolution of the 13 overall symptoms in patients aged <45 years and with ≥1 comorbidity compared with SSC alone (21 vs 31 days, respectively; HR=1.751 [95% CI, 0.836-3.665]) (**Figure 7B**), which suggests that the presence of ≥1 comorbidity may be responsible for the delayed time to resolution of symptoms between the 2 treatment groups.

**Figure 7.**
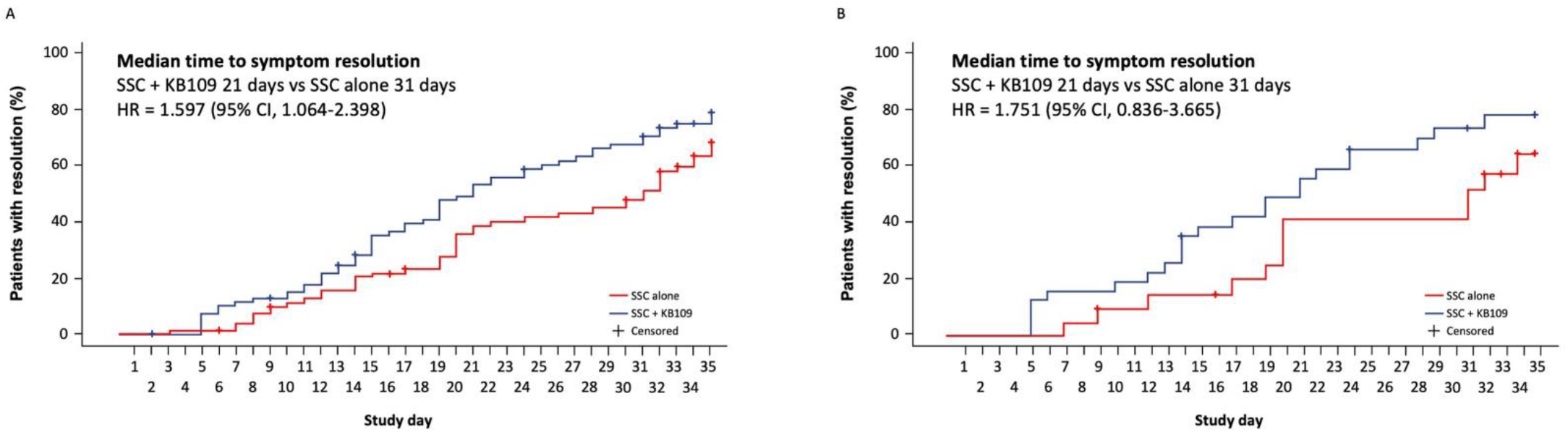
Median time to resolution of 13 overall symptoms in A) patients aged ≥45 years or with ≥1 comorbidity and B) in patients aged <45 year and with ≥1 comorbidity.

## Discussion

To our knowledge, this is one of the first studies to demonstrate that the presence of comorbidities prolongs the time to resolution of COVID-19–related symptoms. Throughout the study, the addition of KB109 to SSC was well tolerated with most TEAEs being GI-related and mild to moderate in severity. Patients receiving KB109 plus SSC had a reduction of 50.0% in medically-attended visits (ie, hospitalization, emergency room visits, or urgent care visits) compared with patients receiving SSC alone. These visits were reduced even further in higher risk populations—by 61.7% in patients with ≥1 comorbidity and by 52.8% in patients aged ≥45 years of age or ≥1 comorbidity. Additionally, results showed patients with ≥1 comorbidity had a longer duration of COVID-19–related symptoms compared with patients without comorbidities. The median time to resolution of the 13 overall symptoms associated with COVID-19 in patients with ≥1 comorbidity was reduced with the administration of KB109 plus SSC compared with SSC alone. This reduction was also evident when aged ≥45 years or ≥1 comorbidity was evaluated as a risk factor, risk factors also identified in other COVID-19 study patient populations.^23^

The COVID-19 pandemic is a global threat that affects individuals and entire healthcare systems alike.^1^ In 2021, BlueCross BlueShield analyzed over 90,000 COVID-19 cases and reported an average outpatient cost of $500 to $1000 per member; that cost was 45-times higher if a member was hospitalized.^24^ Patients identified with high-risk conditions (eg, diabetes, obesity, chronic obstructive pulmonary disease, chronic kidney disease, or heart disease) were 3-times more likely to be hospitalized in an intensive care setting with costs per admission that were 30% higher than for patients without these conditions.^24^ In our study, rates of medically-attended visits, including hospitalizations, were decreased with the administration of KB109 plus SSC compared with SSC alone. In higher-risk patients (eg, those with ≥1 comorbidity or aged ≥45 or ≥1 comorbidity), that rate was even further reduced. Although we did not measure healthcare cost associated with the medically-attended visits, a reduction in these visits, especially hospitalizations in higher-risk patients, has the potential to reduce not only healthcare utilization, but also healthcare costs. The impact of KB109 on healthcare costs should be evaluated in larger clinical trials in the future.

Despite a currently incomplete clinical picture of COVID-19, the broad disease spectrum encompasses asymptomatic infection, mild upper respiratory tract illness, severe respiratory failure, and death.^1^ To date, a paucity of data exists from randomized controlled trials that assess the natural history of disease progression and management of COVID-19 in an outpatient setting despite this group being the largest population of patients with COVID-19.^24^ Most trials have focused on improving patient outcomes for those hospitalized due to severe disease. Recently, a European study of patients with mild to moderate COVID-19 infection concluded that the clinical presentation varies according to the age and sex characteristics of patients (mean age, 39.17 +/- 12.09 years; 962 females, 458 males).^4^ Young patients were found to have ear, nose, and throat complaints, whereas elderly patients experienced fever, fatigue, loss of appetite, and diarrhea. Female patients had higher rates of loss of smell, headache, nasal obstruction, throat pain, and fatigue.^4^ Loss of smell persisted at least 7 days after end of disease in 37.5% of cured patients, and the mean duration of COVID-19 symptoms was 11.5±5.7 days.^4^ In our study, a protracted time to resolution of COVID-19–related symptoms, particularly in patients with comorbidities, was observed.

There was a modest difference in the time to resolution of symptoms with KB109 plus SSC compared with SSC alone. However, in patients receiving KB109 plus SSC with ≥1 comorbidity, median time to resolution of the 13 overall symptoms was reduced by 9 days; median time of resolution of the 8 cardinal symptoms was reduced by 5 days. When patients aged ≥45 years or patients with ≥1 comorbidity were evaluated together, the addition of KB109 reduced the median time to resolution of the 13 overall symptoms to 21 days compared with 31 days in SSC alone. In patients who were <45 years of age and had ≥1 comorbidity, approximately 40% of patients receiving SSC alone continued to experience COVID-19–related symptoms at day 35 compared with approximately 20% of patients receiving KB109 in addition to SSC.

When developing the conditions of use for casirivimab and imdevimab, a monoclonal antibody cocktail used in the treatment of COVID-19, the European Medicines Agency defined target populations as patients aged 12 years and older who do not require supplemental oxygen for COVID-19 and who are at high risk of progressing to severe COVID-19.^23^ Patient comorbidities placing them at high risk for progression included advanced age, obesity, cardiovascular disease (including hypertension), chronic lung disease (including asthma), type 1 or 2 diabetes mellitus, chronic kidney disease (including those on dialysis), chronic liver disease, and immunosuppression (including cancer treatment). Interestingly, hypertension was reported as the highest comorbidity in this study. In previous studies, hypertension in patients with COVID-19 has been associated with a greater risk of developing acute respiratory distress syndrome or requiring ICU care.^25,26^ Therefore, exploration of the impact of comorbidities and concomitant medications, including anti-hypertensives (eg, angiotensin-converting enzyme inhibitors, angiotensin receptor blockers), on time to resolution of COVID-19–related symptoms when combined with KB109 plus SSC could be a consideration in the future.

Although much of the focus of COVID-19 treatment has been focused on viral eradication, modulation of the inflammatory response to the virus is less understood and has been harder to control via treatment with therapeutics. Although a complete understanding of the modulation of respiratory virus infectivity and immune response by the commensal microbiota is still lacking, we sought to evaluate influencing the microbiome to determine its effect on safety and the natural progression of the disease in patients with mild-to-moderate COVID-19 infection. There are data supporting a key role for both gut-derived metabolites (eg, SCFAs) and the direct interaction and migration of immune cells from gut to lung by the common mucosal immune system in pulmonary infections.^19^ For example, an *ex vivo* assay has been used to induce and measure the fermentation of different oligosaccharide ensembles, including KB109, by human fecal samples.^22^ In addition, *ex vivo* assay testing has shown KB109 increased the amount of SCFAs produced over a negative control by 4-to 6-fold across multiple fecal communities.^22^ Finally, direct activation of host immune cells and pathways by microbiota in the gut has been shown to affect the progression of pulmonary infections.^19^ This reveals the importance of commensal microbiota in regulating immunity in the respiratory mucosa through the proper activation of inflammasomes. KB109 is the first synthetic glycan to demonstrate an effect on the host immune response rather than targeting a specific virus. This is of clinical importance in light of the number of emerging SARS-CoV-2 variants. On March 24, 2021, bamlanivimab monotherapy was halted in the US due to the resistance of emerging SARS-CoV-2 variants.^27^ Given the influence KB109 has on the gut microbiome and its role in host immune cells, this synthetic glycan has the potential to retain activity despite emerging variants as its impact is on the immune system, not against a specific strain of SARS-CoV-2.

In this study, by executing a novel, virtual clinical trial design, we successfully examined the natural history of COVID-19 disease progression and the influences of modulating the microbiome in patients with mild to moderate symptoms of COVID-19 in the outpatient setting. Although more patients receiving KB109 and SSC experienced ≥1 TEAE (36.1% [61 of169] of patients) than SSC alone (26.5% [48 of 181] of patients), we found KB109 to be safe and well tolerated. In addition, all GI-related AEs were mild to moderate—with one leading to discontinuation. Furthermore, we showed that patients with ≥1 comorbidity treated with KB109 plus SSC reported a time to resolution in both the 13 overall symptoms and 8 cardinal symptoms similar to patients without comorbidities treated with SSC alone. Based on our results, KB109, as on oral agent, should be considered in the early care of non-hospitalized patients with mild to moderate COVID-19 infections to potentially decrease time to resolution of symptoms and medically attended visits.

## Study Limitations

Limitations of this study included an open-label design, lack of a placebo control, a relatively small sample size, and a lack of racial diversity, with 90% of patients enrolled in study identifying as White. However, a strength is that over half of the population was of Hispanic or Latino ethnicity, a population that has been disproportionately affected by the pandemic and underrepresented in other COVID-19 studies.^28–30^ In addition, remote data collection in itself has limitations, including patients self-reporting comorbidities and clinical signs (eg, temperature, pulse oximeter readings), and imposed limitations on study eligibility which required access to appropriate technology (eg, smartphone/internet access).^31^

## Conclusion

This study provides a better understanding of the natural disease progression and management of COVID-19 in patients with mild to moderate disease in an outpatient setting and shows that the presence of comorbidities prolongs time to resolution of COVID-19–related symptoms. The administration of KB-109, a novel, synthetic glycan, with SSC was well tolerated with the majority of TEAEs being GI in nature and mild to moderate in severity. The promising results of this study suggest that administration of KB109 plus SSC may be associated with reduced times to resolution of COVID-19 symptoms and decreased rates of medically-attended visits in non-hospitalized patients with mild to moderate COVID-19, especially in those with ≥1 comorbidity and those that are at higher risk for COVID-19–related complications. Future evaluation of KB109 in larger studies is warranted to replicate these results.

## Supporting information

CONSORT Checklist

## Data Availability

Qualified scientific and medical researchers may make requests for individual participant data that underlie the results (text, tables, figures, and supplement) reported in this article, after de-identification, at. Methodologically sound proposals for such data will be evaluated and approved by Kaleido Biosciences, Inc, in its sole discretion. All approved researchers must sign a data access agreement prior to accessing the data. Data will be available as soon as possible but no later than within 1 year of the acceptance of the article for publication, and for 3 years following article publication. Kaleido Biosciences, Inc, will not share identified participant data or a data dictionary.

## Acknowledgments

We thank the patients, the investigators, and the investigational staff of the K031 study and the management and execution of the study by Dawn McCollough and Silvia Helou, MD, of Kaleido Biosciences, Inc. The authors acknowledge the medical writing assistance of Laura Jung, PharmD and Larry Wright of PRECISIONscientia in Yardley, PA, USA, which was supported financially by Kaleido Biosciences, Inc, in compliance with international Good Publication Practice guidelines.

## References

1. Machhi J, Herskovitz J, Senan AM, et al. The natural history, pathobiology, and clinical manifestations of SARS-CoV-2 infections. J Neuroimmune Pharmacol. 2020;15(3):359–386.

2. Johns Hopkins. Global map. Updated February 9, 2021. Accessed March 19, 2021. https://coronavirus.jhu.edu/map.html.

3. CDC website. New variants of the virus that causes COVID-19. Updated February 2, 2021. Accessed February 10, 2021. https://www.cdc.gov/coronavirus/2019-ncov/transmission/variant.html.

4. Lechien JR, Chiesa-Estomba CM, Place S, et al. Clinical and epidemiological characteristics of 1420 European patients with mild-to-moderate coronavirus disease 2019. J Intern Med. 2020;288(3):335–344.

5. CDC website. Symptoms of coronavirus. Updated December 22, 2020. Accessed February 10, 2021. https://www.cdc.gov/coronavirus/2019-ncov/symptoms-testing/symptoms.html.

6. Xie P, Ma W, Tang H, Liu D. Severe COVID-19: a review of recent progress with a look toward the future. Front Public Health. 2020;8:189.

7. Cohen PA, Hall LE, John JN, Rapoport AB. The early natural history of SARS-CoV-2 infection: clinical observations from an urban, ambulatory COVID-19 clinic. Mayo Clin Proc. 2020;95(6):1124–1126.

8. Huang C, Huang L, Wang Y, et al. 6-month consequences of COVID-19 in patients discharged from hospital: a cohort study. The Lancet. 2021;397(10270):220–232.

9. CDC website. People with certain medical conditions. Updated February 3, 2021. Accessed February 10, 2021. https://www.cdc.gov/coronavirus/2019-ncov/need-extra-precautions/people-with-medical-conditions.html

10. Sanyaolu A, Okorie C, Marinkovic A, et al. Comorbidity and its impact on patients with COVID-19. SN Compr Clin Med. 2020:1–8.

11. Belkaid Y, Harrison OJ. Homeostatic immunity and the microbiota. Immunity. 2017;46(4):562–576.

12. Haak BW, Littmann ER, Chaubard JL, et al. Impact of gut colonization with butyrate-producing microbiota on respiratory viral infection following allo-HCT. Blood. 2018;131(26):2978–2986.

13. Trompette A, Gollwitzer ES, Pattaroni C, et al. Dietary fiber confers protection against flu by shaping Ly6c(-) patrolling monocyte hematopoiesis and CD8(+) T cell metabolism. Immunity. 2018;48(5):992–1005 e1008.

14. Dhar D, Mohanty A. Gut microbiota and Covid-19-possible link and implications. Virus Res. 2020;285:198018.

15. Chiu L, Bazin T, Truchetet ME, Schaeverbeke T, Delhaes L, Pradeu T. Protective microbiota: from localized to long-reaching co-immunity. Front Immunol. 2017;8:1678.

16. Keely S, Talley NJ, Hansbro PM. Pulmonary-intestinal cross-talk in mucosal inflammatory disease. Mucosal Immunol. 2012;5(1):7–18.

17. Yeoh YK, Zuo T, Lui GC, et al. Gut microbiota composition reflects disease severity and dysfunctional immune responses in patients with COVID-19. Gut. 2021;70(4):698–706.

18. Gu S, Chen Y, Wu Z, et al. Alterations of the Gut Microbiota in Patients With Coronavirus Disease 2019 or H1N1 Influenza. Clin Infect Dis. 2020;71(10):2669–2678.

19. Ichinohe T, Pang IK, Kumamoto Y, et al. Microbiota regulates immune defense against respiratory tract influenza A virus infection. Proc Natl Acad Sci U S A. 2011;108(13):5354–5359.

20. Gou W, Fu Y, Yue L, et al. Gut microbiota may underlie the predisposition of healthy individuals to COVID-19. medRxiv. doi:10.1101/2020.04.22.20076091.

21. Parada Venegas D, De la Fuente MK, Landskron G, et al. Short Chain Fatty Acids (SCFAs)-Mediated Gut Epithelial and Immune Regulation and Its Relevance for Inflammatory Bowel Diseases. Front Immunol. 2019;10:277.

22. Meisner J, Lawrence J, Lee J, Roed M, van Hylckama Vlieg Jet. Development of a novel synthetic glycan to prevent bacterial infections and meliorate respiratory viral infections. Presented at: IDWeek Interactive Program. October 22–25, 2020.

23. European Medicines Agency. REGN-COV2 antibody combination (casirivimab/imdevimab) - COVID19 - Article-5(3) procedure: Conditions of use, conditions for distribution and patients targeted conditions for safety monitoring. First published February 26, 2021. Accessed March 24, 2021. https://www.ema.europa.eu/en/news/ema-issues-advice-use-regn-cov2-antibody-combination-casirivimab-imdevimab.

24. BlueCross BlueShield. Infographic: COVID-19 patients with high-risk conditions 3x more likely to need the ICU. Published February 9, 2021. Accessed March 26, 2021. https://www.bcbs.com/coronavirus-updates/stories/infographic-covid-19-patients-high-risk-conditions-3x-more-likely-need-the-icu.

25. Wu C, Chen X, Cai Y, et al. Risk factors associated with acute respiratory distress syndrome and death in patients with coronavirus disease 2019 pneumonia in Wuhan, China. JAMA Intern Med. 2020;180(7):934–943.

26. Wang D, Hu B, Hu C, et al. Clinical characteristics of 138 hospitalized patients with 2019 novel coronavirus-infected pneumonia in Wuhan, China. JAMA. 2020;323(11):1061–1069.

27. US Department of Health & Human Services. Public Health Emergency. Bamlanivimab: update on COVID-19 variants and impact on bamlanivimab distribution. Published March 24, 2021. Accessed March 26, 2021. https://www.phe.gov/emergency/events/COVID19/investigation-MCM/Bamlanivimab/Pages/default.aspx.

28. Podewils LJ, Burket TL, Mettenbrink C, et al. Disproportionate Incidence of COVID-19 Infection, Hospitalizations, and Deaths Among Persons Identifying as Hispanic or Latino - Denver, Colorado March-October 2020. MMWR Morb Mortal Wkly Rep. 2020;69(48):1812–1816.

29. Chastain DB, Osae SP, Henao-Martínez AF, Franco-Paredes C, Chastain JS, Young HN. Racial disproportionality in Covid clinical trials. N Engl J Med. 2020;383(9):e59.

30. Calo WA, Murray A, Francis E, Bermudez M, J. K. Reaching the hispanic community about COVID-19 through existing chronic disease prevention programs. Prev Chronic Dis. 17:200165. DOI: http://dx.doi.org/200110.205888/pcd200117.200165.

31. Ali Z, Zibert JR, Thomsen SF. Virtual clinical trials: perspectives in dermatology. Dermatology. 2020;236(4):375–382.

