## Supplement 2 for "Targeting the Microbiome With KB109 in Outpatients with Mild to Moderate COVID-19 Reduced Medically Attended Acute Care Visits and Improved Symptom Duration in Patients With Comorbidities"

**Supplemental Methods**

*Eligibility criteria*

Patients with concurrent use of these medications during the study were excluded: immunomodulatory agent within 12 months of study screening; systemic antibiotics, antifungals, or antivirals for treatment of active infection within 28 days of study screening; systemic immunosuppressive therapy within 3 months of study screening; and drugs or other compounds that modulate gastrointestinal motility (including stool softeners, laxatives, or fiber supplements) taken currently or within 7 days of Study screening. Antacid (histamine 2 blockers and proton pump inhibitors) and antidiarrheal agents were not prohibited.

*Healthcare utilization*

From day 2 onwards, patient-assessed measured of healthcare utilization were performed every morning upon awakening until the end of the study. Measures included:

1. Emergency room or urgent care visit: rated as yes or no; if yes, reason was recorded
2. Medical provider visit: rated as yes or no; if yes, then reason as well as if this was as visit for a co-existing condition was recorded
3. Hospital stay: rated as yes or no; if yes, reason was recorded
