## Supplement 1 for "Targeting the Microbiome With KB109 in Outpatients with Mild to Moderate COVID-19 Reduced Medically Attended Acute Care Visits and Improved Symptom Duration in Patients With Comorbidities"

### A Randomized, Open Label, Prospective, Parallel Group Study to Assess the Natural History of COVID-19 and Effects of KB109 in Addition to Supportive Self Care (SSC) Compared to SSC Alone on Measures of Health in Non-hospitalized Patients with Mild-to-Moderate COVID-19

**Protocol #:** K031-120

**Sponsor:** Kaleido Biosciences, Inc.
65 Hayden Ave
Lexington, MA 02421, USA

**Version #:** 4

**Version Date:** 09 Dec 2020

This document is a confidential communication of Kaleido Biosciences, Inc. (Kaleido). The recipient agrees that no information contained herein will be published or disclosed without Kaleido's prior written approval, except that this document may be disclosed to appropriate institutional review boards or duly authorized representatives of the appropriate regulatory agencies under the condition they are requested to keep it confidential.

### SIGNATURE OF PRINCIPAL INVESTIGATOR

**Study Title**: A Randomized, Open Label, Prospective, Parallel Group Study to Assess the Natural History of COVID-19 and Effects of KB109 in Addition to Supportive Self Care (SSC) Compared to SSC Alone on Measures of Health in Non-hospitalized Patients with Mild-to-Moderate COVID-19

**Protocol Number**: K031-120

This protocol is sponsored by Kaleido Biosciences, Inc. (Kaleido). All the data and any other information generated from the execution of this protocol are the sole property of Kaleido and may not be used for any purpose without written permission from Kaleido.

The signature below constitutes the approval of this protocol and provides the necessary assurance that this trial will be conducted according to all stipulations of the protocol, including all statements regarding confidentiality, and according to applicable US Federal regulations, and ICH/GCP guidelines.

Principal Investigator's Signature: ________________________________

Principal Investigator's Name: ________________________________

Institution Name: ________________________________

Date: ________________________________

### SPONSOR APPROVAL PAGE

**Study Title**: A Randomized, Open Label, Prospective, Parallel Group Study to Assess the Natural History of COVID-19 and Effects of KB109 in Addition to Supportive Self Care (SSC) Compared to SSC Alone on Measures of Health in Non-hospitalized Patients with Mild-to-Moderate COVID-19

Version 4.0, 09 Dec 2020

Person authorized to sign the protocol and protocol amendment(s) for the Sponsor, Kaleido:

Sponsor Signature:

Mark Wingertzahn, PhD
VP Research and Development and Head of Clinical Development
Kaleido Biosciences, Inc.

Date: 9-DEC-2020

### 1 PROTOCOL SYNOPSIS

| |
|---|
| **Name of Sponsor/Company:**<br>Kaleido Biosciences, Inc. |
| **Name of Finished Product:**<br>Study product (SP) KB109 (spray-dried, synthetic oligosaccharide food composition) |
| **Name of Substance Investigated:**<br>KB109 |
| **Title of Study:**<br>A Randomized, Open Label, Prospective, Parallel Group Study to Assess the Natural History of COVID-19 and Effects of KB109 in Addition to Supportive Self Care (SSC) Compared to SSC Alone on Measures of Health in Non-hospitalized Patients with Mild-to-Moderate COVID-19 |
| **Study Rationale:**<br>The COVID-19 pandemic threatens individuals, societies and healthcare systems around the world. Host immunity contributes to the susceptibility as well as the progression of infectious disease and patient outcomes. Clinical studies and *ex vivo* experiments using human fecal samples have demonstrated that commensal bacteria selectively ferment proprietary oligosaccharide mixtures to promote their growth and produce metabolites such as the short-chain fatty acids (SCFAs) including acetic, propionic, and butyric acid. SCFAs modulate host inflammation, control adaptive immunity, and promote immune tolerance in the gut as well as systemically (Thaiss 2016; Schirmer 2016). In particular SCFAs and SCFA-producing taxa have been linked to reduced risk of acquiring viral infections (including corona-viral infections) in at-risk populations (e.g., Hematopoietic stem cell transplantation [HSCT] patients, Haak 2018). Studies in mice have demonstrated that SCFAs and SCFA-producing microbial taxa induce virus-specific CD4+ and CD8+ T cells, interferon type 1, and antibody responses involved in the reduction of viral infection severity (Ichinohe 2011; Trompette 2018). Data has also shown that protection against Respiratory Syncytial Virus (RSV) infection can be conferred by acetate, a metabolite produced by the intestinal microbiota, through induction of interferon (IFN)-β in the lung through GPR43 and IFNAR-mediated pathways (Antunes 2019). In addition, SCFAs have also been reported to influence macrophage functionality to mitigate neutrophil-mediated tissue damage (Trompette 2018). This is of particular importance as viral infections may be accompanied by an aggressive pro-inflammatory response (Haak 2018) that can elicit a syndrome known as “cytokine storm”.<br><br>Kaleido’s orally administered proprietary oligosaccharide mixtures are related to substances found in the human diet that modulate the composition and metabolic output of the microbiome and are generally recognized as safe (GRAS). They allow the controlled production of SCFAs with respect to absolute quantities as well as in relative abundance. Capitalizing on this property, Kaleido has selected a proprietary oligosaccharide mixture, termed KB109**,** that holds promise in inducing a beneficial profile of SCFA production in the gut while driving the growth of commensal bacteria. Increased SCFA and the modulation in microbiome taxonomy may lead to a more appropriate immune and inflammatory response and may prevent an over-aggressive response, up to and including “cytokine storm” observed in patients with COVID-19.<br><br>KB109 is a chemically synthesized oligosaccharide mixture resulting from the polymerisation of glucose:galactose:mannose that has been studied in humans and has been determined to be GRAS for its intended use. It is intended for oral administration and is supplied as a powder to be reconstituted in water. In an *ex vivo* model, KB109 increased the amount of SCFAs produced over water control by approximately three-fold across multiple healthy fecal communities. In addition, it is currently being assessed in an ongoing clinical food study in patients colonized with multidrug-resistant (MDR) organisms to reduce the relative abundance of associated taxa including *Enterobacteriaceae*, *Enterococcus*, and *Clostridium difficile* compared to observational controls (NCT 03944369).<br><br>In this study, outpatients with mild-to-moderate COVID-19 illness in the presence and absence of KB109 will be evaluated for its safety and physiologic effects. Physiologic effects will be assessed by measuring biomarkers of inflammation and antibodies as well as physiologic responses of health in patients, quality of life (QOL), and healthcare utilization. |
| **Study Period:** Up to 47 days (including the Screening period through end of assessments) |

| Objective: | Endpoint: |
|---|---|
| **Primary:** To evaluate the safety of KB109 in addition to Supportive Self Care (SSC + KB109) compared to SSC alone in outpatients with mild-to-moderate COVID-19 | • Number of patients experiencing SP-related treatment-emergent adverse events (TEAEs) |
| **Secondary:** To evaluate selected measures of health in outpatients with mild-to-moderate COVID-19 | • Time to resolution of overall 13 COVID-19 related symptoms which is defined as from Day 1 until the day at which the overall composite score of 13 COVID-19 related symptoms becomes 0 or 1 and remains at 0 or 1 for the rest of the Intake Period and for the Follow-up Period. Overall composite score of 13 COVID-19 related symptoms is the sum of 13 COVID-19 related symptom scores (i.e., cough, chills/repeated shaking with chills, muscle pain, fever, headache, anosmia/ageusia, shortness of breath, sore throat, gastrointestinal disturbance/symptoms, diarrhea, fatigue, nasal congestion, and chest tightness (CDC 2020). Each COVID-19 symptom will be recorded by patients on a scale of 0: Absent, 1: Mild, 2: Moderately severe, 3: Very severe. The overall composite score ranges from 0 (no symptoms) to 39 (very severe)<br>• Time to resolution of overall 8 cardinal COVID-19 related symptoms which is defined as from Day 1 until the day at which the overall composite score of 8 cardinal COVID-19 related symptoms becomes 0 or 1 and remains at 0 or1 for the rest of the Intake Period and for the Follow-up Period. Overall composite score of 8 cardinal COVID-19 related symptoms is the sum of 8 cardinal COVID-19 related symptom scores (i.e., cough, chills/repeated shaking with chills, muscle pain, fever, headache, anosmia/ageusia, shortness of breath, and sore throat)<br>• Proportion of patients with reduction from Baseline (symptom present at Baseline) in each of 13 individual COVID-19 related symptom at End of Intake Period (EOI) and Follow-up<br>• Proportion of patients with symptom becomes absent (symptom present at Baseline) at EOI and Follow-up for each of 13 individual COVID-19 related symptom<br>• Change from Baseline to EOI in overall composite score of 13 COVID-19 related symptoms<br>• Change from Baseline to EOI in overall composite score of 8 cardinal COVID-19 related symptoms<br>• Time to resolution of fever (defined as from Day 1 until the day at which a patient's daily maximum temperature achieves and remains below 100.4 °F for the rest of the Intake Period and for the Follow-up Period without an antipyretic medication)<br>• Proportion of patients with oxygen saturation <95% on Day 14 and Day 35<br>• Proportion of patients with oxygen saturation <98% on Day 14 and Day 35 |

| | |
|---|---|
| | • Measures collected from the Healthcare Provider Wellness Visits<br>• Proportion of patients experiencing hospital admissions during the Intake Period and Follow-up Period (all cause, and COVID-19-related)<br>• Healthcare Utilizations during the Intake Period and Follow-up Period |
| **Exploratory:** To evaluate measures of health in outpatients with mild-to-moderate COVID-19 during the Follow-up Period | • Change from Baseline to the EOI in individual COVID-19 related symptom score: cough, chills/repeated shaking with chills, muscle pain, fever, headache, anosmia/ageusia, shortness of breath, sore throat, gastrointestinal disturbance/symptoms, diarrhea, fatigue, nasal congestion, and chest tightness<br>• Individual measures of QOL<br>• Change from Baseline to EOI in bedrest time measured as patient-assessed daily cumulative total rest (measured in hours)<br>• Proportion of patients with increases in patient global impression on COVID-19 condition (PGIC)<br>• Proportion of patients with temperature below 100.4 °F without an anti-pyretic medication |
| **Exploratory:** To evaluate the changes in laboratory measures, specific biomarkers, serology and viral load in outpatients with mild-to-moderate COVID-19. | • Change from Baseline (Day 1) in:<br> o Laboratory measures<br> o Biomarkers of infection, antibody response, and inflammation (e.g., D-dimer, lipocalin, cytokines, IgM/IgG sero-conversion, and Neutralization Assays) |

**Methodology:**

This is a randomized, controlled, multi-site, open label clinical food study. It is intended to assess KB109 on safety as well as measures of signs, symptoms, healthcare utilization (including hospitalizations), laboratory/biochemical indices, and QOL measures in outpatients who have tested positive with COVID-19, have mild-to-moderate disease, and have been advised to manage their disease at home with SSC under quarantine protocols set forth by the CDC or local ordinances or practices as advised by their healthcare provider.

Patients who attend an outpatient clinic for suspected COVID-19 will be recruited for this study. All patients who enter this study will continue to follow the supportive self-care (SSC) guidance as instructed by the treating healthcare provider at the outpatient clinic. Note that during this outpatient study, the interaction between study staff and patients will be minimized to limit risk of infection spread. Patients will be recording responses to study-related questions in a secure website called "TrialPace™ diary" (TrialPace™) and may have healthcare professionals visit their home for study procedures and assessments as outlined in the Schedule of Assessments (SOA).

This study comprises 2 parts.

**Part 1:** The SARS-CoV-2 diagnostic testing (RT-PCR test, antigen test or equivalent test to detect active infection) will be performed. Hospital, academic, or industry-based assays will be acceptable for the diagnosis of COVID-19; home-based tests are not accepted for this study. After informed consent, patients will be asked to record COVID-19 symptoms in TrialPace™ using a unique log-in and password; this may occur prior to or after having received the result of the SARS-CoV-2 test. Potential study patients may have received a test for COVID-19 at a different clinic than the study site location and may present to the study site for consideration for study enrollment. If feasible blood samples for hematology, chemistry, biomarkers and serological measures of immunity, nasal/oropharyngeal swabs for quantitative vial load assessments (research purposes only) will be collected. Patients who are symptomatic at the time of getting a positive COVID-19 test result and meet eligibility criteria will enter Part 2 of the study within 48 hours of testing positive. Patients who are pre-symptomatic at the time of testing, develop symptoms within 7 days of testing positive and meet eligibility criteria will enter Part 2 of the study within 5 days of the symptoms showing.

**Part 2:** Part 2 is comprised of an Intake Period (Days 1-14) and a Follow-up Period (Days 15-35):

Intake Period (Days 1-14): Eligible patients will be randomized (1:1) to receive either SSC + KB109 or remain on SSC alone. The randomization will be stratified by age sub-group and presence or absence of any co-morbidities. Upon confirmation of study eligibility, patients will be provided the SP (KB109, if assigned to that group) and the Kaleido At-home Study Kit (KaSK) that will contain the SP dosing instructions (as applicable), a thermometer, a pulse oximeter, and telemedicine contact information.

During the Intake Period, patients assigned to SSC+KB109 will self-administer KB109 (twice daily) starting on the morning after receipt of the KaSK according to the schema outlined below in addition to maintaining a stable diet.

Patients will consume KB109 orally after reconstitution in water, twice daily, at least 8 hours apart, according to the following dosing schedule:

| **SP Dose Administration Period** | **KB109** |
|---|---|
| Days 1–2 | 18 g total daily dose consumed in two divided doses (9 g each dose) |
| Days 3-4 | 36 g total daily dose consumed in two divided doses (18 g each dose) |
| Days 5-14 | 72 g total daily dose consumed in two divided doses (36 g each dose) |

During the Intake Period, patients in both groups (SSC+KB109 or SSC alone) will continue to follow the SSC guidance as instructed by the treating healthcare provider. Patients will also record signs and symptoms, and responses to QOL and healthcare utilization questions using TrialPace™ for 14 days. For patients in the SSC+KB109 group, Day 1 measures will be recorded prior to the first dose of KB109. For all patients in both groups, temperature and oxygen saturation will be measured and recorded as needed throughout the day prior to taking anti-pyretic medication.

Patients will have wellness visits by telephone on Days 3, 7, 10, and 14 where compliance, safety, and COVID-19 health status will be monitored. If the wellness visit falls on the same day as a telemedicine visit, then the wellness visit assessments will be captured during the telemedicine visit (i.e., there will be no separate wellness visit on Day 14). On the last day of the Intake Period (Day 14), patients will have a follow-up telemedicine visit to assess safety, and if feasible, blood samples for clinical chemistries, biomarkers and serological measures of immunity, and nasal and oropharyngeal swabs for quantitative viral load assessments (research purposes only), will be taken as outlined in the SOA; sample collection may occur at the patient's home by a healthcare professional, remotely, or at the study site or clinic.

Follow-up Period (Days 15–35): During the Follow-up Period, patients will continue to report on signs and symptoms, QOL, and healthcare utilization. Patients will have wellness visits by telephone on Days 21 and 28 where safety and COVID-19 health status will be monitored. On Day 35, patients will have a follow-up telemedicine visit to assess safety, and if feasible blood samples for clinical chemistries, biomarkers and serological measures of immunity, and nasal and oropharyngeal swabs for quantitative viral load assessments (research purposes only) will be taken as outlined in the SOA; sample collection may occur at the patient's home by a healthcare professional, remotely, or at the study site or clinic. Wellness visit assessments will be captured during the telemedicine visit on Day 35 (i.e., there will be no separate wellness visit on Day 35).

Throughout the study, safety will be monitored by adverse events (AEs). COVID-19 related symptoms of the disease under study will not be classified as TEAEs as long as they are within the normal day-to-day fluctuation or expected progression of the disease not including hospitalizations and are part of the clinical data of the disease that is being collected in the study. Any patient experiencing a TEAE, significant worsening of COVID-19 symptoms or intolerable GI symptoms will be evaluated by the principal investigator (PI) or designee via a telemedicine visit and referred as needed for emergent follow-up, or in the case of intolerable GI symptoms for patients in the SSC+KB109 group, for interruption and/or down-titration of SP dose to the previously tolerated level.

In addition to the final analysis at the completion of the study, an interim analysis is planned when approximately 40% of the total randomized patients have completed or discontinued prior to completion of the EOI period.

**Number of patients planned**: Approximately 350-400 (175-200 patients per group).

**Eligibility Criteria:**

To be considered for enrollment into this study, each patient must meet **all** of the following Inclusion Criteria:

1. Be male or female, ≥18 years of age
2. Be willing and able to give informed consent, provide medical history, and secondary contact information
3. If symptomatic at the time of COVID-19 testing, the extended symptom(s), 8 cardinal plus 5 additional symptom(s) (see Section 4.3.4.1 for reference) must be new or worsening at baseline and must not have been present (see Section 4.3.4.1 for reference) for more than 5 days. Symptomatic patients must be screened and randomized within 48 hours of a positive test.
4. If pre-symptomatic at time of COVID-19 testing, new cardinal symptoms (see Section 4.3.4.1 for reference) must be reported within 7 days of a positive test, and the patient must be screened and randomized within 5 days of them developing symptoms
5. Mild to moderate COVID-19 and self-reported outpatient management indicated by their healthcare provider
6. Able to adhere to the study visit schedule and other protocol requirements
7. Has consistent internet or cellphone access with a data plan and access to a smartphone, tablet or computer

Patients who meet **any** of the following Exclusion Criteria at the Randomization Visit will not be enrolled into the study:

1. In the Primary Investigator's judgement, patients likely to require hospitalization for COVID-19
2. Patients who are hospitalized for in-patient treatment or currently being evaluated for potential hospitalization at the time of informed consent for conditions other than COVID-19
3. History of chronic lung disease with chronic hypoxia
4. History of documented cirrhosis or end-stage liver disease
5. Ongoing requirement for oxygen therapy
6. Shortness of breath in resting position
7. Diagnosis of sleep apnea requiring Bilevel Positive Airway Pressure (BIPAP) / Continuous Positive Airway Pressure (CPAP)
8. Female patients who are pregnant, trying to become pregnant or lactating.
9. Concurrent use of any of the following medications:
    a. Therapy with an immunomodulatory agent within 12 months of study screening
    b. Systemic antibiotics, antifungals, or antivirals for treatment of active infection within 28 days of study screening
    c. Systemic immunosuppressive therapy within 3 months of study screening
    d. Drugs or other compounds that modulate GI motility (including stool softeners, laxatives, or fiber supplements) taken currently or within 7 days of Study screening. Antacid (H2 blockers and PPIs) and antidiarrheal agents are not prohibited.
    Note: Prebiotics and probiotics intake should not be changed during the study (see Section 6.6 for additional details and reference).
10. History of GI surgery (6 months prior to Randomization) including but not limited to bariatric surgery and bowel resection, or history of, or active GI disease(s) that may affect assessment of tolerability, including but not limited to the following:
    a. Inflammatory bowel disease
    b. Irritable bowel syndrome
    c. Autoimmune disease
    d. GI malignancy

11. Participation in an interventional clinical trial or use of any investigational agent within 30 days before Randomization
12. Has a clinically significant or uncontrolled concomitant medical condition that would put the patient at risk or jeopardize the objectives of the study in the opinion of the PI
13. Is considered, in the opinion of the PI, to be unlikely for any reason to be able to comply with study procedures
14. Contraindications, sensitivities, or known allergy to the use of the study product or its components

**Statistical Methods:**

The study will enroll approximately 350 to 400 patients. The enrollment target is chosen for practical reasons. Assuming a 15% attrition rate, this will provide approximately 296 to 340 evaluable patients (148 to 170 per group). The precision of the point estimate in terms of half-width of the 95% confidence interval (CI) of the AE rate is summarized in Table 4 for a range of evaluable number of patients and assumptions on the SP-related TEAE rates.

The analysis of AEs will be carried out on the Safety Analysis Set (Section 8). Summaries will be presented by Medical Dictionary for Regulatory Activities (MedDRA) system organ class and preferred term using frequency counts and percentages by group and overall.

The following endpoints will be analyzed based on the Full Analysis Set. The Kaplan-Meier method will be used to estimate median time to resolution of overall 13 COVID-19 related symptoms, overall 8 cardinal COVID-19 related symptoms, fever and resolution rate specific to each group. The change from Baseline to EOI in overall composite score of 13 COVID-19 related symptoms, overall composite score of 8 cardinal COVID-19-related symptoms, and bedrest time will be analyzed using analysis of covariance (ANCOVA) model. Proportion of patients with oxygen saturation <95% (or <98%) will be summarized by group using frequencies and percentages.

The following endpoints will be analyzed based on the Safety Analysis Set. Proportion of patients experiencing hospital admissions (all cause, and COVID-19-related) will be summarized by group using frequencies and percentages. Measures collected from the Healthcare Provider Wellness Visits and Healthcare Utilizations will be summarized by group using the following descriptive statistics: sample size, mean, standard deviation, median, minimum value, and maximum value or using frequencies and percentages.

More details of the statistical methods are provided in Section 8. Methods related to exploratory objectives and supportive analyses will be described in a separate document such as Statistical Analysis Plan (SAP) which will be developed and completed prior to database lock.

### LIST OF ABBREVIATIONS

| Abbreviation | Expanded Term |
|---|---|
| ADL | Activities of Daily Living |
| AE | Adverse event |
| AESI | Adverse events of special interest |
| ALT | Alanine Aminotransferase |
| ANCOVA | Analysis of Covariance |
| aPTT | Activated Partial Thromboplastin Time |
| AST | Aspartate aminotransferase |
| BID | Twice daily (from the Latin *"bis in die"*) |
| BIPAP | Bilevel Positive Airway Pressure |
| BMI | Body Mass Index |
| CDC | Centers for Disease Control and Prevention |
| CFR | Code of Federal Regulations |
| CI | Confidence interval |
| COVID-19 | Coronavirus Disease 2019 |
| $CO_2$ | Cardon dioxide |
| CPAP | Continuous Positive Airway Pressure |
| CRE | Carbapenem-resistant Enterobacteriaceae |
| CRO | Contract Research Organization |
| CRP | C-reactive protein |
| CSA | Clinical Study Agreement |
| DC | Dendritic cell |
| eCRF | Electronic case report form |
| EDC | Electronic database capture |
| eGFR | Estimated glomerular filtration rate |
| EOI | End of Intake |
| FDA | Food and Drug Administration |
| GCP | Good Clinical Practice |
| GI | Gastrointestinal |
| GPCR | G-protein Coupled Receptors |
| GRAS | Generally recognized as safe |
| HCP | Healthcare Personnel |
| HDL | High density lipoprotein |

| | |
|---|---|
| HSCT | Hematopoietic Stem Cell Transplantation |
| ICF | Informed consent form |
| ICH | International Council for Harmonization |
| INR | International normalized ratio |
| IRB | Institutional Review Board |
| ISF | Investigator Site File |
| KaSK | Kaleido Study Kit |
| LDH | Lactate dehydrogenase |
| LDL | Low density lipoprotein |
| LSM | Least-squares mean |
| MCH | Mean corpuscular hemoglobin |
| MCH | Mean corpuscular hemoglobin concentration |
| MDRO | Multi-drug Resistant Organisms |
| MedDRA | Medical Dictionary for Regulatory Activities |
| MP | Monitoring Plan |
| PI | Principal Investigator |
| PGIC | Patient Global Impression on COVID-19 Condition |
| PP | Per-Protocol |
| PT | Prothrombin time |
| QC | Quality Control |
| QOL | Quality of Life |
| RBC | Red blood cell |
| RDW | Red cell distribution width |
| RSV | Respiratory Syncytial Virus |
| RT-PCR | Reverse Transcription-Polymerase Chain Reaction |
| SAE | Serious Adverse Event |
| SAP | Statistical Analysis Plan |
| SARS-CoV-2 | Novel coronavirus, Severe Acute Respiratory Syndrome 2 |
| SCFA | Short-chain Fatty Acid |
| SD | Standard Deviation |
| SOA | Schedule of Assessments |
| SOB | Shortness of Breath |
| SP | Study product |
| SSC | Supportive Self Care |
| TEAE | Treatment-emergent adverse event |
| VRE | Vancomycin-resistant Enterococcus |

| | |
|---|---|
| WBC | White blood cell |

### 2 INTRODUCTION

#### 2.1 Background

##### 2.1.1 Oligosaccharides and the Gut Microbiome

The gut microbiome is an ecosystem of >10 trillion microbial cells in the intestine and is responsible for functions that human cells cannot carry out independently (Moya 2016). The gut microbiome plays a significant role in human health and disease by affecting nutrient utilization, colonization resistance, development of the immune system, modulation of host metabolism, and other diverse aspects of the host's physiology (Sommer 2013). A person's diet profoundly affects the composition of the gut microbiota, which has a downstream effect on their physiology, immunity and susceptibility to infectious diseases (Kau 2011).

Non- or low-digestible carbohydrates, including oligosaccharides, have been shown to modulate the composition and/or activity of the gut microbiota through influencing its metabolism, thus conferring a beneficial effect on the host (Bindels 2015; Gray 1975). Some dietary oligosaccharides are susceptible to hydrolytic digestion in the stomach and small intestine. The portion of the orally ingested carbohydrate that is not susceptible to hydrolytic digestion reaches the colon where it is available for fermentation by the colonic microbiota. In the colon, there is a vast repertoire of carbohydrate-active enzymes produced by the complex community of resident-commensal bacteria. Fermentation of carbohydrates by gut bacteria results in the production and release of the short-chain fatty acids (SCFAs) acetate, propionate, and butyrate. These SCFAs serve an important role in the maintenance of healthy gut epithelial function.

Non-digestible oligosaccharides have a very well-established safety profile. Several non-digestible carbohydrates are produced using starting materials composed of glucose monomers and are generally recognized as safe (GRAS), or approved as food additives for use in a variety of foods (GRAS 436; GRAS 610; GRAS 711; 21 Code of Federal Regulations [CFR] 172.841), including infant formula (GRAS 233).

A considerable body of information for this group of non-digestible glucose-based carbohydrates has been generated in *in vivo* and *in vitro* toxicology studies (Yokishawa 2013; Burdock 1999; Wils 2008; Bito 2016) and in human clinical studies including many that were reviewed in the Food and Drug Administration's (FDA) scientific review of isolated and synthetic non-digestible carbohydrates published in November 2016 (FDA 2016). Data from animal toxicology studies of these carbohydrates indicate that the effects observed were expected osmotic and fermentative effects with subsequent changes in the gastrointestinal (GI) tract; there was no evidence for direct target organ toxicity, and the effects appear independent of the oligosaccharide composition. Further, adverse effects in human clinical studies are limited to GI adverse effects that are easily managed by decreasing the dose amount (Grabitske 2009).

##### 2.1.2 COVID-19

In December 2019, an outbreak of pneumonia of unknown cause presented. By early January 2020, scientists had isolated a novel coronavirus, severe acute respiratory syndrome coronavirus 2 (SARS-CoV-2; previously known as 2019-nCoV), from these patients with virus-infected pneumonia. In February 2020, this severe acute respiratory syndrome was designated as COVID-19 and has since been characterized as a pandemic by the World Health Organization.

While the complete clinical picture with regards to COVID-19 is not fully known, the clinical spectrum of SARS-CoV-2 infection appears to be wide, encompassing asymptomatic infection, mild upper respiratory tract illness, severe respiratory failure and even death.

As of 11 Nov 2020, the coronavirus COVID-19 has affected 210 countries and territories around the world and two international conveyances. It is estimated that COVID-19 has infected over 50.5 million people worldwide and has been responsible for over 1.25 million deaths. As of November 2020, in the U.S., COVID-19 has affected over 10 million people and has been responsible for over 230,000 deaths making it a major cause of mortality and morbidity (Johns Hopkins Coronavirus Resource Center, 2020).

#### 2.2 Study Rationale

There are few randomized controlled trials assessing the natural history of disease and management for patients with COVID-19 in an outpatient setting. Existing studies to date have focused on improving outcomes in patients who have been hospitalized and/or are in Intensive Care Units.

Kaleido Biosciences, Inc. (Kaleido, Sponsor) has developed technology to produce novel, defined mixtures of oligosaccharides synthesized from food-based monosaccharide sources. These glycans belong to a class of oligosaccharide substances which are accepted as safe by regulators worldwide for use in food. KB109 has been determined to be GRAS for the intended use. FDA regulations recognize self-determination of GRAS status, and KB109 is being investigated in this study under the regulations supporting research with food.

Kaleido's products are orally administered, non-absorbed synthetic proprietary oligosaccharide mixtures that modulate the composition and metabolic output of the gut microbiome. These oligosaccharide mixtures escape digestion by the limited number of human carbohydrate-modifying enzymes in the small intestine and pass into the large intestine where they encounter a vast repertoire of carbohydrate-active enzymes produced by a complex community of resident commensal bacteria. The major products of carbohydrate/oligosaccharide fermentation by gut bacteria are butyric acid, propionic acid, and acetic acid (short-chain fatty acids, SCFAs). These SCFAs serve an important role in the maintenance of healthy gut epithelial function (Arpaia 2013, Smith 2013). Butyrate is used as the primary source of energy for gut epithelial cells and promotes maintenance of epithelial integrity. Maintenance of epithelial integrity is important to prevent inappropriate activation of innate and adaptive immune cells. Butyric acid activates G-protein coupled receptors (GPCRs) displayed on the surface of epithelial and immune cells, as well as inhibits histone deacetylases in the nucleus of these cell types. Propionic acid can also promote gut immune homeostasis by activating these same GPCRs. In addition to producing SCFAs, carbohydrate fermentation results in the growth of commensal bacteria and creates a nutritionally competitive ecosystem in the gut which may restrict the growth of unwanted pathobiont species through multiple mechanisms including acidification of the colon.

Current understanding of the modulation of respiratory virus infectivity by the commensal microbiota of the host and the underlying mechanisms in this regulation are still inadequate. However, there are substantial data supporting a key role for both gut-derived metabolites (e.g., SCFAs) and the direct interaction and migration of immune cells from gut to lung by the common mucosal immune system in pulmonary infections.

Kaleido's *ex vivo* assay has been used to induce and measure the fermentation of different oligosaccharide mixtures, including KB109, by human fecal samples. SCFAs produced through fermentation can be quantified using gas chromatography-mass spectroscopy analysis. The *ex vivo* assay is performed under anaerobic conditions using media that support the growth of commensal bacteria. In *ex vivo* assays testing KB109, it increased the amount of SCFAs produced over water control by ca. three-fold across multiple fecal communities from healthy donors (Figure 1).

**Figure 1 Representative KB109 batch SCFA production average across three healthy fecal communities normalized to water control**

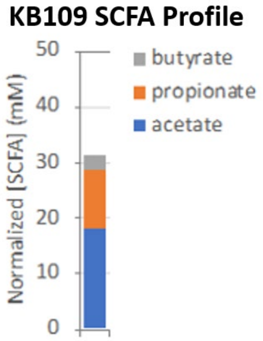

SCFAs modulate host inflammation, control adaptive immunity, and promote immune tolerance locally in the gut as well as systemically (Thaiss 2016; Schirmer 2016). In particular, SCFAs and SCFA-producing taxa have been linked to reduced risk of acquiring viral infections, including corona-viral infections, in at-risk populations such as HSCT patients (Haak 2018). Studies in mice have demonstrated that SCFAs and SCFA-producing microbial taxa, induce virus-specific CD4+ and CD8+ T cells, type 1 interferon, and antibody responses involved in the reduction of viral infection severity (Ichinohe 2011; Trompette 2018). Data has also shown that protection against RSV infection can be conferred by acetate, a metabolite derived from the intestinal microbiota, through induction of IFN-β in the lung through GPR43 and IFNAR-mediated pathways (Antunes 2019). In addition, SCFAs have also been reported to influence macrophage functionality to mitigate neutrophil-mediated tissue damage (Trompette 2018). This is of particular importance as viral infections may be accompanied by an aggressive pro-inflammatory response (Haak 2018) that can elicit a syndrome known as the “cytokine storm”.

In addition to gut microbiome derived metabolites influencing peripheral inflammatory responses, direct activation of host immune cells and pathways by microbiota in the gut has been shown to impact the progression of pulmonary infections. Following influenza virus infection, inflammasome activation can led to migration of dendritic cells (DCs) from the lung to the draining lymph node and T-cell priming. This reveals the importance of commensal microbiota in regulating immunity in the respiratory mucosa through the proper activation of inflammasomes (Ichinohe 2011). It was recently reported that reconstitution of the gut microbiota from wild mice confers potent protective effects to laboratory germ free mice during lethal influenza virus infections, an effect mainly mediated through the prevention of an excessive inflammatory response via IL-10 and IL-13 (Rosshart 2019). These studies and others suggest a dysbiosis in the microbiome community exhibited as a loss in overall commensal diversity or pathobiont overgrowth might contribute to unfavorable outcomes in respiratory infections.

There is potential that patients in this study may suffer secondary infections. Kaleido's *ex vivo* screening platform has shown that the KB109 may reduce the relative abundance of multi-drug resistant organisms (MDRO) that colonize the gut, including carbapenem-resistant Enterobacteriaceae (CRE), Vancomycin-resistant Enterococcus (VRE) and C. difficile. The clinical pathogen strains used were obtained from the Centers for Disease Control and Prevention (CDC) and collaborating laboratories and represent multiple sequence types and geographic regions. The experiments described below used fecal samples obtained from ICU patients, as these patients were heavily treated with IV antibiotics and represent the most dysbiotic microbiomes Kaleido has observed to date (Figure 2). In these experiments, the microbial culture obtained upon fermentation of KB109 underwent community composition analysis using shotgun metagenomic sequencing of the entire community genomic DNA. Comparison of the community composition obtained from incubation with or without KB109 allowed measurement of pathogen reduction. To ensure a robust response, each starting community received a CRE or VRE pathogen spike-in (frequently >70% relative abundance of the total community). Note that fecal samples from patient #1, #6, and #12 had virtually no commensal organisms, which we hypothesize explains the lack of Enterobacteriaceae reduction in these samples. Moreover, single-strain experiments demonstrate KB109 does not support the growth of MDR pathogens directly, as measured by optical density (OD600) of the culture (Figure 3). The carbohydrate monomer glucose served as a positive control and supported robust growth of all pathogens. These data further de-risked the ability of KB109 to inadvertently grow gut-colonizing pathogens. By reducing or completely removing the number of pathogenic organisms that colonize the gut, in addition to improving gut barrier function through the action of microbiome-produced SCFAs, the chance of secondary infection arising from translocation of gut colonizing pathogens is hypothesized to be greatly reduced.

**Figure 2 Reduction of Enterobacteriaceae in ICU patients' microbiota (spiked with CRE *E. coli*) incubated with KB109 relative to water control**

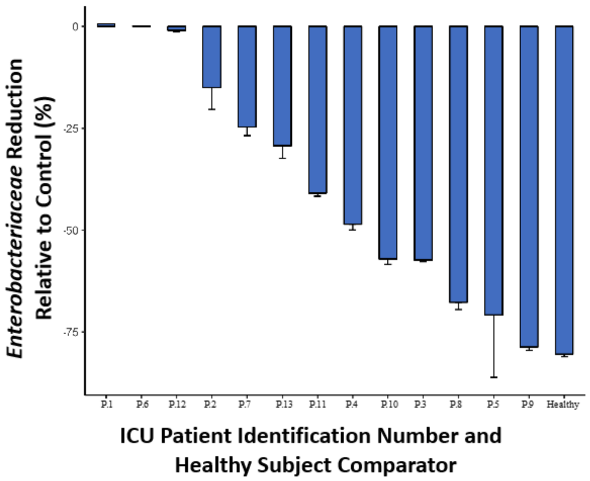

**Figure 3 CRE and VRE growth on KB109 demonstrated similar growth to water control, suggesting an inability of these pathogens to directly utilize KB109**

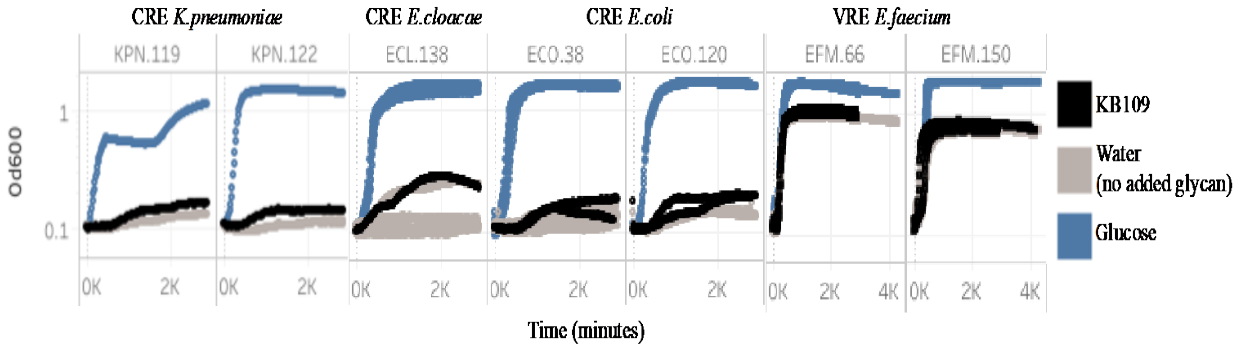

Complications of COVID-19 are known to occur in the patient population eligible for this study, and patients will be monitored for such complications throughout the study. In summary, this is a study designed to safely examine the natural history of disease progression and the influences of the microbiome in patients with mild to moderate symptoms of COVID-19 in the outpatient setting. The study will allow for a better understanding of the treatment and management of this novel infection where there is a paucity of data.

#### 2.3 Benefit/Risk Assessment

KB109 is made from food-based monosaccharide sources, and it belongs to a class of commercially available compounds (non-digestible glucose-based carbohydrates) that have a long history of safe use in food. The Sponsor has engaged an external scientific consulting organization that is frequently used by the food industry to prepare and make GRAS determinations. KB109 was concluded to be Generally Recognized as Safe (GRAS) for use in this study by experts recognized in the field to evaluate GRAS in accordance with the scientific requirements outlined in 21 CFR §170.30(b). This report is a sufficient basis for establishing the GRAS status of KB109 as intended for use as food in this study. FDA regulations recognize self-determination of GRAS status. Although tolerability varies among individuals and among nondigestible carbohydrates, the adverse effects of nondigestible carbohydrates are not qualitatively different from each other and are dose-dependent, quickly reversible, and localized to the GI tract (Grabitske 2009).

Kaleido's orally administered proprietary oligosaccharide mixture, KB109, holds promise in inducing a beneficial profile of SCFA production in the gut while driving the growth of commensal bacteria. Increased SCFA and the modulation in microbiome taxonomy may lead to a more appropriate immune and inflammatory response and may prevent an over-aggressive response, up to and including "cytokine storm" observed in patients with COVID-19. KB109 is determined to be GRAS its intended use in this study.

### 3 STUDY OBJECTIVES AND ENDPOINTS

#### 3.1 Study Objectives

##### 3.1.1 Primary Objective

The primary objective of the study is to evaluate the safety of KB109 in addition to Supportive Self Care (SSC + KB109) compared to SSC alone in outpatients with mild-to-moderate COVID-19.

##### 3.1.2 Secondary Objective

The secondary objective is to evaluate selected measures of health in outpatients with mild-to-moderate COVID-19.

##### 3.1.3 Exploratory Objectives

The exploratory objectives are to evaluate:

- Measures of health in outpatients with mild-to-moderate COVID-19 during the follow-up period
- The changes in laboratory measures, specific biomarkers, serology and viral load in outpatients with mild-to-moderate COVID-19

#### 3.2 Study Endpoints

##### 3.2.1 Primary Endpoint

The primary endpoint is number of patients experiencing study product (SP)-related treatment-emergent adverse events (TEAEs).

##### 3.2.2 Secondary Endpoints

The secondary endpoints are as follows:

- Time to resolution of overall 13 COVID-19 related symptoms which is defined as from Day 1 until the day at which the overall composite score of 13 COVID-19 related symptoms becomes 0 or 1 and remains at 0 or1 for the rest of the Intake Period and for the Follow-up Period. Overall composite score of 13 COVID-19 related symptoms is the sum of 13 COVID-19 related symptom scores (i.e., cough, chills/repeated shaking with chills, muscle pain, fever, headache, anosmia/ageusia, shortness of breath, sore throat, gastrointestinal disturbance/symptoms, diarrhea, fatigue, nasal congestion, and chest tightness (CDC 2020). Each COVID-19 symptom will be recorded by patients on a scale of 0: Absent, 1: Mild, 2: Moderately severe, 3: Very severe. The overall composite score ranges from 0 (no symptoms) to 39 (very severe)
- Time to resolution of overall 8 cardinal COVID-19 related symptoms which is defined as from Day 1 until the day at which the overall composite score of 8 cardinal COVID-19 related symptoms becomes 0 or 1 and remains at 0 or1 for the rest of the Intake Period and for the Follow-up Period. Overall composite score of 8 cardinal COVID-19 related symptoms is the sum of 8 cardinal COVID-19 related symptom scores (i.e., cough, chills/repeated shaking with chills, muscle pain, fever, headache, anosmia/ageusia, shortness of breath, and sore throat)
- Proportion of patients with reduction from Baseline (symptom present at Baseline) in each of 13 individual COVID-19 related symptom at EOI and Follow-up
- Proportion of patients with symptom becomes absent (symptom present at Baseline) at EOI and Follow-up for each of 13 individual COVID-19 related symptom
- Change from Baseline to EOI in overall composite score of 13 COVID-19 related symptoms
- Change from Baseline to EOI in overall composite score of 8 cardinal COVID-19 related symptoms
- Time to resolution of fever (defined as from Day 1 until the day at which a patient's daily maximum temperature achieves and remains below 100.4 °F for the rest of the Intake Period and for the Follow-up Period without an antipyretic medication)
- Proportion of patients with oxygen saturation <95% on Day 14 and Day 35

- Proportion of patients with oxygen saturation <98% on Day 14 and Day 35
- Measures collected from the Healthcare Provider Wellness Visits
- Proportion of patients experiencing hospital admissions during the Intake Period and Follow-up Period (all cause, and COVID-19-related)
- Healthcare Utilizations during the Intake Period and Follow-up Period

##### 3.2.3 Exploratory Endpoints

The exploratory endpoints are as follows:

- Change from Baseline to EOI in individual COVID-19 related symptom score: cough, chills/repeated shaking with chills, muscle pain, fever, headache, anosmia/ageusia, shortness of breath, sore throat, gastrointestinal disturbance/symptoms, diarrhea, fatigue, nasal congestion, and chest tightness
- Individual measures of quality of life (QOL)
- Change from Baseline to EOI in bedrest time measured as patient-assessed daily cumulative total rest (measured in hours)
- Proportion of patients with increases in patient global impression on COVID-19 condition (PGIC)
- Proportion of patients with temperature below 100.4 °F without an anti-pyretic medication
- Change from Baseline (Day 1) in:
  - Laboratory measures
  - Biomarkers of infection, antibody response, and inflammation (e.g., D-dimer, lipocalin, cytokines, IgM/IgG sero-conversion, and Neutralization Assays)

### 4 STUDY DESIGN

#### 4.1 Overall Design

This study is a randomized, controlled, multi-site, open label clinical study. It is intended to assess KB109 on safety as well as measures of signs, symptoms, healthcare utilization (including hospitalizations), laboratory/biochemical indices and QOL measures in outpatients who have tested positive with COVID-19, have mild-to-moderate disease, and have been advised to manage their disease at home with SSC under quarantine protocols set forth by the CDC (CDC 2020) or local ordinances or practices as advised by their healthcare provider.

Approximately 350–400 patients will be randomized (1:1) to receive either SSC + KB109 or remain on SSC alone. The randomization will be stratified by site/center, age subgroup (≥18 to <45 years, ≥45 to <65 years, ≥65 years), and comorbidity status (Yes, No); see Section 8.1 for further details. The study consists of a Screening/Randomization Visit, Intake Period (14 Days) followed by a 21-day Follow-up Period (Figure 4).

Details of the procedures and assessments conducted during the study are provided in Section 4.3, and the timing for each is presented in the Schedule of Assessments (SOA; Table 1) and summarized in Figure 4.

To be eligible for the study, patients must be at least 18 years of age, have tested positive for COVID-19, and be medically stable at study entry per eligibility criteria. Patients must have also been advised by a healthcare provider that self-management of COVID-19 is indicated; eligibility criteria are detailed in Section 5.1 and Section 5.2.

Patients will be recruited via outpatient clinics or testing centers performing SARS-CoV-2 testing or via online portals. They will have an opportunity to voluntarily consent into the study either at the time they are making their outpatient clinic appointment, at the outpatient testing center/clinic itself, or following discharge from the outpatient clinic. SARS-CoV-2 tests (RT-PCR test, antigen test or equivalent test to detect active infection) are part of the standard of care, though may be provided as part of the study procedures at some sites. Results from a local testing clinic that is not the study site location may be used to determine eligibility virtually (not at site). Patients who are symptomatic at the time of getting a positive COVID-19 test result and meet eligibility criteria will enter Part 2 of the study within 48 hours of testing positive. Patients who are pre-symptomatic at the time of testing, develop symptoms within 7 days of testing positive and meet eligibility criteria will enter Part 2 of the study within 5 days of the symptoms showing. All enrolled patients will have same level of care and same procedures conducted.

**Part 1**

This study comprises two parts. In Part 1, patients who consent to the study will undergo a full assessment of inclusion and exclusion criteria (Section 5.1 and Section 5.2). If feasible blood samples for Baseline hematology, chemistry, biomarkers and serological markers of immunity, and nasal and oropharyngeal swabs for quantitative vial load assessments (research purposes only) will be taken (See Section 4.3.9 and Table 1); sample collection may occur at the patient's home by a healthcare professional, remotely, or at the study site or clinic. Patients will be asked to begin recording COVID-19-related symptoms using a secure website called "TrialPace™ diary" (TrialPace™) with a unique log-in and password.

It is possible that patients may only consent to Part 1 of this study initially. If a positive test result for COVID-19 is received, then the patient may be contacted by the study team (or designee such as a centralized telemedicine provider, physician or nurse practitioner) to review study eligibility criteria and obtain informed consent.

Information entered into TrialPace™ for patients who are deemed ineligible for the study will not be used for any purpose.

**Part 2**

Only patients who have a positive test result for COVID-19 can continue to Part 2 of this study. Part 2 of the study comprises an Intake Period and a Follow-up Period. In Part 2, following confirmation of eligibility criteria, the site will notify an independent, third party vendor that a patient is eligible for randomization no later than 48 hours after a positive test result is received by the patient for COVID-19. Upon randomization (SSC + KB109 or SSC alone), a Kaleido at-home Study Kit (KaSK) and SP (KB109) and dosing instructions (as applicable) will be shipped via overnight delivery to the patient's home. The KaSK will include a thermometer, a pulse oximeter, and telemedicine contact information. Patients should continue to record COVID-19-related symptoms using TrialPace™ until the KaSK is delivered.

**Part 2, Intake Period**

The Intake Period (Days 1-14) will begin the morning after receipt of the KaSK. All patients will continue to follow the SSC guidance as provided by the treating healthcare provider throughout study participation. In the Intake Period, all patients will continue to record their daily COVID-19-related symptoms, selected COVID-19 signs, responses to questions related to QOL measures, healthcare utilization measures, and concomitant medications taken in the previous 24 hours using TrialPace™ where appropriate. Patients randomized to SSC + KB109 will begin consuming KB109 on Day 1 after recording the required information. All KB109 usage will be noted in the patient's SP log. For all patients in both groups, temperature and oxygen saturation will be measured and recorded as needed throughout the day prior to taking anti-pyretic medication.

Wellness visits by telephone call between Day 1 and Day 14 will be conducted to follow-up on the patient's health status, to ascertain compliance with SP usage or completion of TrialPace™ questions, and to reeducate the patient on the importance of adherence to study instructions including SP usage (where applicable). When wellness and telemedicine visits fall on the same day, they will be combined as a telemedicine visit.

On Day 14, all patients will undergo a telemedicine visit where the following will be conducted: an abbreviated physical examination, an assessment of safety and other protocol-specified measures of health, and an evaluation of whether follow-up treatment is recommended due to a deterioration of COVID-19 symptoms. Patients in the SSC + KB109 group will stop taking KB109 on Day 14. If feasible blood samples for clinical chemistries, biomarkers and serological measures of immunity, and nasal/oropharyngeal swabs for quantitative vial load assessments (research purposes only), will be collected; sample collection may occur at the patient's home by a healthcare professional, remotely, or at the study site or clinic.

**Part 2, Follow-up Period**

On Day 15, all patients will enter the Follow-up Period (Days 15 to 35) where COVID-19 signs, symptoms and healthcare utilization indices will be collected weekly in TrialPace™ where appropriate. On Day 14, the Investigator will instruct the patients to stop the intake and return the SP during the Follow-up Period (Days 15 to 35). In addition, wellness visits by telephone call will be conducted on Days 21, 28, and 35 to follow-up on the patient's health status.

On Day 35, all patients will undergo another telemedicine visit where the following will be conducted: an abbreviated physical examination, an assessment of safety and other protocol-specified measures of health, and an evaluation of whether follow-up treatment is recommended. When wellness and telemedicine visits fall on the same day, they will be combined as a telemedicine visit. If feasible blood samples for clinical chemistries, biomarkers and serological measures of immunity, and nasal/oropharyngeal swabs for quantitative vial load assessments (research purposes only) will be collected; sample collection may occur at the patient's home by a healthcare professional, remotely, or at the study site or clinic.

For convenience, standard terms language included in International Conference for Harmonization (ICH) guidance relevant to the conduct of clinical trials are used in this protocol. An example of such a term used in this protocol is "treatment emergent," included in ICH E9. Use of this standard term is not intended to connote treatment with a medicinal product administered (ICH E9 2018).

Throughout the study, safety will be monitored by TEAEs, clinical signs and symptoms, and selected vital signs (i.e., temperature and oxygen saturation). Concomitant medications/supplements will be recorded throughout the study. Any patient experiencing a TEAE, or intolerable GI symptoms will be evaluated by the principal investigator (PI) or telemedicine provider for possible interruption and/or down-titration of SP dose to the previously tolerated level in consultation with the medical monitor.

In the event of significant worsening of COVID-related symptoms or two sequential pulse oximetry readings of <94% within 2 minutes, patients will be instructed to seek medical attention immediately (see Section 6.7).

In the event of worsening GI symptoms of other attributable AEs, the SP may be down titrated or temporarily interrupted (see Section 6.8). Details of the patient's SP dose (including the date, time, and amount) will be recorded in SP dosing logs.

See Section 4.3 for further information on the study procedures, and Table 1 for the SOA.

**Figure 4 Study Design**

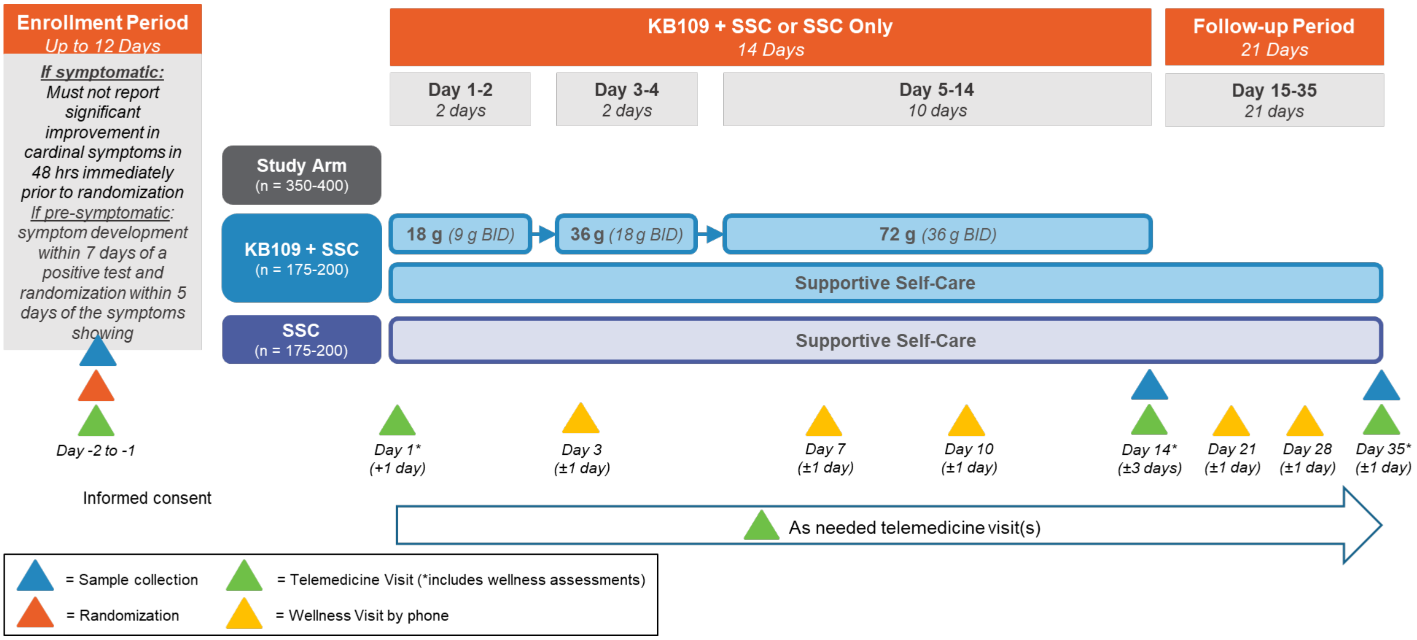

*Abbreviations: BID = Twice daily dosing; SSC = Supportive self-care*

An interim analysis is planned when approximately 40% of the total randomized patients have completed or discontinued prior to completion of the EOI period (See Section 8.4.6).

#### 4.2 Study Duration

The total duration of the study per study patient is up to 47 days from Screening/Randomization (up to 12 days [Day -12] for an asymptomatic patient) through to the end of the Follow-up Period (Day 35); see Figure 4.

#### 4.3 Study Procedures

Details of the individual study procedures are provided in Section 4.3.1 through Section 4.3.11. Timing for when each procedure is to be conducted is presented in the SOA below (Table 1). All self-assessments and patient-reported responses will be recorded in TrialPace™ using the patient's unique log-in and password.

#### Table 1. Schedule of Assessments

| Assessment | Part 1 | Part 2 | | | | | | | | | | |
|---|---|---|---|---|---|---|---|---|---|---|---|---|
| | Screening | Intake Period: (SSC + KB109 or SSC Only) | | | | | | Follow-up Period | | | | ET Visit |
| | Up to D -12 to -1 | D 1 *(+1 D)* | D 2 | D 3 *(±1 D)* | D 7 *(±1 D)* | D 10 *(±1 D)* | D 14 *(±3 D)* | D 15 to D 20 | D 21 *(±1 D)* | D 28 *(±1 D)* | D 35 *(±1 D)* | |
| Informed Consent | X | | | | | | | | | | | |
| Registration in TrialPace™ | X | | | | | | | | | | | |
| Demographics and medical history | X | | | | | | | | | | | |
| Confirm eligibility criteria[1] | X | | | | | | | | | | | |
| Randomization[2] | X | | | | | | | | | | | |
| Consume KB109 for patients in this group (SSC + KB109) | | X | Daily | | | | X | | | | | |
| Physical Examination | X[3] | X[4] | | | | | X[4] | | | | X[4] | X[4] |
| Height, weight, BMI calculation | X | | | | | | | | | | | |
| Patient-assessed COVID-19 Signs (temperature and oxygen saturation)[5] | X | X[6] | Daily | | | | X | | X | X | X | X |
| Patient-assessed Wellness Check COVID-19 Condition[7] | X | X[6] | Daily | | | | | | | | | X |
| Patient Global Impression on COVID-19 Condition[8] | | | Daily | | | | | | | | | X |
| Quality of life indices | | | | | | | X | | | | X | |
| Healthcare utilization questions | | | Daily | | | | | | | | | X |
| Patient-assessed Bedrest Time | | X[6] | Daily | | | | | | | | | X |
| Nasal swab[9] | X | | | | | | X | | | | X | |
| Oropharyngeal swab[9] | X | | | | | | X | | | | X | |
| Blood samples[9] | X | | | | | | X | | | | X | |
| Urine Pregnancy test[10] | X | | | | | | | | | | | |
| Telemedicine Visit[11] | | X | | | | | X | | | | X | X |

| | | | | | | | | | | | |
|---|---|---|---|---|---|---|---|---|---|---|---|
| Wellness Visit by telephone | | X[12] | | X | X | X | X[12] | | X | X | X[12] |
| AE review | Throughout the study | | | | | | | | | | |
| Concomitant medication | Throughout the study | | | | | | | | | | |
| Return of study product | | | | | | | | X[13] | | | |

*Abbreviations: AE = Adverse event; BMI = Body mass index; D = Day; ET = Early termination; KaSK = Kaleido at-home Study Kit; SSC = Supportive self-care*

[1]Note that the patient will have had a SARS-CoV-2 test (RT-PCR test, antigen test or equivalent test to detect active infection). Positive SARS-CoV-2 test results from a local testing clinic that is not the study site location may be used to determine eligibility virtually (not at site). Hospital, academic, or industry-based assays will be acceptable for the diagnosis of COVID-19; home-based tests are not accepted for this study.

[2] If symptomatic and currently positive for COVID-19 diagnostic test, the patient must not report significant improvement in their cardinal COVID-19 symptoms in the 48 hours immediately prior to randomization.

[3]The physical examination at screening can be a) a basic face-to-face physical examination at the patient's outpatient clinic visit, b) based on findings from a physical examination conducted as part of the standard patient intake evaluation into an emergency department or testing clinic (at the discretion of the Investigator), or c) an abbreviated telemedicine-based physical examination (see Section 4.3.3 for further details).

[4]Abbreviated telemedicine-based physical examinations (see Section 4.3.3).

[5] patients will receive the thermometer and pulse oximeter device in the KaSK delivered to their home prior to Day 1, at which time they will begin to record their temperature and oxygen saturation in the TrialPace™ diary.

[6]These measures should be completed prior to the first dose of KB109 for patients in the SSC + KB109 group.

[7]The Patient-assessed Wellness Check COVID-19 Condition (Section 4.3.4) includes patient-assessed COVID-19 symptoms and signs. Responses are recorded in TrialPace™.

[8]The Patient Global Impression on COVID-19 Condition (PGIC) responses are recorded in TrialPace™.

[9]See Section 4.3.9 for the blood tests. If feasible blood samples for clinical chemistries, biomarkers and serological measures of immunity, and nasal/ oropharyngeal swabs for quantitative viral load assessments (research purposes only) will be collected; sample collection may occur at the patient's home by a healthcare professional, remotely, or at the study site or clinic.

[10]Pregnancy status for all females of childbearing potential (as defined in accordance hospital policies and procedures at the patient's outpatient clinic visit) may be collected by patient self-report or urine pregnancy test.

[11]Telemedicine Visits will take place on Days 1, 14, 35, and at an Early Termination Visit, and may also take place throughout the study as needed.

[12]Wellness visit by telephone will be included in the telemedicine visit when they fall on the same day.

[13]Pateints will return the SP during the follow-up period.

##### 4.3.1 Patient Demographics and Baseline Characteristics

Demographic information and baseline characteristics obtained will include age, sex, race, ethnicity, weight, and height.

##### 4.3.2 Medical History and Concomitant Medications/Procedures

The medical history of the patient will be obtained by the PI or qualified designee. Medical history will be recorded and should include prior/existing medical conditions and surgical procedures, physical examination findings and clinically significant laboratory abnormalities.

Medication history information to be obtained also includes any medication (prescription or nonprescription) and any dietary or nutritional supplement relevant to eligibility criteria stopped at or within 28 days before signing of informed consent.

Concomitant medications and procedures will be recorded throughout the study.

##### 4.3.3 Physical Examination

At screening, in order to minimize contact between patients and providers, physical examinations to determine eligibility may be conducted via one of the following methods:

a) A basic face-to-face physical examination performed by a qualified healthcare provider in accordance hospital policies and procedures at the patient's outpatient clinic visit. The examination will include a general assessment (e.g., overall appearance, orientation), pulmonary auscultation, and an assessment for work of breathing (e.g., speech cadence, lifting hands above head, observing ambulation).

b) At the discretion of the Investigator, findings from a physical examination conducted as part of the standard patient intake evaluation at an emergency department or testing clinic can be used to assess relevant inclusion/exclusion criteria provided the physical examination was done within 24 hours of consent and diagnostic COVID-19 testing.

c) An abbreviated telemedicine-based physical examination comprising a general assessment (e.g., overall appearance, orientation) and an assessment for work of breathing (e.g., speech cadence, lifting hands above head), and will be performed according to guidelines outlined via the central telemedicine provider.

Abbreviated telemedicine-based physical examinations will be conducted for all post-screening assessments (see Table **1**). All findings from this examination will be recorded on source documents.

Any new abnormal findings upon physical examination noted after informed consent is obtained and deemed clinically meaningful by the PI or designee may be recorded in the eCRF as an AE.

##### 4.3.4 Patient-assessed COVID-19 Condition

Patients will begin reporting COVID-19-related symptoms in TrialPace™ after consent to the study (Part 1) following the outpatient clinic visit for evaluation of suspected COVID-19 infection. Responses should be recorded consistently upon wakening in the morning. Patients should always complete the Patient-assessed COVID-19 score prior to the PGIC.

Following randomization in Part 2 of the study, patient-reported COVID-19 symptom measurements will continue to be performed in the morning upon wakening and according to the SOA (Table 1). See Section 6.7 regarding patient instructions in the event COVID-19 symptoms worsen.

###### 4.3.4.1 Patient-assessed COVID-19 Symptom Score

Symptom severity will be assessed based on how the patient is feeling at the moment. This will be recorded in TrialPace™.

Patients will self-assess the following 8 cardinal symptoms known to be associated with COVD-19 (CDC 2020):

1. Fever
2. Chills/Repeated shaking with chills
3. Cough
4. Shortness of breath
5. Headache
6. Muscle pain
7. Anosmia/ageusia
8. Sore throat

Additionally, patients will also report on the following 5 symptoms:

1. Gastrointestinal disturbance/symptoms (other than diarrhea as this will be assessed separately)
2. Diarrhea
3. Fatigue
4. Nasal congestion
5. Chest tightness

Patients will be instructed to rate their symptoms on a scale of 0-3 where:

- 0: Absent (no symptoms evident)
- 1: Mild (symptom present but easily tolerated)
- 2: Moderately severe (definite awareness of symptoms; bothersome but tolerable)
- 3: Very severe (hard to tolerate; interferes considerably with daily activity)

###### 4.3.4.2 Patient-assessed COVID-19 Signs

Following randomization, selected patient-assessed COVID-19 signs will be performed daily, upon wakening, as needed and according to the SOA (Table 1).

Patients will self-assess temperature and oxygen saturation according to the instructions for use of the thermometer and pulse oximeter included in their KaSK and will record measures in TrialPace™. See Section 6.7 regarding patient instructions in the event COVID-19 signs and symptoms worsen.

##### 4.3.5 Patient Global Impression on COVID-19 Condition (PGIC)

From Day 2 onwards, daily COVID-19 symptom severity will be assessed reflectively based on how the patient felt over the past 24 hours. This will be recorded in TrialPace™.

Over the past 24 hours, the patient would be asked to rate overall COVID-19 condition change into 7 categories: Very much worse, Much worse, minimally worse, no change, minimally improved, much improved, Very much improved.

##### 4.3.6 Quality of Life Indices

Following randomization, on Days 14 and Day 35, during the telemedicine visit, the telemedicine provider will ask the following questions to assess QOL (Table 1). Telemedicine providers must read the questions as they are written and cannot provide the patient with assistance on interpreting the meaning of questions as it is paramount that the interpretation be left up to the patient.

1.) Overall, how would you rate your health during the past 14 (or 21) Days?
*Rated as: Excellent, Very Good, Good, Fair, Poor, Very Poor*

2.) During the past 14 (or 21) Days, how much did your COVID-19 symptoms limit your physical activities (such as walking or climbing stairs)?
*Rated as: Not at all, Very little, Somewhat, Quite a lot, Could not do physical activities*

3.) How much bodily pain have you had during the past 14 (or 21) Days?

*Rated as: None Very mild, Mild, Moderate, Severe, Very severe*

4.) During the past 14 (or 21) Days, how much energy did you have?
*Rated as: Very much, Quite a lot, Some, A little, None*

5.) During the past 14 (or 21) Days, how much have you been bothered by emotional problems (such as feeling anxious, depressed or irritable)?
*Rated as: Not at all, Slightly, Moderately, Quite a lot, Extremely*

6.) Thinking about any emotional problems you may have had such as feeling, anxious, depressed or irritable, for how many days during the past 14 (or 21) Days was your emotional health not good?
*Rated as number of days from 1-14 days at the end of the Intake Period (Day 14) and 15-35 days at the end of the Follow-up Period.*

7.) Thinking about your overall physical health for how many days during the past 14 (or 21) Days was your physical health not good?
*Rated as number of days from 1-14 days at the end of Intake Period (Day 14) and 15-35 days at the end of the Follow-up Period.*

*8.)* During the past 14 (or 21) Days, how much did your COVID-19 symptoms interfere with your normal sleep patterns?
*Rated as: Not at all, Slightly, Moderately, Quite a lot, Extremely*

##### 4.3.7 Healthcare Utilization

From Day 2 onwards, patient-assessed measures of healthcare utilization will be performed every morning upon wakening until the end of the study (Intake Period plus Follow-up Period) (Table 1).

Measures will include:

- Prescription Medication Use – recorded as reason, type, dose and frequency
- Over-the-counter Medication Use – recorded as reason, type, dose and frequency
- Supplement Use – recorded as reason, type, dose and frequency
- Emergency room or Urgent Care visit – Rated as Y/N; if Y then reason will be recorded
- Medical provider visit – Rated as Y/N; if Y then reason as well as if this is a visit for a co-existing condition (mental health visit, physical therapy, etc.) will be recorded;
- Hospital stay – Rated as Y/N; if Y then reason will be recorded

##### 4.3.8 Patient-assessed Bedrest Time

Patients will be asked to estimate the total amount of time (in hours) of bedrest each day. For the purposes of this protocol, bedrest is defined as all rest including resting on the couch or recliner.

Patients will begin self-assessing total daily bedrest (in hours) at the time they provide consent, each morning upon wakening in a reflective manner (reflecting back over the cumulative time of bedrest from the previous day).

Following randomization, patients will self-assess total daily bedrest in a reflective manner (reflecting back over the previous day) the total amount of their bedrest time in hours and according to the SOA (Table 1).

##### 4.3.9 Laboratory and Inflammatory Biomarkers

If feasible blood samples for hematology, clinical chemistries, and serological measures of immunity, and nasal/oropharyngeal swabs for quantitative viral load assessments (research purposes only) will be collected (see Table 1 and Table 2). Samples collection may occur at the patient's home by a healthcare professional, remotely, or at the study site or clinic. Sampling will be in accordance with acceptable laboratory procedures; additional analyte values may also be reported, consistent with the study endpoints and objectives.

**Table 2 Clinical Laboratory Tests**

| Hematology | Chemistry | | Nasal and Oropharyngeal Swabs |
|---|---|---|---|
| RBC<br>RDW<br>MCH<br>MCHC<br>WBC with differential (% and absolute)<br>Hemoglobin<br>Hematocrit<br>Platelets<br>aPTT<br>PT<br>INR | ALT<br>Albumin<br>Alkaline phosphatase<br>AST<br>Total bilirubin<br>Total protein<br>Creatinine<br>Blood urea nitrogen<br>Magnesium<br>Phosphorus<br>hsCRP<br>Ferritin<br>D-dimer | Potassium<br>Sodium<br>Glucose<br>Chloride<br>Bicarbonate or $CO_2$<br>Calcium<br>eGFR<br>Total cholesterol<br>HDL cholesterol<br>LDL cholesterol<br>Triglycerides<br>LDH | COVID-19 Quantitative Viral Load (research purposes only) |

*Abbreviations: aPTT: activated partial thromboplastin time, ALT: alanine aminotransferase, aPTT: activated partial thromboplastin time, AST: aspartate aminotransferase, $CO_2$: carbon dioxide, CRP: C-reactive protein, eGFR: estimated glomerular filtration rate, HDL: high density lipoprotein, INR: international normalized ratio, LDH: lactate dehydrogenase; LDL: low density lipoprotein, MCH: mean corpuscular hemoglobin, MCHC: mean corpuscular hemoglobin concentration, PT: prothrombin time, RBC: red blood cell, RDW: red cell distribution width, WBC: white blood cell.*
Note: the list of laboratory tests may not be inclusive of all tests conducted throughout the study.

Blood samples may also be assessed for markers of infection, antibody response, and inflammation such as:

- D-dimer, lipocalin, cytokines, IgM/IgG sero-conversion, and Neutralization Assays

Instructions relating to blood and nasal and oropharyngeal swab collection methodologies storage requirements, and shipment requirements will be detailed in the Laboratory Manual.

##### 4.3.10 Telemedicine Visit

As outlined in the SOA (Table 1), a healthcare provider (PI or designee) or a centralized telemedicine vendor will conduct a telemedicine study visit. This visit may be used to interview and collect safety information along with other protocol-specified measures of health as needed.

##### 4.3.11 Healthcare Provider Wellness Visit by Telephone

As outlined in the SOA (Table 1), a healthcare provider (PI or designee) or a centralized telemedicine vendor will conduct a wellness visit by telephone. This phone call to the patient will be conducted to follow-up on the patient's health status and to ascertain compliance with study procedures.

### 5 STUDY POPULATION

#### 5.1 Inclusion Criteria

To be considered for enrollment into this study, each patient must meet **all** of the following Inclusion Criteria:

1. Be male or female, ≥18 years of age
2. Be willing and able to give informed consent, and provide medical history, and secondary contact information
3. If symptomatic at the time of COVID-19 testing, the extended symptom(s), 8 cardinal plus additional 5 symptom(s) (see Section 4.3.4.1 for reference) must be new or worsening at baseline and must not have been present (see Section 4.3.4.1 for reference) for more than 5 days. Symptomatic patients must be screened and randomized within 48 hours of a positive test.
4. If pre-symptomatic at time of COVID-19 testing, new cardinal symptoms (see Section 4.3.4.1 for reference) must be reported within 7 days of a positive test, and the patient must be screened and randomized within 5 days of them developing symptoms

5. Mild to moderate COVID-19 and self-reported outpatient management indicated by their healthcare provider
6. Able to adhere to the study visit schedule and other protocol requirements
7. Has consistent internet or cellphone access with a data plan, and access to a smartphone, tablet or computer

#### 5.2 Exclusion Criteria

Patients who meet **any** of the following Exclusion Criteria will not be enrolled into the study:

1. In the Primary Investigator's judgement, patients likely to require hospitalization for COVID-19
2. Patients who are hospitalized for in-patient treatment or currently being evaluated for potential hospitalization at the time of informed consent for conditions other than COVID-19
3. History of chronic lung disease with chronic hypoxia
4. History of documented cirrhosis or end-stage liver disease
5. Ongoing requirement for oxygen therapy
6. Shortness of breath in resting position
7. Diagnosis of sleep apnea requiring Bilevel Positive Airway Pressure (BIPAP) / Continuous Positive Airway Pressure (CPAP)
8. Female patients who are pregnant, trying to become pregnant or lactating.
9. Concurrent use of any of the following medications:
    a. Therapy with an immunomodulatory agent within 12 months of study screening
    b. Systemic antibiotics, antifungals, or antivirals for treatment of active infection within 28 days of study screening
    c. Systemic immunosuppressive therapy within 3 months of study screening
    d. Drugs or other compounds that modulate GI motility (including stool softeners, laxatives, or fiber supplements) taken currently, or within 7 days of Study screening. Antacid (H2 blockers and PPIs) and antidiarrheal agents are not prohibited.
    Note: Prebiotics and probiotics intake should not be changed during the study (see Section 6.6 for additional details and reference).
10. History of GI surgery (6 months prior to Randomization), including but not limited to bariatric surgery and bowel resection, or history of, or active GI disease(s) that may affect assessment of tolerability, including but not limited to the following:
    a. Inflammatory bowel disease
    b. Irritable bowel syndrome
    c. Autoimmune disease
    d. GI malignancy
11. Participation in an interventional clinical trial with or use of any investigational agent within 30 days before Randomization

12. Has a clinically significant or uncontrolled concomitant medical condition that would put the patient at risk or jeopardize the objectives of the study in the opinion of the PI
13. Is considered, in the opinion of the PI, to be unlikely for any reason to be able to comply with study procedures
14. Contraindications, sensitivities, or known allergy to the use of the study product or its components

#### 5.3 Screen Failures

Patients who provide informed consent but do not meet the study entry criteria during Screening and consequently are not enrolled in the study are screen failures. Patients may not be re-screened.

#### 5.4 Enrollment

Patients will be randomized in a 1:1 ratio to either SSC + KB109 or SSC alone; see Section 6 for further information on the SP. Consented patients who are withdrawn from the study prior to randomization are deemed screen failures; patients withdrawing after randomization will not be replaced nor re-consented for new participation.

#### 5.5 Withdrawal of Patients

Patients have the right to withdraw from the study at any time for any reason, without giving a reason and without penalty or loss of benefits they are entitled to. The investigator also has the right to withdraw patients from the study if this is in the best interest of the patient.

Patients will be discontinued from the study in the event of any of the following:

- Patient independently withdraws consent from the study (e.g., patient declines further participation in the study)
- PI determines that the patient has developed a concurrent illness, condition, or an AE due to which continued participation in the study and/or SP dosing is considered potentially harmful to the patient and withdrawal is in the best interest of the patient
- Patient discontinues from SP dosing (i.e., due to development of an AE), but continues his/her consent to participate in the study for follow-up only (e.g., for continued collection of AE information)
- Patient failure to comply with study requirements (e.g., failure to properly consume the SP or failure to comply with other protocol-specific assessments) resulting in protocol deviations that make the interpretation of the data not feasible
- Patient becomes pregnant

Patient requires hospitalization due to worsening COVID-19

##### 5.5.1 Handling of Discontinuation of SP and Early Terminations

If the PI discontinues a patient from SP due to development of an AE, the patient wishes to withdraw from the study, or the patient is terminated from the study for any other reason every effort should be made to:

- Complete the Early Termination visit assessments, as directed in the SOA (Table 1)
- Monitor the patient for resolution of any existing AEs, and onset of new AEs for 3 weeks (21 days) after the last dose of SP.

The data should be documented on the appropriate eCRF in the electronic database capture (EDC) system.

If, due to extenuating circumstances, the PI and/or study site team are concerned that the patient may not be able to safely complete all the required visits and assessments the PI should contact the Sponsor's Medical Monitor to discuss whether an early termination visit should occur to enable the return of the SP, and the completion of key end of intake assessments/measures, while ensuring the patient's safety.

##### 5.5.2 Premature Termination or Suspension of Study

This study may be suspended or prematurely terminated at the Sponsor's discretion. Written notification documenting the reason for study suspension or termination will be provided by the Sponsor to the PI. If the study is prematurely terminated or suspended, the PI will promptly inform the Institutional Review Board (IRB) and will provide the reason(s) for the termination or suspension.

Circumstances that may warrant termination or suspension include, but are not limited to:

- Determination of unexpected, significant, or unacceptable risk to patients as determined by the ongoing review of aggregate safety data by the medical monitor

Circumstances that may warrant termination or suspension of an individual site include but are not limited to:

- Insufficient compliance with protocol requirements by the site
- Site submits data that are consistently insufficient, incomplete, and/or unevaluable

If the study is suspended temporarily at a site, the study may resume once concerns about safety, protocol compliance, and/or data quality are addressed and satisfy the Sponsor and/or IRB.

##### 5.5.3 Lost to Follow-up

A patient will be considered lost to follow-up if he or she repeatedly cannot be contacted for scheduled telemedicine visits or sample collections and is unable to be contacted by the study site.

If a patient fails to attend a required study visit, the site must:

- Attempt to contact the patient and reschedule the missed visit as soon as possible
- Counsel the patient on the importance of maintaining the assigned visit schedule
- Ascertain whether the patient wishes to and/or should continue in the study

Before a patient is deemed lost to follow-up, the PI or designee must make every effort to regain contact with the patient until the end of the study, including potentially contacting the emergency contact listed by the patient. These contact attempts should be documented in the patient's study record. Should the patient continue to be unreachable, he/she will be considered lost to follow-up.

### 6 STUDY PRODUCTS

#### 6.1 Description of Study Product

### 6.1.1 KB109

As depicted in Figure 5, KB109 is a mixture of polymers of glucose, galactose, and mannose in proportions of approximately 45%, 45%, and 10%, by weight respectively. The formula is $H\text{-}[C_6H_{10}O_5]_n\text{-}OH$, where the total number of monomer units in a single polymer of the mixture ranges from 2 to approximately 60 (n = 2–60), with a mean value for the mixture of approximately 12.6 monomer units. Each monomer unit may be unsubstituted, singly, doubly, or triply substituted with another glucose, galactose, or mannose unit by any glycosidic isomer.

**Figure 5 KB109 Structure**

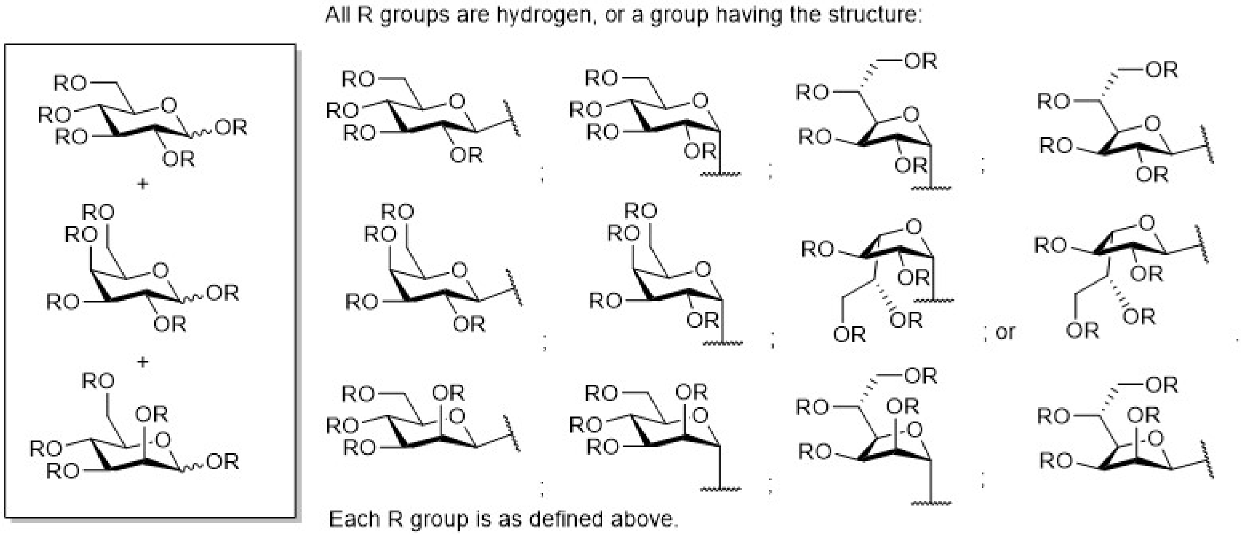

#### 6.2 SP Dosing, Preparation and Administration

Patients randomized to SSC + KB109 will consume their SP orally BID (at least 8 hours apart). Preparation of the SP involves adding 120 mL of water into a glass/container, instilling the directed amount of SP powder on top, and mixing to dissolve prior to drinking as a beverage. The SP should be consumed within 1 hour after reconstitution.

Dosing of KB109 is detailed in Table 3.

**Table 3. Dose Up-titration of the Study Product**

| SP Administration Period | Dose (KB109) |
|---|---|
| Days 1–2<br>Days 3–4<br>Days 5–14 | 9 g BID (18 g/day)<br>18 g BID (36 g/day)<br>36 g BID (72 g/day) |

*Abbreviations: BID: Twice daily; SP: Study Product.*

Doses should be taken at the same time each day. The morning dose should be taken by noon each day, and the evening dose should be taken approximately 8 hours after the morning dose. A missed morning dose may still be taken before noon on the same day; a missed evening dose may be taken within 4 hours of the routine daily evening administration dose.

Details of the patient's SP dosing (including the date, approximate time, and amount) will be recorded in SP dosing logs.

#### 6.3 SP Storage and Accountability

The SP is to be dispensed by the trained team at the storage and shipping vendor. Patients will be instructed to return used and unused SP to the storage and shipping vendor, per the instructions included in the SP packaging. At the termination of the study or at the request of the Sponsor, the storage and shipping vendor will destroy or return unused SP and all partially dispensed or empty packets.

#### 6.4 Unblinding

This is an open label study. There are no requirements for unblinding.

#### 6.5 Concomitant Medications/Dietary Supplements

All concomitant medications and dietary supplements taken after consent is provided will be recorded. Medications to be reported include all prescription medications, over-the-counter medications, and other non-prescription medications taken by a patient during study participation. All dietary supplements taken by patients during study participation will be reported as well. For this study, a prescription medication is defined as a medication that can be prescribed only by a properly authorized/licensed clinician.

Information regarding eligibility criteria relating to the use of certain medications can be found in Section 5.1 and Section 5.2 of the protocol.

See Section 6.6 for prohibited medications and dietary supplements.

#### 6.6 Prohibited Medications/Dietary Supplements

Medications or dietary supplements prohibited at screening and during the study provided that appropriate safety considerations are followed, including:

- GI increasing motility agents such as stool softeners, laxatives, or fiber supplements within 7 days prior to screening and during the study (intake period and follow up).
- Antacid (H2 blockers and PPIs) and antidiarrheal agents are not prohibited
- Systemically administered immunosuppressant medications 3 months of study screening and during the study (intake period and follow up)
- Therapy with any immunomodulatory agent 12 months previous of study screening and during the study (intake period and follow up)

- Systemic antibiotics, antifungals, or antivirals for treatment of active infection 14 days previous of study screening and during the study (intake period and follow up)

Note: Prebiotics and/or probiotics intake is permitted provided the patient has been taking them consistently in the 28 days prior to screening and continues to do so during the study (i.e., no changes in dose, frequency, or type of prebiotic/probiotic). Patients not taking prebiotics and/or probiotics at study entry are prohibited from initiating them during the study. There are no restrictions on diet.

Additionally, other medications NOT listed that would interfere with the objectives of this study, pose a safety risk, or confound the interpretation of the study results (in the PI's judgement) are prohibited while a patient is participating in the study.

Should circumstances that require use of prohibited medications arise (such as anti-viral or immunomodulating medications), the PI (in consultation with the Medical Monitor and the Sponsor) will make the assessment whether to discontinue the patient from the study for the patient to receive any necessary and appropriate medical care.

#### 6.7 Self-supportive Care

Patients with COVID-19 should follow the steps as instructed by their healthcare provider (consistent with guidance from the CDC, local ordinances or practices) to care for themselves and to help protect other people in the home and community from potentially getting sick (CDC 2020).

In order to manage the symptoms associated with COVID-19, over-the-counter cough, cold, and anti-pyretic medications can be used as necessary by patients in accordance with their respective drug facts label or as instructed by their healthcare provider.

The brand name, generic name, dose, and time must be recorded for every usage occasion.

Since this in an out-patient based study, in the event of significant worsening of COVID-19-related symptoms or two sequential pulse oximetry readings of <94%, patients will be instructed to seek medical attention immediately with their treating provider or go to the emergency room. If the PI or study staff are contacted due to significant clinical worsening of COVID-19 symptoms, the patient will be triaged appropriately to the emergency room or to the outpatient treating provider per clinical judgement. The patient will be removed from the study only if hospitalized (refer to Section 5.5).

Patients will be provided with information consistent with CDC guidance regarding emergency warning signs for COVID-19 (CDC 2020) instructing them to seek medical attention immediately, if any of the following occur:

- Trouble breathing
- Persistent pain or pressure in the chest
- New confusion or inability to arouse
- Bluish lips or face

A medical provider must be contacted for any other symptoms that are severe or concerning to the patient. Patients will be instructed to call 911 if they have a medical emergency and be instructed to notify the operator that they have, or think they might have, COVID-19.

Patients will be instructed to put on a cloth face covering before medical help arrives.

#### 6.8 Dose Reductions and Holds

In the event of significant worsening of COVID-19 symptoms, severe or intolerable GI symptoms, or other attributable AEs, the KB109 dose may be down-titrated to a lower dose, or temporarily interrupted, after consultation with the Medical Monitor.

KB109 down-titration occurs by reducing the dose in a step-wise fashion. Specifically, a patient's dose can be reduced after evaluation by the PI or designee, and in consultation with the Sponsor's medical monitor or designee from 72 g/day (36 g BID) to 36 g/day (18 g BID) followed by 18 g/day (9 g BID) until the attributable AE resolves or adequate tolerability is achieved.

During the Intake Period, and at the discretion of the PI and in consultation with the Medical Monitor, patients who have down-titrated their KB109 dose may attempt to increase their dose. This up-titration can only occur at the next up-titration period. Patients can also remain at the down titrated dose, for the remainder of the intake period.

If the patient requires down-titration below 18 g/day (9 g/dose BID), then consultation with the Medical Monitor and PI or designee must occur, and the patient may be withdrawn from the study.

#### 6.9 Other Study Restrictions

Female patients of childbearing potential (as defined in accordance hospital policies and procedures at the patient's outpatient clinic visit) must indicate that they will use one highly effective form of birth control from the day of the first SP dose through the study and for 90 days after the last dose of SP.

Pregnancy status at screening for all females of childbearing potential may be collected by patient self-report or urine pregnancy test.

#### 6.10 Randomization, Stratification and Blinding

All patients deemed eligible for the study will be randomized in a 1:1 ratio to SSC + KB109 or SSC alone group using an interactive response technology (IRT) system. Randomization will be stratified by study site/center, age group (≥18 to <45 years, ≥45 to <65 years, ≥65 years), and comorbidity status (Yes, No).

The study is an open-label study; therefore, no blinding is necessary.

### 7 ADVERSE EVENT REPORTING

#### 7.1 Definitions of AEs, Period of Observation and Recording of AEs

For convenience, standard terms language included in ICH guidance relevant to the conduct of clinical trials are used in this protocol. An example of such a term included in this protocol is "treatment-emergent," included in ICH E9. Use of this standard term is not intended to connote treatment with a medicinal product administered.

An AE is defined as any untoward medical occurrence in a patient involved in a clinical study administered a SP and that does not necessarily have a causal relationship with the product. An AE can, therefore, be any unfavorable and unintended sign (including a clinically significant abnormal laboratory finding which has worsened from baseline), symptom or disease temporally associated with the use of a product, accidents, whether or not considered related to the product or study-related procedure.

All AEs are collected from the time the informed consent is signed until the end of the Follow-up Period. This includes events occurring during the Screening Period of the study, regardless of whether or not SP is administered. Where possible, a diagnosis rather than a list of symptoms should be recorded. If a diagnosis has not been made, then each sign or symptom should be listed individually. All AEs should be captured on the appropriate AE pages in the eCRF and in source documents.

All AEs must be followed to closure (the patient's health has returned to his/her baseline status or all variables have returned to normal), regardless of whether the patient is still participating in the study. Closure indicates that an outcome is reached, stabilization achieved (the PI does not expect any further improvement or worsening of the event), or the event is otherwise explained. When appropriate, medical tests and examinations are performed so that resolution of event(s) can be documented.

#### 7.2 AE Assessments

##### 7.2.1 Severity

The severity of AEs must be recorded, including the start and stop dates for each change in severity. An event that changes in severity should be captured as a new event. After initiation of KB109, worsening of events that started prior to initiating intake of KB109 must be recorded as new AEs (for example, if a patient experiences mild intermittent dyspepsia prior to dosing of KB109, but the dyspepsia becomes severe and more frequent after first dose of KB109 has been administered, a new AE of severe dyspepsia [with the appropriate date of onset] is recorded in the eCRF).

The medical assessment of severity is determined by using the following definitions:

**Mild:** A type of AE that is usually transient and may require only minimal treatment or therapeutic intervention. The event does not generally interfere with usual activities of daily living.

**Moderate:** A type of AE that is usually alleviated with specific therapeutic intervention. The event interferes with usual activities of daily living, causing discomfort but poses no significant or permanent risk of harm to the research patient.

**Severe:** A type of AE that interrupts usual activities of daily living, or significantly affects clinical status, or may require intensive therapeutic intervention.

#### 7.3 Causality

The PI or designee must make the assessment of relationship to SP for each AE. The PI or designee should decide whether, in his or her medical judgment, there is a reasonable possibility that the event may have been caused by the SP. If there is no valid reason for suggesting a relationship, then the AE should be classified as “not related”. Otherwise, if there is any valid reason, even if undetermined or untested, for suspecting a possible cause-and-effect relationship between the SP and the occurrence of the AE, then the AE should be considered “related”. The causality assessment must be documented in the source document.

The following additional guidance may be helpful:

- **Related (definitely, probably, possibly related)**: The temporal relationship between the event and the administration of KB109 is compelling and/or follows a known or suspected response pattern to that product, and the event cannot be explained by the patient’s medical condition, other therapies, or accident.
- **Not Related (definitely not, unlikely related)**: The event can be readily explained by other factors such as the patient’s underlying medical condition, concomitant

therapy, or accident and no plausible temporal or biologic relationship exists between the SP and the event.

##### 7.3.1 Expectedness

An AE will be considered unexpected if the nature, severity, or frequency of the event is not consistent with the risk information described for the study product discussed within this protocol. The study product is characterized as a prebiotic non-digestible carbohydrate, which has the potential to be poorly tolerated due to local GI when taken at higher amounts. Undesirable GI effects can include acid reflux and heartburn, burping, flatulence, colic (spasmodic abdominal pain), borborygmi (flatulence in the bowels), laxation, reduced appetite, abdominal distension or pain, nausea, abdominal rumbling or an increased defecation frequency. At still higher amounts, watery stools and diarrhea can be expected.

##### 7.3.2 Outcome Categorization

The outcome of AEs must be recorded during the course of the study in the eCRF. Outcomes are as follows:

- Fatal
- Not recovered/Not resolved
- Recovered/Resolved
- Recovered/Resolved with Sequelae
- Recovering/Resolving
- Unknown

##### 7.3.3 Symptoms of the Disease Under Study

COVID-19 related symptoms of the disease under study should not be classified as TEAEs as long as they are within the normal day-to-day fluctuation or expected progression of the disease *not including hospitalizations* and are part of the clinical data of the disease that is being collected in the study; however, clinically significant worsening of the symptoms should be recorded as a TEAE (or SAE).

##### 7.3.4 Clinical Laboratory and Other Assessments

A change in the value of a study assessment can represent an AE if the change is a clinically significant worsening from baseline. This includes abnormal assessments where there is a shift of a parameter from a normal value to an abnormal value, or a significant worsening of an already abnormal value. Clinical significance is defined as an abnormal study assessment that leads to a diagnosis or results in patient intervention such as further monitoring (excluding confirmatory repeat testing) or medical treatment.

*Note: Medications allowed and used as part of SSC to acutely relieve symptoms of COVID-19 (see Section 6.6) will be captured in the Healthcare Utilization assessment.*

If, at the end of the SP Intake Period, there are abnormal study assessments which were not present pre-treatment, the value observed closest to the start of study treatment should be used as baseline. The PI or designee should decide, based on the above criteria and the clinical condition of a patient, whether a worsening of an abnormal study assessment is clinically significant and therefore represents an AE.

##### 7.3.5 Pregnancy

All pregnancies are to be reported from the time informed consent is signed to the end of follow-up.

Any report of pregnancy for any female study patient must be reported within 24 hours to the Sponsor. Pregnant female patients must be withdrawn from the study.

Every effort should be made to gather information regarding the pregnancy outcome and condition of the infant. It is the responsibility of the PI to obtain this information within 30 calendar days after the initial notification and approximately 30 calendar days and 1-year post-partum.

Pregnancy complications such as spontaneous abortion/miscarriage or congenital abnormality are considered SAEs and must be reported as such. Note: an elective abortion is not considered an SAE.

In addition to the above, if the PI determines that the pregnancy meets SAE criteria, it must be reported as an SAE. The test date of the first positive urine human chorionic gonadotropin test or ultrasound result will determine the pregnancy onset date.

#### 7.4 SAE Procedures

##### 7.4.1 SAE Definition

A SAE is defined as any untoward medical occurrence (whether considered to be related to KB109 or not) that at any dose fulfills at least one of the following criteria:

1. Results in death
2. Is life-threatening

(*Note*: the term "life-threatening" in the definition of "serious" refers to an event in which the patient was at risk of death at the time of the event; it does not refer to an event which could hypothetically have caused death had it been more severe)

3. Requires inpatient hospitalization or prolongation of existing hospitalization
   (*Note*: hospitalizations which are the result of elective or previously scheduled surgery for pre-existing conditions, or which have not worsened after initiation of treatment, should not be classified as SAEs. For example, an admission for a previously scheduled ventral hernia repair would not be classified as an SAE; however, complication[s] resulting from a hospitalization for an elective or previously scheduled surgery that meet[s] serious criteria must be reported as SAE[s]).
4. Results in persistent or significant disability/incapacity
5. Is a congenital anomaly/birth defect
   (*Note:* congenital anomaly/birth defect in offspring of patient taking KB109 regardless of time to diagnosis)
6. Is an important medical event
   (*Note*: important medical events that may not result in death, be life-threatening, or require hospitalization may be considered an SAE when, based upon appropriate medical judgment, they may jeopardize the patient and may require medical or surgical intervention to prevent one of the outcomes listed in this definition. Examples of such medical events include allergic bronchospasm requiring intensive treatment in an emergency room or at home; blood dyscrasias or convulsions that do not result in inpatient hospitalization; or the development of drug dependency or drug abuse)

##### 7.4.2 SAE Reporting Procedures

All SAEs occurring from the time a patient signs informed consent until the end of the follow-up period must be reported to Medpace Clinical Safety within 24 hours of the knowledge of the occurrence.

To report SAEs, the site should complete the SAE form electronically in the electronic data capture (EDC) system for the study. When the form is completed, Medpace Safety personnel will be notified electronically by the EDC system and will retrieve the form.

If the event meets serious criteria and it is not possible to access the EDC system, send an email to Medpace Safety or call the Medpace SAE hotline (contact information listed below), and fax/email the completed back up paper SAE form to Medpace within 24 hours of awareness. When the EDC system becomes available, the SAE information must be entered within 24 hours of the system becoming available.

Safety Contact Information: Medpace Clinical Safety
Medpace SAE reporting line – USA:

##### 7.4.3 SAE Collection Timeframe

All SAEs (regardless of relationship to KB109) are collected from the time the patient signs the informed consent until the end of follow-up and must be reported to the Sponsor and CRO within 24 hours of the first awareness of the event (Section 7.4.2).

In addition, any SAE considered "related" to KB109 and discovered by the PI or designee at any interval after the study has completed must be reported to the CRO within 24 hours of the first awareness of the event (Section 7.4.2).

##### 7.4.4 SAE Onset and Resolution Dates

The onset date of the SAE is defined as the date the event meets serious criteria. The resolution date is the date the event no longer meets serious criteria, the date the symptoms resolve, or the event is considered chronic. In the case of hospitalizations, the hospital admission and discharge dates are considered the onset and resolution dates, respectively.

In addition, any signs or symptoms experienced by the patient after signing the informed consent form or leading up to the onset date of the SAE, or following the resolution date of the SAE, must be recorded as an AE, if appropriate.

##### 7.4.5 Fatal Outcome

Any SAE that results in the patient's death (i.e., the SAE was noted as the primary cause of death) must have 'Fatal' checked as an outcome with the date of death recorded as the resolution date. For all other events ongoing at the time of death that did not contribute to the patient's death, the outcome should be considered 'Not Resolved', without a resolution date recorded.

For any SAE that results in the patient's death or any ongoing events at the time of death, the action taken with the SP should be recorded as "dose not changed" or "not applicable" (if the patient never received SP).

##### 7.4.6 Reporting to health authorities, independent ethics committee and investigators

The PI must comply with any applicable site-specific requirements related to the reporting of SAEs (and in particular deaths) involving his/her patients to the Independent Ethics Committee

(IRB) that approved the trial. In accordance with ICH-GCP guidelines, the Sponsor will inform the PI of "findings that could adversely affect the safety of patients, impact the conduct of the trial, or alter the IRB's approval/favorable opinion to continue the trial." In line with respective regulations, the Sponsor will inform the PI of AEs that are both serious and unexpected and are considered to be related to the administered product ("suspected unexpected serious adverse reactions"). The Sponsor will send appropriate safety notifications to relevant health authorities if required in accordance with applicable laws and regulations.

The PI should place copies of safety reports in the Investigator Site File (ISF). National regulations regarding safety report notifications to the PI will be taken into account. When specifically required by regulations and guidelines, the Sponsor will provide appropriate safety reports directly to the concerned IRB and will maintain records of these notifications. When direct reporting by the Sponsor is not clearly defined by national or site-specific regulations, the PI will be responsible for promptly notifying the concerned IRB of any safety reports provided by the Sponsor and of filing copies of all related correspondence in the ISF.

### 8 STATISTICAL CONSIDERATIONS

This section outlines the statistical analysis strategy and procedures for the study. The details of the statistical methods will be provided in a separate technical document or in the Statistical Analysis Plan (SAP).

#### 8.1 Sample Size Justification

The study will enroll approximately 350 to 400 patients. The enrollment target is based on practical reasons. Assuming a 15% attrition rate, this will provide approximately 296 to 340 evaluable patients (148 to 170 per group).

The precision of the point estimate in terms of half-width of the 95% confidence interval (CI) of the AE rate is summarized for a range of evaluable number of patients and assumptions on the SP-related TEAE rates. A larger (or smaller) number of patients will provide narrower (or wider) CIs, and the CI would be narrower for rarer events with lower event rates.

**Table 4 Half-width of the 95% CI for SP-related TEAE Rate**

| Evaluable Sample Size (N/Group) | SP-Related TEAE Rate | 95% CI | Half-Width of the 95% CI |
|---|---|---|---|
| 148 | 10% | [5.8%, 16.2%] | 5.2% |
| | 20% | [14.1%, 27.7%] | 6.8% |
| | 40% | [31.9%, 48.2%] | 8.2% |
| | 60% | [51.8%, 68.1%] | 8.2% |
| 170 | 10% | [5.9%, 15.5%] | 4.8% |

| | | | |
|---|---|---|---|
| | 20% | [14.3%, 26.8%] | 6.3% |
| | 40% | [32.6%, 47.8%] | 7.6% |
| | 60% | [52.2%, 67.4%] | 7.6% |

#### 8.2 Analysis Endpoints

The study endpoints for evaluation are listed in Section 3.2.

#### 8.3 Analysis Sets

##### 8.3.1 Safety Analysis Set

The Safety Analysis Set will include all randomized patients. Patients will be included in the group based on the fact that whether or not they actually consumed any amount of KB109: patients who actually consumed any amount of KB109 will be in the SSC + KB109 group; otherwise patients will be in the SSC alone group.

##### 8.3.2 Full Analysis Set

The Full Analysis Set will include all randomized patients and have both Baseline and at least one post-Baseline endpoint observation during intake period. Patients will be analyzed in the group to which they were randomized.

#### 8.4 Statistical Methods

This section describes the statistical methods that address the primary and secondary objectives of the study. Methods related to exploratory objectives and supportive analyses will be described in a separate document such as Statistical Analysis Plan (SAP).

##### 8.4.1 Patient Disposition

The frequency and percentage of patients in each analysis set along with disposition (completed study, early termination, with breakdown for reasons for discontinuation) will be summarized overall and by group (SSC or SSC + KB109).

##### 8.4.2 Demographics and Baseline Characteristics

Patient demographics (including age, gender, race, ethnicity, weight, height, and BMI) and baseline characteristics, medical history, and concomitant medications/procedures will be summarized by group either by descriptive statistics or categorical tables. The analysis set will be provided for the Safety and Full Analysis Sets.

##### 8.4.3 Analysis of Endpoints of Interest

Adverse Events (AEs)

The analysis of AEs will be based on the Safety Analysis Set. AEs will be coded using the most recent version of Medical Dictionary for Regulatory Activities (MedDRA).

A TEAE is defined as any AE starts or worsens in severity on or after the first dose of SP is taken for SSC + KB109 group or on or after the randomization for SSC group. Adverse event summary tables will include all TEAEs, SP-related TEAEs, SAEs, TEAEs by severity, and TEAEs leading to product discontinuation. Summaries will be presented by MedDRA system organ class and preferred term using frequency counts and percentages by group and overall.

The analysis of the following endpoints will be based on the Full Analysis Set unless otherwise specified.

Time to resolution of overall 13 COVID-19 related symptoms and the 8 cardinal COVID-19 related symptoms

The Kaplan-Meier method will be used to estimate median time to resolution and resolution rate specific to each group. Kaplan-Meier graphs will be generated; quartiles and point probabilities will be calculated. Interval estimates will be calculated using 95% point-wise CIs.

Proportion of patients with reduction from Baseline (symptom present at Baseline) and proportion of patients with symptom becomes absent (symptom present at Baseline) at EOI and Follow-up for each of 13 individual COVID-19 related symptom will be summarized by group using frequencies and percentages.

Change from Baseline to EOI in overall composite scores of the 13 COVID-19 related symptoms and the 8 cardinal COVID-19 related symptoms

Baseline is calculated as the average of non-missing measurements on Day -2, -1 and 1 (before first dose of KB109), and the EOI will be calculated as the average of non-missing measurements on Day 12, 13 and 14. These measurements will be analyzed using analysis of covariance (ANCOVA) model. In this model, the response variable is the change from Baseline to EOI measure, and the model will include factors for group, visit (Day 2-4, Day 5-11, and Day 12-14), the interaction of group by visit, site, age group (≥18 to <45 years, ≥45 to <65 years, ≥65 years), and comorbidity status (Yes, No); and the Baseline value. Average of non-missing measures from multiple days will be used at each visit. The least-squares mean (LSM) change from Baseline with the associated standard errors will be displayed for each group. Estimated group differences ([SSC + KB109] – [SSC]) along with corresponding 95% confidence intervals (CIs) will also be presented for SSC + KB109 group. In addition, the mean of the overall composite score will be presented by group and by visit.

Time to resolution of fever

The Kaplan-Meier method will be used to estimate median time to resolution of fever and resolution rate specific to each group. Kaplan-Meier graphs will be generated; quartiles and point probabilities will be calculated. Interval estimates will be calculated using 95% point-wise CIs.

Proportion of patients with oxygen saturation <95% (or <98%) on Day 14 and Day 35, and proportion of patients experiencing hospital admissions during the Intake Period and Follow-up Period will be summarized by group using frequencies and percentages.

Measures collected from the Healthcare Provider Wellness Visits and Healthcare Utilizations during the Intake Period and Follow-up Period will be summarized by group based on the Safety Analysis Set using the following descriptive statistics: sample size, mean, standard deviation, median, minimum value, and maximum value or using frequencies and percentages.

Further details of the model specification, assumptions, and SAS implementation codes will be provided in the SAP.

##### 8.4.4 Subgroup Analyses

Analysis for the secondary endpoint of time to resolution of overall 13 COVID-19 related symptoms and time to resolution of overall 8 cardinal COVID-19 related symptoms, change from Baseline to EOI in overall composite score of 13 COVID-19 related symptoms and overall composite score of the 8 cardinal COVID-19-related symptoms will be conducted for the following subgroups based on the Full Analysis Set:

- Study site/center
- Age group (≥18 to <45 years, ≥45 to <65 years, ≥65 years)
- Comorbidity status (Yes, No)
- Baseline BMI subgroup (<30 kg/m$^2$, ≥30 kg/m$^2$)
- Ethnicity (Hispanic or Latino, Not Hispanic or Latino)

The Kaplan-Meier method will be used to estimate median time to resolution and resolution rate specific to each group for each subgroup. A similar ANCOVA model as the analysis of change from Baseline to EOI in overall composite score of 13 COVID-19 related symptoms and overall composite score of the 8 cardinal COVID-19 related symptoms will be performed. For each subgroup, summary statistics including mean, SD, and 95% CIs will be provided for each group.

##### 8.4.5 Extent of Exposure and Compliance

The cumulative exposure to SP, including duration of exposure and cumulative amount of SP received, will be summarized using descriptive statistics by group for the Safety Analysis Set. Intake information will be provided in a data listing.

##### 8.4.6 Interim Analysis

As large parts of the study are conducted remotely, an interim analysis to assess whether there are indications that require changes in the conduct of the study will be performed. This analysis is planned when approximately 40% of the total randomized patients have completed or discontinued prior to completion of the EOI period.

All available data for the specific endpoints will be included in the interim analysis at the data cut-off date. The results of interim analysis will not be shared with the investigators prior to the completion of the study.

Statistical details of the interim analysis will be provided in the SAP.

### 9 SUPPORTING DOCUMENTS AND OPERATIONAL CONSIDERATIONS

#### 9.1 Regulatory, Ethical, and Study Oversight Considerations

This study will be initiated only after all required documentation has been reviewed and approved by the respective IRB. The same applies for the implementation of changes introduced by amendments.

The PI will ensure that this study is conducted in full conformity with Regulations for the Protection of Human Patients of Research codified in 45 CFR Part 46, 21 CFR Parts 50 and 56, and/or the principles in the ICH E6 (R2) GCP guideline.

##### 9.1.1 Informed Consent

Informed consent is a process that is initiated prior to the individual’s agreeing to participate in the study and continues throughout the individual’s study participation. Discussion of risks and possible benefits of participation will be provided to the patients. Consent forms will be IRB-approved, and the patient will be asked to read and review the document. The PI, or designee, will explain the research study to the patients and answer any questions that may arise. All patients will receive a verbal explanation in terms suited to their comprehension of the purposes, procedures, and potential risks of the study and of their rights as research patients. Patients will have the opportunity to carefully review the consent form and ask questions prior to completing. The rights and welfare of the patients will be protected by emphasizing to them that the quality of their medical care will not be adversely affected if they decline to participate in this study. The patients may withdraw consent at any time throughout the course of the study.

The ICF will be retained in the patient's records and a copy of the ICF will be provided to the patient.

If a protocol amendment is required, then the ICF may need to be revised to reflect the changes to the protocol. If the ICF is revised it must be reviewed and approved by the responsible IRB. A determination will be made regarding whether previously consented patients need to be re-consented.

##### 9.1.2 Institutional Review Board/Ethics Committee Review

The protocol, ICF, and all patient materials will be submitted to the IRB for review and approval. A copy of the IRB approval letter must be supplied to the Sponsor prior to starting the study. Approval of both the protocol and the ICF must be obtained before any patient is enrolled. Any amendment to the protocol will require review and approval by the IRB before the changes are implemented to the study. All changes to the ICF will be IRB approved; a determination will be made regarding whether previously consented patients need to be re-consented.

A protocol change intended to eliminate an apparent immediate hazard may be implemented immediately provided that the Sponsor, the Contract Research Organization, and the IRB are immediately notified.

##### 9.1.3 Safety Monitoring Plan

Safety oversight of the study will be under the direction of the study medical monitor, overseen by the Sponsor. Detailed information on the nature of safety data review will be described in a study-specific Safety Monitoring Plan (MP).

##### 9.1.4 Quality Assurance and Quality Control

Quality Control (QC) procedures will be implemented beginning with the data entry system and data QC checks that will be run on the database will be generated. Any missing data or data anomalies will be communicated to the site(s) for clarification/resolution. Additional details may be found in the study-specific Data Management Plan.

Following written standard operating procedures, the monitors will verify that the clinical study is conducted, and data are generated, documented (recorded), and reported in compliance with the protocol, the principles of GCP in ICH E6 (R2) and any applicable regulatory requirements.

The investigational sites will provide direct access to study-related sites, source data/documents, and reports for the purpose of monitoring and auditing by the Sponsor and inspection by local and regulatory authorities.

##### 9.1.5 Direct Access to Source Data and Documents

Clinical site monitoring is conducted to ensure that the rights and well-being of human patients are protected, that the reported study date are accurate, complete, and verifiable, and that the conduct of the study complies with the currently approved protocol/amendment(s), with applicable regulatory requirements, and with GCP guidelines. The Sponsor or their designees will monitor the conduct of the study by monitoring visits and in-house data quality review. The PI will permit study-related monitoring, audits, IRB review and regulatory inspections. Direct access must be provided to the ICFs, eCRF and source documents/data, including progress notes, copies of laboratory and medical test results. The accuracy of the data will be verified by direct comparison with source documents.

##### 9.1.6 Publication

The results of this clinical study may be published or presented at scientific meetings by the Sponsor and/or PI. If this is foreseen, the PI agrees to submit all manuscripts or abstracts to the Sponsor prior to submission. This allows the Sponsor to protect proprietary information and to provide comments based on information from other studies that may not yet be available to the PI (as specified in the contract).

In accordance with standard editorial and ethical practice, the Sponsor will generally support the publication of multicenter trials only in their entirety and not as individual center data. In this case, a coordinating PI will be designated by mutual agreement.

Authorship will be determined by mutual agreement.

#### 9.2 Administrative and Legal Obligations

##### 9.2.1 Protocol Amendments

The protocol will be submitted to the IRB for review and approval prior to initiation of study. Any amendment to the protocol will require review and approval by the IRB before the changes are implemented to the study.

##### 9.2.2 Halting of Study

SP administration may be halted if the PI or the Sponsor identifies any unexpected, significant, or unacceptable risk to patients upon ongoing review of aggregate safety data. The Sponsor will inform the PI(s) promptly should the study be halted. If SP administration is halted, the PI will promptly inform the IRB and provide reason(s) for halting.

##### 9.2.3 Study Termination

This study may be suspended or prematurely terminated by the Sponsor if there is reasonable cause. Written notification documenting the reason for study suspension or termination will be provided by the Sponsor to the PI. If the study is prematurely terminated or suspended, the PI will promptly inform the IRB and will provide the reason(s) for the termination or suspension.

Circumstances that may warrant termination or suspension include, but are not limited to:

- Determination of unexpected, significant, or unacceptable risk to patients, as determined by the ongoing review of aggregate safety data by the Sponsor
- Site submits data that are consistently insufficient, incomplete, and/or unevaluable

If the study is suspended temporarily, the study may resume once concerns about safety, protocol compliance, and/or data quality are addressed and satisfy the Sponsor and/or IRB.

##### 9.2.4 Study Documentation and Archive

The site and telemedicine provider will maintain appropriate medical and research records for this study, in compliance with the protocol, the principles of GCP in ICH E6 (R2), relevant standard operating procedures, and any applicable regulatory and institutional requirements, including for the protection of confidentiality of patients. The PI must make study data accessible to the Sponsor, to other authorized representatives of the Sponsor, and to the appropriate regulatory authority inspectors.

Source data are information, original records of clinical findings, observations, or other activities in a clinical study necessary for the reconstruction and evaluation of the study. Examples of these original documents and data records include, but are not limited to, hospital records, clinical and office charts, laboratory notes, memoranda, patients' questionnaires and diaries, pharmacy dispensing records, copies or transcriptions certified after verification as being accurate and complete, X-rays, and patient files and records kept at the pharmacy and at the laboratories involved in the clinical study.

##### 9.2.5 Electronic Case Report Forms

Data collection is the responsibility of the centralized telemedicine provider and clinical study staff at the site under the supervision of the site PI. The PI is responsible for ensuring the accuracy, completeness, legibility, and timeliness of the data reported.

Clinical data will be entered directly into the eCRFs from the source documents. Data reported in the eCRF derived from source documents should be consistent with the source documents, or the discrepancies should be explained and captured in a progress note and maintained in the patient's official electronic study record.

Clinical data (including AEs, concomitant medications, and expected adverse reactions data) and clinical laboratory data will be entered into the EDC system. The data system includes password protection and internal quality checks, such as automatic range checks, to identify data that appear inconsistent, incomplete, or inaccurate.

##### 9.2.6 Record Retention

No records will be destroyed without the written consent of the Sponsor, per the terms agreed upon in the Clinical Study Agreement (CSA). It is the responsibility of the Sponsor to inform the PI when these documents no longer need to be retained.

##### 9.2.7 SP Control, Accountability, and Disposition

All used and unused SP materials, including the SP packaging (as applicable) will be sent back to the Sponsor or designee for reconciliation and destruction. SP reconciliation may take place virtually.

##### 9.2.8 Confidentiality

The Sponsor affirms the patient's rights to protection against invasion of privacy and to follow ICH/GCP guidelines and other local regulations. The study protocol, documentation, data, and all other information generated will be held in strict confidence. Except for emergency or specialist care, no confidential information concerning the study, or the data will be released to any unauthorized third party without prior written approval of the Sponsor.

The study monitor, other authorized representatives of the Sponsor, and/or representatives of the IRB may inspect all documents and records required to be maintained by the PI or designee, including but not limited to, medical records (office, clinic, or hospital) and pharmacy records for the patients in this study. The Sponsor requires the PI to permit access to such records in accordance with local laws.

The study patient's contact information will be securely stored for internal use during the study. At the end of the study, all records will continue to be kept in a secure location until records no longer need to be retained per the terms in the CSA and applicable regulatory requirements.

Study patient data provided to the Sponsor will not include the patient's contact or identifying information. Patients will be identified by an assigned unique patient identification number on eCRFs, SAE reports, and other documents submitted to the Sponsor or Sponsor designated representative.

###### 9.2.9 Disclosure of Data

Details and terms on publication and data sharing will be specified in the CSA between the Sponsor and site.

##### 9.3 Protocol Deviations

A protocol deviation is any non-compliance with the requirements of the clinical study protocol, GCP, or other study agreements. The non-compliance may be either on the part of the patient, the PI, or the study site staff. As a result of deviations, corrective actions are to be developed by the site and implemented promptly. There will be no preapproved protocol deviations in this study.

These practices are consistent with the principles in ICH E6 (R2):

- 4.5 – Compliance with Protocol
- 5.1 – Quality Assurance and QC
- 5.20 – Non-compliance

Protocol deviations relating to individual patients are to be addressed in the patient's source documents and on the appropriate eCRF (as applicable) and reported to the Sponsor. Deviations that are not patient-specific (e.g., unauthorized use of an SP outside of the study) will be reported to the Sponsor in writing and a copy of the report will be filed in the study-specific trial master file. Protocol deviations must be sent to the IRB per their guidelines. The PI and study staff are responsible for knowing and adhering to their IRB requirements. Further details about the handling of protocol deviations will be included in study oversight documents and plans.

##### 9.4 Research Use of Stored Human Samples and Data

- **Intended Use:** Samples and data collected under this protocol will be used to study the objectives described in Section 3.1.
- **Storage:** Access to stored samples will be limited. Samples and data will be stored using codes assigned throughout the study. Data will be kept in password-protected computers,

access- and role-restricted database, and sample tracking systems. Only the site's delegated study personnel and Sponsor's study team members will have access to the stored samples and data.

- **Tracking:** Samples will be tracked using the Sponsor's (or Sponsor's designated sample storage vendor's) specified sample tracking system. Disposition at the completion of the study will be conducted as follows:
  - Clinical study data will be archived as described in the Data Management Plan
  - Samples collected through this study will be stored by the Sponsor beyond completion of the study up to a period of five years after approval of the clinical study report. Study patients who request destruction of samples will be notified of compliance with such request and all supporting details will be maintained for tracking. Once a sample has been analyzed, the sample is considered consumed and cannot be destroyed. Data associated with this consumed sample will remain in the analysis dataset.

#### 9.5 Future Use of Stored Samples and Data Derived from Samples

With the patient's consent and as approved by IRBs, de-identified biological samples and data will be stored by the Sponsor or its designee. These samples and data could be used for the Sponsor's research and development activities. The Sponsor's researchers and its designees may also be provided with a code-link that will allow linking the biological samples with the phenotypic data from each patient, maintaining the de-identification of each patient's identity. The identification of each patient beyond linking of the existing samples and data will not be possible after the completion of the study.

When the study is completed, access to study data and/or samples (if available) will be provided to the Sponsor.

The Sponsor will store data derived from the analysis of these samples. After study completion, the de-identified, archived data will be stored by the Sponsor. The informed consent will include permission for the Sponsor to store or transfer data to repository vendors. De-identified or anonymized data may be published or shared with third parties.

### 10 REFERENCES

Antunes KH, Fachi JL, de Paula R, da Silva EF, Pral LP, Dos Santos AÁ, Dias GBM, Vargas JE, Puga R, Mayer FQ, et al. Microbiota-derived acetate protects against respiratory syncytial virus infection through a GPR43-type 1 interferon response. Nat Commun. 2019;10,3273.

Arpaia N, Campbell C, Fan X; Dikiy S, van der Veeken J, deRoos P, Liu H, Cross JR, Pfeffer K, Coffer PJ, Rudensky AY. Metabolites produced by commensal bacteria promote peripheral regulatory T-cell generation. Nature 2013;504(7480):451-5.

Bindels L, Delzenne N, Cani P, Walter J. Towards a More Comprehensive Concept for Prebiotics. Nature Reviews Gastroenterology & Hepatology 2015;12;303-310.

Bito H, Hamaguchi N, Hirai H. Safety evaluation of a newly-developed dietary fiber: resistant glucan mixture. J Toxicol Sci. 2016;41(1):33-44.

Burdock GA, Flamm WG. A review of the studies of the safety of polydextrose in food. Food Chem Toxicol.1999; 37(2-3):233-264.

Center for Disease Control and Prevention (CDC). Coronavirus Disease 2019 (COVID-19), Symptoms of Coronavirus. https://www.cdc.gov/coronavirus/2019-ncov/symptoms-testing/symptoms.html Accessed on May 5, 2020.

Food and Drug Administration (FDA). Science Review of Isolated and Synthetic Non-Digestible Carbohydrates (November 2016).

Grabitske HA, Slavin JL. Gastrointestinal Effects of Low-Digestible Carbohydrates. Critical Reviews in Food Science and Nutrition 2009;49(4):327-360.

Gray M. Carbohydrate digestion and absorption: role of the small intestine. New Engl J Med. 1975;292.23:1225-1230.

GRAS Notice (GRN) No. 233.
http://wayback.archiveit.org/7993/20171031051722/https://www.fda.gov/downloads/Food/IngredientsPackagingLabeli ng/GRAS/N0tice1nventory/UCM269127 .pdf
GRAS Notice (GRN) No. 436.
http://wayback.archiveit.org/7993/20171031044353/https://www.fda.gov/downloads/Food/IngredientsPackagingLabeli ng/GRAS/NoticeInventory/UCM316569 .pdf
GRAS Notice (GRN) No. 610.
https://www.fda.gov/downloads/Food/IngredientsPackagingLabeling/GRAS/NoticeInventory/ucm502983.pdf

GRAS Notice (GRN) No. 711. https://www.fda.gov/downloads/Food/IngredientsPackagingLabeling/GRAS/NoticeInventory/ucm572901.pdf

Haak, BW, Littmann, ER, Chaubard, J-L, Pickard, AJ, Fontana, E, Adhi, F, Gyaltshen, Y, Ling, L, Morjaria, SM, Peled, JU, et al. Impact of gut colonization with butyrate producing microbiota on respiratory viral infection following allo-HCT. Blood 2018;131,2978–2986.

Ichinohe, T, Pang, IK, Kumamoto, Y, Peaper, DR, Ho, JH, Murray, TS, Iwasaki, A. Microbiota regulates immune defense against respiratory tract influenza A virus infection. Proc. Natl. Acad. Sci. U.S.A. 2011;108,5354–5359.

Johns Hopkins Coronavirus Resource Center 2020, https://coronavirus.jhu.edu/ (accessed July 2020)

Kau AL, Ahern PP, Griffin NW, Goodman AL; Gordon JI. Human nutrition, the gut microbiome and the immune system. Nature 2011;474(7351),327-336.

Rosshart SP, Herz J, Vassallo BG, Hunter A, Wall MK, Badger JH, McCulloch JA, Anastasakis DG, Sarshad AA, Leonardi I, Collins N, Blatter JA, Han S-J, Tamoutounour S, Potapova S, St. Claire MBF, Yuan W, Sen SK, Dreier MS, Hild B, Hafner M, Wang D, Iliev ID, Belkaid Y, Trinchieri G, Rehermann B. Laboratory mice born to wild mice have natural microbiota and model human immune responses. Science 2019;365(6452):EAAW4361.

Schirmer M, Smeekens SP, Vlamakis H, Jaeger M, Oosting M, Franzosa EA, ter Horst R, Jansen T, Jacobs L, Bonder MJ, et al. Linking the Human Gut Microbiome to Inflammatory Cytokine Production Capacity. Cell 2016;167,1125-1136.e8.

Smith PM, Howitt MR, Panikov N, Michaud M, Gallini CA, Bohlooly-Y M, Glickman JN, Garrett WS. The microbial metabolites, short-chain fatty acids, regulate colonic Treg cell homeostasis. Science 2013;341(6145),569-73.

Sommer F, Bäckhed F. The Gut Microbiota-Masters of Host Development and Physiology. Nat Rev. 2013; 11: 227-238.

Thaiss CA, Zmora N, Levy M; Elinav E. The microbiome and innate immunity. Nature 2016;535(7610),65-74.

Trompette A, Gollwitzer ES, Pattaroni C, Lopez-Mejia IC, Riva E1, Pernot J, Ubags N, Fajas L, Nicod LP, Marsland BJ. Dietary Fiber Confers Protection against Flu by Shaping Ly6c- Patrolling Monocyte Hematopoiesis and CD8+ T Cell Metabolism. Immunity. 2018;15,48(5),992-1005.e8.

Wils D, Scheuplein RJ, Deremaux L. Safety profile of a food dextrin: Acute oral, 90-day rat feeding and mutagenicity studies. Food and Chemical Toxicology. 2008; 46: 3254-3261.

Yokishawa Y, Kishimoto Y, Tagami H. Assessment of the safety of hydrogenated resistant maltodextrin: reverse mutation assay, acute and 90-day subchronic repeated oral toxicity in rats, and acute no-effect level for diarrhea in humans. J Toxicol Sci. 2013; 38(3): 459-470.

### 11 APPENDICES

#### 11.1 Protocol Amendments

Following finalization of K031-120 Protocol Version 1 (Dated: 23 Apr 2020) the protocol received 3 revisions, culminating in Protocol Version 4.0 (Dated: 09 Dec 2020). Tabulated summaries of the substantial changes for the revisions are provided in reverse chronological order in Section 11.1.1, Section 11.1.2, and Section 11.1.3.

In addition to the changes summarized below minor editorial changes (including correction of misspellings and other inadvertent typographical errors, minor adjustments in formatting, and edits which do not affect the safety of patients or the scientific value or conduct of the study) may have been made during each revision; for brevity these are not included in the tabulated summaries.

##### 11.1.1 Protocol Version 4.0

A tabulated summary of the changes that contributed to K031-120 protocol Version 4.0 (Dated: 09 Dec 2020) is presented in Table 5.

**Table 5: Summary of Changes between Protocol Version 4.0 and Protocol Version 3.0**

| **Protocol Version 4.0 compared to Version 3.0** | | |
|---|---|---|
| **Protocol Section(s)** | **Description of the Change** | **Rationale** |
| 2.1.2 COVID-19 | Text detailing the worldwide effect of COVID-19 was updated | To more appropriately capture the impact of the pandemic at the time of the protocol amendment. |
| 1.Synopsis (Objectives/Endpoints)<br>3.2.2 Secondary Endpoints | Secondary endpoints have been updated to include:<br>• Time to resolution of overall 13 COVID-19 related symptoms which is defined as from Day 1 until the day at which the overall composite score of 13 COVID-19 related symptoms becomes 0 or 1 and remains at 0 or1 for the rest of the Intake Period and for the Follow-up Period. Overall composite score of 13 COVID-19 related symptoms is the sum of 13 COVID-19 related symptom scores (i.e., cough, chills/repeated shaking with chills, muscle pain, fever, headache, | These endpoints have been added (or moved from exploratory endpoints) due to their clinical significance. |

| Protocol Version 4.0 compared to Version 3.0 | | |
|---|---|---|
| **Protocol Section(s)** | **Description of the Change** | **Rationale** |
| | anosmia/ageusia, shortness of breath, sore throat, gastrointestinal disturbance/symptoms, diarrhea, fatigue, nasal congestion, and chest tightness (CDC 2020). Each COVID-19 symptom will be recorded by patients on a scale of 0: Absent, 1: Mild, 2: Moderately severe, 3: Very severe. The overall composite score ranges from 0 (no symptoms) to 39 (very severe).<br>• Time to resolution of overall 8 cardinal COVID-19 related symptoms which is defined as from Day 1 until the day at which the overall composite score of 8 cardinal COVID-19 related symptoms becomes 0 or 1 and remains at 0 or1 for the rest of the Intake Period and for the Follow-up Period. Overall composite score of 8 cardinal COVID-19 related symptoms is the sum of 8 cardinal COVID-19 related symptom scores (i.e., cough, chills/repeated shaking with chills, muscle pain, fever, headache, anosmia/ageusia, shortness of breath, and sore throat).<br>• Proportion of patients with reduction from Baseline (symptom present at Baseline) in each of 13 individual COVID-19 related symptom at End of Intake Period (EOI) and Follow-up.<br>• Proportion of patients with symptom becomes absent (symptom present at Baseline) at EOI and Follow-up for each of 13 individual COVID-19 related symptom.<br>• Change from Baseline to EOI in overall composite score of 13 COVID-19 related symptoms.<br>• Change from Baseline to EOI in overall composite score of 8 cardinal COVID-19 related symptoms | |

| Protocol Version 4.0 compared to Version 3.0 | | |
|---|---|---|
| **Protocol Section(s)** | **Description of the Change** | **Rationale** |
| 1.Synopsis (Methodology)<br>4.1 Overall Design | Text regarding Diagnostic COVID-19 tests was removed | There is now flexibility for sites to perform COVID-19 test that are provided by the Sponsor as part of study conduct. |
| Table 1 | Study Product Return day updated | Patients will now return the SP during the Follow-up Period as opposed to the last intake period. |
| 4.3.2 Medical History and Concomitant Medications/Procedure | Text on detailed COVID-19 information was removed. | COVID-19 information is not part of medical history. This information will not be collected as medical history information for the current condition. |
| 1. Synopsis, Eligibility Criteria, Inclusion Criteria<br>Table 1, Footnote 2<br>Figure 4<br>5.1 Inclusion Criteria | Inclusion Criterion #3 has been updated to remove time limitation of cardinal COVID-19 symptoms prior randomization. However, it does include the requirement that the patient must not report significant improvement in their cardinal COVID-19 symptoms in the 48 hours immediately prior to randomization. | Given the evolving testing paradigms and turnaround time from sample collection to COVID-19 diagnosis results in various regions and clinical sites, the study protocol is revised to allow for symptomatic patients who are positive for COVID-19 to be eligible for potential enrollment should their cardinal COVID-19 symptoms not be significantly improving at the time of randomization, irrespective of symptoms start. |
| 1. Synopsis, Eligibility Criteria, Exclusion Criteria<br>5.2 Exclusion Criteria | Exclusion Criterion #9d has been updated to include: Antacid (H2 blockers and PPIs) and antidiarrheal agents are not prohibited | These medications for concurrent use are not prohibited and the updated text reflects a clarification. |
| 6.6 Prohibited Medications/Dietary Supplements | The whole section was revised and updated | The text was revised to provide better clarity. |
| 7.3.1 Expectedness | This section was created under Section 7.3 Causality. | This sub-section was created to align with the Safety Monitoring Plan for this study. |
| 1. Synopsis (Statistical Methods)<br>8.4.3 Analysis of endpoints | Updated text on endpoint analysis. | The methodology was updated to reflect the updated secondary endpoints. |
| 8.4.4 Subgroup Analyses | Baseline BMI subgroup and Ethnicity were added. Methodology was also updated. | To identify if there are any differences in the role of the MMT in these subgroups. |
| 8.4.6 Interim Analysis | Added a planned interim analysis | As large parts of the study are conducted remotely, an interim analysis to assess whether there are indications that require changes in the conduct of the study will be performed. |
| Appendices, Section 11 | Updated to reflect summary of comparison of protocol version 4.0 to protocol version 3.0 | N/A |

##### 11.1.2 Protocol Version 3.0

A tabulated summary of the changes that contributed to K031-120 protocol Version 3.0 (Dated 14 Jul 2020) is presented in Table 6.

**Table 6: Summary of Changes between Protocol Version 3.0 and Protocol Version 2.0**

| **Protocol Version 3.0 compared to Version 2.0** | | |
|---|---|---|
| **Protocol Section(s)** | **Description of the Change** | **Rationale** |
| 1. Synopsis (Objectives/Endpoints)<br>3.2.3 Exploratory Endpoints<br>4.3.8 Patient-assessed Bedrest Time | The definition of bedrest has been broadened by removal of the stipulation that it had to be rest in the supine position to be counted. | Bedrest is not limited to a supine position only. Patients may still be resting but not lying supine. |
| 4.3 Study Procedures (Table 1 – Schedule of assessments)<br>6.9 Other Study Restrictions | Clarified that pregnancy status may be self-reported | Due to COVID-related restrictions imposed by the building's management from freely accessing facilities to enable sample collection, some clinical study sites reported that they do not have the capacity to test pregnancy at clinic. The study protocol also allows for eligibility to be determined virtually for some patients. |
| 4.3.9 Laboratory and Inflammatory Biomarkers | Clarified that blood samples and swabs may collected if feasible, and sample collection may occur at the patients' home, remotely or at the study site or clinic. | To minimize potential exposure of other patients and study staff, increased flexibility for visits and procedures to be conducted virtually has been added |
| 1. Synopsis (Eligibility Criteria)<br>1. Synopsis (Methodology)<br>List of Abbreviations<br>4.1 Overall Design<br>4.3 Study Procedures (Table 1 – Schedule of assessments)<br>4.3.9 Laboratory and Inflammatory Biomarkers | Clarified that COVID-19 testing for eligibility is part of the standard of care, and not as part of the study procedures; the types of diagnostic tests permitted to determine eligibility have also been clarified. | To avoid confusion with the nasal and oropharyngeal swabs collected for quantitative viral load assessments (research purposes only), it has been clearly stated that eligibility is based on the diagnostic COVID-19 test carried out in the patients' standard of care setting, as well as stating what diagnostic test types are permitted. |
| 4.3 Study Procedures (Table 1 – Schedule of assessments)<br>4.2.2 Physical Examination | Clarified that the physical examination required for determination of study eligibility can be a) face-to-face, b) use information from an examination conducted as standard of care (at the Investigator's discretion and provided it was conducted within 24 hours of consent and COVID-19 testing) or c) virtually | More flexibility for physical examinations at screening has been added, to minimize potential exposure of patients and study staff, and permit virtual assessments. |

| Protocol Version 3.0 compared to Version 2.0 | | |
|---|---|---|
| **Protocol Section(s)** | **Description of the Change** | **Rationale** |
| | using an abbreviated telemedicine-based physical exam. | |
| 6.1.1 KB109 | Revised chemical formula of KB109 from H-[$C_6H_{9-11}O_5$]$_n$-OH to H-[$C_6H_{10}O_5$]$_n$-OH | $H_{9-11}$ was an error; corrected to $H_{10}$ |
| 4.3 Study Procedures (Table 1 – Schedule of assessments) | Clarification that patients will receive the KaSK prior to Day 1, at which point they can begin to self-monitor temperature and oxygen saturation | To clarify that self-reported temperature and oxygen saturation are not expected until the patients have the thermometer and pulse oximeter provided in the KaSK. |
| 4.1 Overall Design (Figure 4 – Study Design) | Study schematic updated to acknowledge that SSC is applicable to the KB109 group as well as the control, and that SSC continues through the intake and follow-up periods | Correction of schematic for internal consistency with other protocol sections describing SSC. |
| Table 1 – Schedule of assessments; 4.3.5 Patient Global Impression on COVID-19 Condition (PGIC);<br>4.3.7 Healthcare Utilization | Clarify that PGIC and Healthcare utilization reporting begins on Day 2 | Schedule clarification |
| 4.1 Overall Design (Figure 4 – Study Design)<br>4.3 Study Procedures (Table 1 – Schedule of assessments) | Addition of visit windows to the SOA and study schematic | Visit windows have been added to better accommodate potential scheduling challenges while still preserving the timely collection of data at appropriate intervals. |
| 1. Synopsis (Eligibility Criteria)<br>5.1 Inclusion Criteria | Clarification of eligibility criteria related to pre-symptomatic and symptomatic COVID-19 patients | Given the evolving testing paradigms in various regions and clinical sites and given the evolving understanding of COVID-19 presentation in patients, the study protocol is revised to allow for potential enrollment of pre-symptomatic patients who may test positive for COVID-19. Allowing entry into the study in Part 2 based on early symptoms still preserves the study population intended for evaluation in this clinical trial. |
| 1. Synopsis (Eligibility Criteria)<br>5.2 Exclusion Criteria<br>6.6 Prohibited Medications/Dietary Supplements | Clarification added regarding the permitted use of prebiotics/probiotics at screening and during the study, and clarification that there are no dietary restrictions otherwise. | Although prebiotic or probiotic use may influence the patient's gut microbiome a priori, requiring stability of their use prior to study enrollment and throughout study participation minimizes confounding factors since their gut microbiome will have already established exposure |

| Protocol Version 3.0 compared to Version 2.0 | | |
|---|---|---|
| **Protocol Section(s)** | **Description of the Change** | **Rationale** |
| | | to the products prior to study start and the exposure will not change during the study. Allowing their use also recognizes the real-world approach in which this study is being conducted. |

#### 11.1.3 Protocol Version 2.0

A tabulated summary of the substantial changes that contributed to K031-120 protocol Version 2.0 (Dated 12 May 2020) is presented in Table 7.

##### Table 7: Summary of Changes between Protocol Version 2.0 and Protocol Version 1.0

| Protocol Version 2.0 compared to Version 1.0 | | |
|---|---|---|
| **Protocol Section(s)** | **Description of the Change** | **Rationale** |
| 1. Synopsis (Study Rationale) | Revised text to clarify the primary goal of the study is related to safety evaluation of KB109 and physiologic effects of KB109 in patients with mild-to-moderate COVID-19 illness. | This is a clinical food study that is designed to evaluate the safety of KB109 and describe the natural course of COVID-19 illness in the outpatient setting. The objectives, endpoints, and statistical analysis were updated to reflect this primary goal |
| 1. Synopsis (Primary Objective) | The Primary Objective has been revised to evaluate the safety of KB109 in addition to Supportive Self Care (SSC + KB109) compared to SSC alone in outpatients with mild-to-moderate COVID-19. | Clarification of the primary objective was made to align with the primary goal |
| 1. Synopsis (Secondary Objective), 3.1.2 Secondary Objective | The Secondary Objective has been modified to evaluate selected measures of health in outpatients with mild-to-moderate COVID-19. | Clarification of the secondary objective was made to align with the primary goal. |

| **Protocol Version 2.0 compared to Version 1.0** | | |
|---|---|---|
| **Protocol Section(s)** | **Description of the Change** | **Rationale** |
| 1. Synopsis (Secondary Endpoints), 3.2.2 Secondary Endpoints | Several secondary endpoints were removed or revised. A secondary endpoint was revised to change from baseline to EOI in overall composite score which is the sum of the 8 cardinal COVID-19-related symptoms scores per CDC, 2020.<br><br>Revised secondary endpoints of proportion of patients with oxygen saturation <95% and proportion of patients with oxygen saturation <98% on Day 14 to also be evaluated on Day 35.<br><br>The secondary endpoint of measures collected from the Healthcare Provider Wellness Visits was added.<br>The secondary endpoint of proportion of patients experiencing hospital admissions during the intake period was revised to also be evaluated during the follow-up period.<br><br>The secondary endpoint of Healthcare Utilizations during the Intake Period was revised to also be evaluated during the follow-up period. | Clarification of the secondary endpoints was made to align with primary goal.<br><br>The collected cardinal symptoms were updated to reflect the most up to date published information regarding the natural history of COVID-19 (CDC, 2020). |
| 1. Synopsis (Exploratory Objectives), 3.1.3 Exploratory Objectives | Several exploratory objectives were removed or revised.<br>The following exploratory objectives were removed:<br>Evaluate the effects of KB109 based on categorized composite scores of COVID-19 related symptoms.<br><br>Evaluate the effects of KB109 on measures of health during the Follow-up Period<br><br>Evaluate the effects of KB109 on gut microbiota structure and function and stool inflammatory biomarkers | Clarification of the exploratory objectives was made to align with the primary goal. |

| Protocol Version 2.0 compared to Version 1.0 | | |
|---|---|---|
| **Protocol Section(s)** | **Description of the Change** | **Rationale** |
| 1. Synopsis (Exploratory Endpoints),<br>3.2.3 Exploratory Endpoints | Several exploratory endpoints were removed or revised.<br>The exploratory endpoints related to the COVID-19 related symptom score were revised to reflect 8 cardinal COVID-19 symptoms per CDC 2020.<br><br>The exploratory endpoints of individual measures of QOL, change from Baseline to EOI in bedrest time measured as a patient-assessed daily cumulative total resting time in supine position (measured in hours), proportion of patients with improvements in patient global impression on COVID-19 condition (PGIC), and proportion of patients with temperature below 100.4 °F without an antipyretic were added. | Clarification of the exploratory endpoints was made to align with the primary goal.<br><br>The collected cardinal symptoms were updated to reflect the most up to date published information regarding the natural history of COVID-19 (CDC, 2020). |
| 1. Synopsis (Methodology),<br>4.1 Overall Design | Added text clarifying the purpose of a provider wellness visit telephone call occurring between Day 1 and Day 14.<br><br>Added text explaining that a wellness visit telephone call will be conducted on Days 21 and 28.<br><br>Removed text regarding patient’s optionality to “opt in” to a stool collection on Day 35. | Provider wellness visits were added as additional measures to assess patient safety and wellness so as to align with the primary goal of the study.<br><br>Stool samples were removed as the sponsor will no longer be evaluating microbiome taxonomy in this study. |
| 1. Synopsis (Methodology) | Added text explaining that wellness visits by telephone occurring on Days 3, 7, 10, and 14 to monitor COVID-19 health status, safety, and compliance.<br><br>Updated reference to the secure website, “TrialPace™,” in which patients will record responses to study assessments outlined in the protocol. This change was also implemented throughout the body of the protocol where applicable. | Wellness visits were added to the study to further monitor safety and health status to align with the primary goal.<br><br>Provided detail regarding the secure website used for this study. |

| Protocol Version 2.0 compared to Version 1.0 | | |
|---|---|---|
| **Protocol Section(s)** | **Description of the Change** | **Rationale** |
| 1. Synopsis (Number of Patients Planned)<br>8.1 Sample Size Justification | Revised sample size | The enrollment target is chosen for practical reasons. Assuming a 15% attrition rate, this will provide approximately 296 to 340 evaluable patients (148 to 170 per group). The change was made to align with the updated endpoints to meet the primary goal of safety and wellness analysis. |
| 1. Synopsis (Eligibility Criteria), 5.1 Inclusion Criteria, 5.2 Exclusion Criteria | Revised inclusion criterion to remove requirement specifying English or Spanish as a primary language.<br><br>Revised inclusion criteria related to defining mild-to-moderate COVID-19 illness. The phrase in inclusion criterion #5 defining mild to moderate COVID-19 with self-reported outpatient management indicated by their healthcare provided; "*defined as low-grade fever [<101.5o F], cough, discomfort with and no evidence of Pneumonia*" was eliminated.<br><br>Added exclusion criterion to exclude patients with historical documented cirrhosis or end-stage liver disease.<br><br>Updated exclusion criterion to exclude patients participating in other interventional clinical trials. | Enrollment will not be limited to English or Spanish speaking patients. The study site may accommodate patients who speak alternative languages per the study sites standards procedures.<br><br>There were eligibility criteria embedded within the inclusion criterion #5 that were redundant and therefore removed. Patients with pneumonia and in need of treatment with antibiotics will be excluded per exclusion criterion #9 and the symptoms defining patient population of fever or cough are described in inclusion criterion #4.<br><br>Patients with cirrhosis and end stage liver disease are at higher risk developing severe COVID-19 disease course and therefore, should be excluded for safety reasons. Additionally, the severity of dysbiosis in these patients may confound the study results.<br><br>Since the interventional studies may introduce confounding factors for safety and wellness evaluation, patients will not be permitted to concomitantly enroll in this study and a different interventional study. The protocol exclusion criteria have been clarified to specify exclusion of concurrent participation in an interventional clinical study. Concurrent participation in an observational study is permitted. |
| 1. Synopsis (Statistical Methods) | Statistical methods were revised to summarize changes made to Section 8 Statistical Considerations. | The changes were made to align with the sample size, updated endpoints, and study design. |

| **Protocol Version 2.0 compared to Version 1.0** | | |
|---|---|---|
| **Protocol Section(s)** | **Description of the Change** | **Rationale** |
| 1. Synopsis (Eligibility Criteria)<br><br>5.1 Inclusion Criteria<br><br>6.9 Other Study Restrictions<br><br>7.3.4 Pregnancy | Removed inclusion criterion related to requiring male patients to have non-pregnant female sexual partners. Removed test regarding birth control requirements for male study patients.<br><br>Removed text requiring partners of male patients to report pregnancy within 24 hours to the Sponsor. | Confirmation of pregnancy in female partners of male patients, birth control measures for male patients, and reporting requirements of pregnant female partners of male patients have been removed. This is supported by the KB109 GRAS designation (per this protocol). |
| 4.1 Overall Design | Updated description of study design to reflect the following changes:<br>sample size,<br><br>clarification that SARS-CoV-2 testing result from a local clinic (not the study site) may be used for eligibility,<br><br>clarification of secure website TrialPace™,<br><br>addition of wellness visits by telephone. | The change was made to align with the primary goal of safety and updated endpoints.<br><br>The update was made to clarify the sequence of events and to clarify that the COVID-19 testing is done in clinical setting.<br><br>TrialPace™ is the specific secure website where the patients record clinical status updates.<br><br>Wellness visits were added to the study to further monitor safety and health status to align with the primary goal. |
| 4.3 Study Procedures, Table 1 Schedule of Assessments | Updated the tabulated Schedule of Assessments to reflect revisions to the study assessments and corresponding study visits. | Schedule of Assessments summarizes study design updates. |
| 4.3.4.1 Patient-assessed COVID-19 Symptom Score | Revised the symptoms of COVID-19 to reflect 8 cardinal symptoms per CDC.<br><br>Patients will also report on 5 other symptoms listed. | The collected symptoms were adapted to reflect the most up to date published information regarding the natural history of COVID-19 (CDC, 2020). The symptoms were also reordered based on clinical grouping. |
| 4.3.4.2 Patient-assessed COVID-19 Signs | Section renumbered; text remains unchanged. | Administrative update. |
| 4.3.5 Patient Global Impression on COVID-19 Condition (PGIC) | Updated the likert scale to reflect 7 categories total by adding “Very much improved” and “Very much worse” at either end of the scale. | The likert scale is validated method for collection of PGIC. |
| 4.3.6 Quality of Life (QOL) Indices | QOL on day 1 is removed | QOL indices are collected on Days 14 and 35 and not on Day 1. |

| Protocol Version 2.0 compared to Version 1.0 | | |
|---|---|---|
| **Protocol Section(s)** | **Description of the Change** | **Rationale** |
| 4.3.9 Laboratory and Inflammatory Biomarkers | Clarified that quantitative viral load test will be conducted for research purposes only and not for diagnostic purposes. | Clarification that the nasal and oropharyngeal swabs are for research purposes only and not intended for clinical use, therefore patients and providers will not receive results. |
| 4.3.10 Telemedicine Visit | Added text describing telemedicine study visits. | This section clarifies the telemedicine visits. |
| 4.3.11 Healthcare Provider Wellness Visit by Telephone | Added text describing wellness visits by telephone. | Additional added measure to monitor patient health status and safety throughout the study. |
| 5.5 Withdrawal of Patients | Added text to discontinue patients from the study who require hospitalization due to worsening COVID-19. | Clarification that once hospitalized for COVID-19, the patient will be discontinued from the study because this is an outpatient study. |
| 6.2 SP Dosing, Preparation and Administration | Table 3: Dose Up-titration of the Study Product Revised the dose on Days 5-14 from 27 g BID (54 g/day) to 36 g BIG (72 g/da) | Correction of inadvertent typographical error; now corrected to be consistent with the correct dose reported in the Synopsis. |
| 6.6 Prohibited Medications/Dietary Supplements | Clarified that the PI should consult with the Medical Monitor if the patient requires the use of prohibited medications including anti-viral or immunomodulating medications for clinical management of their condition. | It is not known whether treatments in the future may be prescribed to outpatients with COVID-19 and it is possible that these unforeseen anti-viral and immumodulating medications may confound the safety and wellness of the patients enrolled in the study and therefore need to be considered individually should they prescribed by their treating physician. |
| 6.7 Self-supportive Care | Clarified text that the PI should refer the patient for follow-up clinical management of a worsening condition based on their clinical judgement. | Clarification was made to emphasize that the PI will be monitoring patient safety throughout the study. |
| 7.4.2 SAE Reporting Procedures | Clarified SAE reporting logistics. | SAEs remains the same, but the logistics have changed so that SAEs will be reported electronically instead of hand written. |
| 8.2 Hypothesis | Deleted section. | Since there is no hypothesis testing in this study, this section has been deleted. . |
| 8.3 Analysis Sets<br>8.3.1 Safety Population Analysis Set<br>8.3.2 Full Analysis Set | Changed the names of the analysis populations | Updated to better reflect the revised study design, objectives and planned analysis. |
| 8.4.1 Efficacy Population<br>8.4.2 Per protocol Population | Removed sections. | The populations to be analyzed are now described in section 8.3.1 Safety Analysis Set and 8.3.2. Full Analysis Set. |
| 8.4.3 Analyses of Endpoints of Interest | Removed Sections 8.5.3 Efficacy Analyses and 8.5.4 Safety Analysis and replaced with list of endpoints of interest that will be analyzed. | This is a clinical food study and is not designed to show efficacy of KB109. Data on endpoints of interest will be summarized as described. |

| **Protocol Version 2.0 compared to Version 1.0** | | |
|---|---|---|
| **Protocol Section(s)** | **Description of the Change** | **Rationale** |
| 8.4.4 Subgroup Analyses | Revised text to reflect the 8 cardinal COVID-19 symptoms with subgroup analyses conducted based on the Full Analysis Set. | The change was made to reflect the updated endpoint |
| Appendix 1 Optional Stool Sample Collection | Removed Appendix 1 for stool sample collection. | Stool samples were removed as the sponsor will no longer be evaluating microbiome taxonomy in this study. |
